# A Novel Barrier Device and Method for Protection against Airborne Pathogens During Endotracheal Intubation

**DOI:** 10.1101/2022.01.24.22269341

**Authors:** Julio M. Alonso, Jeffrey Lipman, Kiran Shekar

**Affiliations:** Adult Intensive Care Services and Critical Care Research Group, The Prince Charles Hospital, Brisbane, Queensland, Australia; The University of Queensland, Brisbane. Queensland, Australia; Jamieson Trauma Institute and Intensive Care Services, Royal Brisbane and Women’s Hospital, Brisbane; Nimes University Hospital, University of Montpellier, Nimes, France; Queensland University of Technology, Brisbane; University of Queensland, Brisbane; and Bond University, Gold Coast. Queensland, Australia

**Keywords:** Endotracheal intubation, SARS-CoV-2, COVID-19, Prevention, Safety

## Abstract

**Background:** The risk of SARS-CoV-2 transmission to healthcare workers increases during aerosol-generating procedures such as endotracheal intubation.

**Objectives:** We tested the effectiveness of a novel barrier mouthpiece in reducing clinician exposure to aerosols and droplets during endotracheal intubation.

**Design:** A prospective case control study was carried out, with a single operator performing eight simulated intubations with and without the device on two different high-fidelity manikin models which produced aerosols and droplets.

**Setting:** The study was performed during June 2020, at the Clinical Skills Development Service, Brisbane, Australia.

**Interventions:** Simulated scenarios included 1) intubation during cardiopulmonary resuscitation 2) intubation while pre-oxygenating via high flow nasal cannula. Photographic images were obtained during each intubation and digitally analyzed using ImageJ v2.1.0/1.53c.

**Patients:** Not applicable.

**Main outcome measures:** Aerosol and droplets were quantified using pixel counts. Overall results were expressed as means (± SD), with comparisons between groups made using a two-tailed Student’s T-test under the assumption of unequal variances. A P value of ≤ 0.05 was considered as statistically significant.

**Results:** First pass intubation was achieved in all scenarios, with and without the barrier device. Pixel counts demonstrated significant overall reduction in aerosol and droplet exposure when the barrier device was used during intubation [Mean (SD) count:509 (860) vs 10169 (11600); P=0.014]. The highest exposure risk to airborne particles was observed during simulated induction, prior to laryngoscopy and intubation.

**Conclusions:** The novel barrier device was effective in reducing environmental exposure to aerosols and droplets during intubation without negatively affecting first pass intubation. The highest risk of exposure to airborne particles was during induction, before intubation takes place. Clinical trials are indicated to further test the feasibility and efficacy of this device.

**Trial registration:** Not applicable.

**KEY POINTS:**

- This prospective, preclinical study represents a pilot trial of a novel barrier mouthpiece for reducing clinician exposure to aerosols and droplets during endotracheal intubation.
- In eight simulated intubations with and without the barrier mouthpiece, the device proved effective in reducing environmental exposure to aerosols and droplets (measured in pixels) during intubation, without negatively affecting first pass intubation.
- The novel barrier mouthpiece represents a possible solution for reducing the risk of respiratory pathogen transmission during endotracheal intubation without hampering the procedure itself, although larger preclinical and clinical trials are necessary.

## Background

From December 2019 - when the first cases of novel SARS-CoV-2 infection were described^1^ - to May 2020 alone, over 152,888 healthcare workers had already been infected by the virus^2^, accounting for up to 17.2% of hospital admissions (including healthcare workers’ households) for COVID-19^3^. Subsequent studies have confirmed that COVID-19 is an airborne disease^4,5^, pointing to an increased risk of transmission during aerosol generating procedures such as cardiopulmonary resuscitation (CPR) or endotracheal intubation (ETI), a procedure performed in up to 20% of severely ill patients with COVID-19^6^. Worldwide shortages of personal protective equipment (PPE), especially in under-resourced environments and developing countries, have led to increased concerns about inequalities regarding access to necessary protection for healthcare workers during this and other procedures^7^. On the other hand, PPE itself may not be the panacea for eliminating the risk of SARS-CoV-2 transmission during aerosol-generating procedures. In their observational cohort, Boghdadly *et. al* describe around 10% incidence of COVID-19 symptoms in healthcare workers involved in the ETI of patients with the disease, despite the majority wearing adequate PPE as defined by the World Health Organization^8^.

To provide an extra level of protection for frontline workers during intubation of COVID-19 patients, recent publications have documented the rise of initiatives such as ‘intubation teams’ comprising of experienced anesthesiologists^9,10^, *ad hoc* intubation protocols^11^, dedicated negative pressure rooms^12^, and protective barrier enclosures, of which the best-known example is the ‘aerosol box’, designed by Dr. Hsien Yung Lai, a Taiwanese anesthesiologist^13^. Further versions of the aerosol box, including the addition of a negative-pressure chamber, have also been developed^14^. However, despite initial traction and widespread use in clinical practice, subsequent studies in simulation models and clinical practice^15^ have provided evidence that these devices could pose a risk for patient safety, due to increased intubation times^16^. In addition, these devices may hamper access to the airway, and may paradoxically increase healthcare worker infection rates because of increased exposure to aerosols during removal of the barrier enclosure^17^. These findings led to the FDA withdrawing its Emergency Use Approval for protective barrier enclosures in August 2020.

To the best of our knowledge, no current alternative to protective barrier enclosures exists that offers an extra layer of protection for physicians and their teams during intubation of patients with COVID-19 or other respiratory infections, while ensuring correct access to the airway and guaranteeing the feasibility of the procedure. Therefore, we designed a barrier mouthpiece – the Airway Shield™ - which restricts the spread of aerosols and droplets during the endotracheal intubation procedure by covering the patient’s mouth, while facilitating intubation itself via a cannula that is inserted between the patient’s tongue and the palate, allowing the introduction of the laryngoscope’s blade and the endotracheal tube, and guiding the endotracheal tube towards the larynx.

The objective of our study is to evaluate the effectiveness of this novel device regarding the reduction of aerosols and droplets liberated during ETI, compared to conventional intubation without the device. Secondary objectives include assessing the feasibility of endotracheal intubation with the device, measured as first pass intubation, and quantifying the risk of aerosol and droplet spread in different moments of the procedure.

## Methods

The study was carried out at the Clinical Skills Development Service (CSDS) in Brisbane, Australia, during June 2020. This institution provides simulation-based training for healthcare professionals from the Queensland Health workforce and offers a large range of devices and venues for preclinical research.

The study was designed to evaluate Airway Shield’s efficacy in reducing droplet (airborne particles > 5μm in diameter) and aerosol (airborne particles < 5μm in diameter)^18^ exposure when performing ETI during CPR and during controlled ETI while maintaining oxygenation with High Flow Nasal Canula (HFNC), with and without Airway Shield. High-fidelity simulated ETIs were performed on manikins capable of producing droplets and aerosols. Imaging software (ImageJ) was used to measure and compare the spread of droplets and aerosols in the different scenarios.

### Ethics

Our study was carried out in accordance with the ethical principles set out in the Declaration of Helsinki. Ethical approval for this study was not required as no animals or human patients were involved.

### Device characteristics

The device tested in this study was a 3-D printed prototype, built in medical grade TPU material with a Shore A hardness durometer scale of 80, specifically chosen to give a consistency that maintains and recovers its form, while being soft enough to avoid damage of the mucosa of a patient when introduced through the mouth and placed between the palate and the tongue (Figures 1, 2 and 3). This novel barrier device has three main components: A shield, to cover the patient’s mouth; a working channel, with two openings, a proximal one, at the level of the shield, and a distal one at the other end; and a seal, to cover the proximal opening and restrict aerosol spread while permitting the insertion of the blade of a laryngoscope and the endotracheal tube through it. Additionally, two small, sealed openings at both sides of the main proximal opening at the level of the shield, provide access for catheters for the suction of fluids or aerosols during ETI (Figures 1 and 2).

**Figure 1A:**
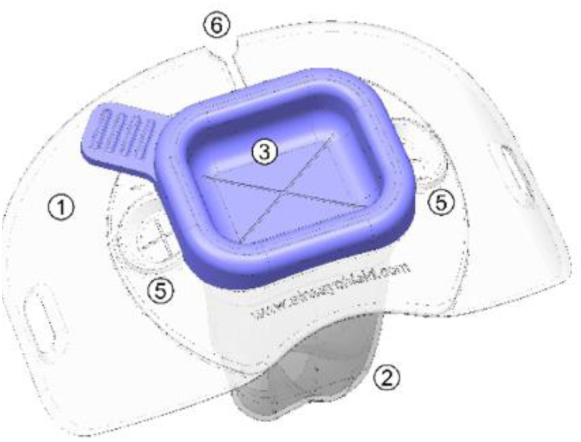
Components of the Airway Shield™ barrier mouthpiece device; Front view. 1. Sheild 2. Working channel 3. Seal of the proximal opening of the working channel 4.Distal opening of the working channel 5. Suction parts 6. Line for peel-off removal

**Figure 1B:**
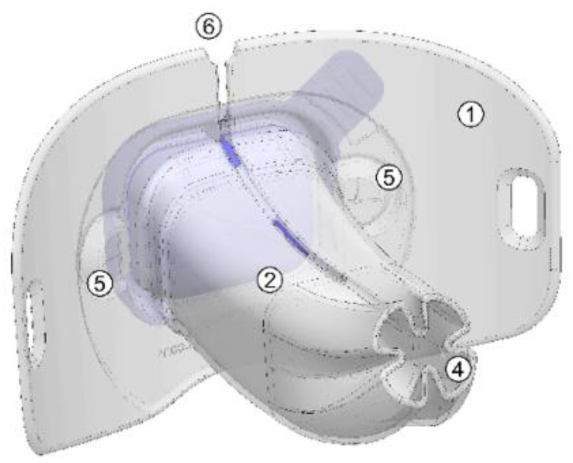
Components of the Airway Shield™ barrier mouthpiece device; Rear view. 1. Sheild 2. Working channel 3. Seal of the proximal opening of the working channel 4.Distal opening of the working channel 5. Suction parts 6. Line for peel-off removal

**Figure 1C:**
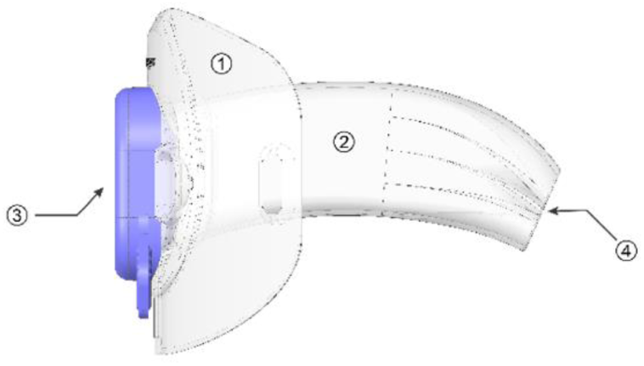
Components of the Airway Shield™ barrier mouthpiece device; Right side view. 1. Sheild 2. Working channel 3. Seal of the proximal opening of the working channel 4.Distal opening of the working channel 5. Suction parts 6. Line for peel-off removal

**Figure 2:**
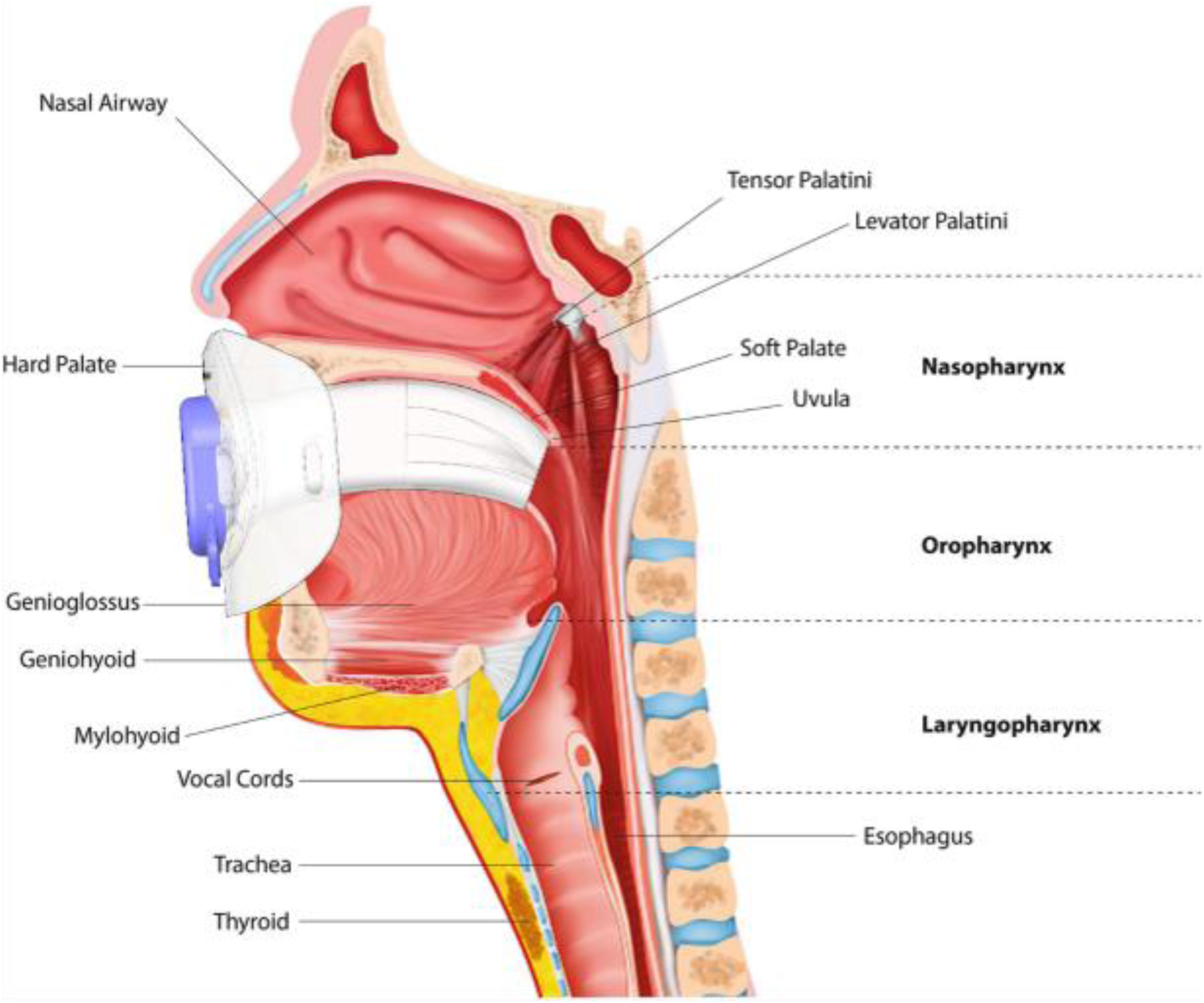
Functional position of the Airway Shield™: Working channel allocated between the tongue and the palate; shield covering the patient’s mouth

**Figure 3A:**
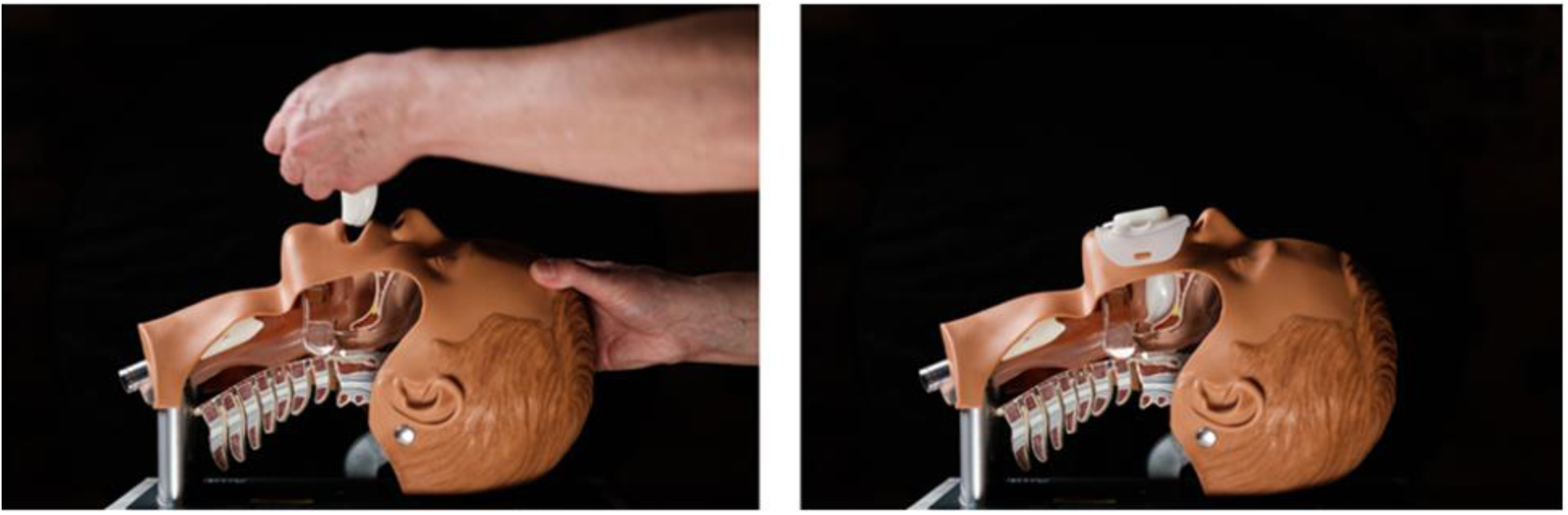
Endotracheal intubation with the aid of the Airway Shield™: Insertion.

**Figure 3B:**
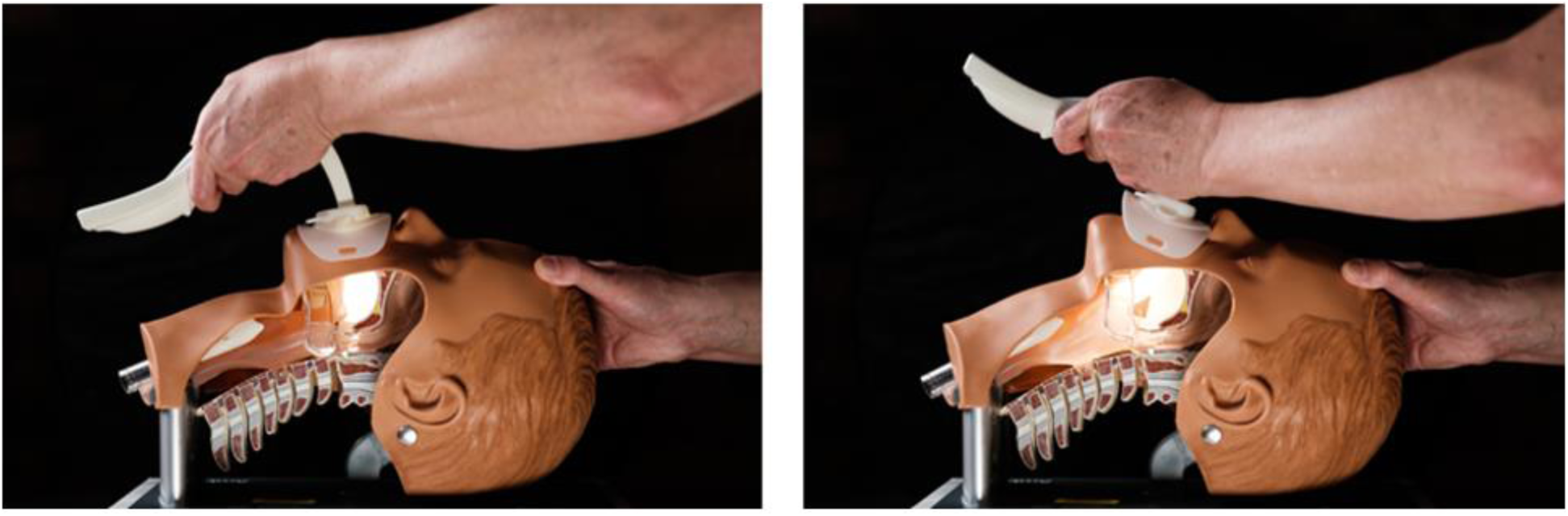
Endotracheal intubation with the aid of the Airway Shield™: Laryngoscopy.

**Figure 3C:**
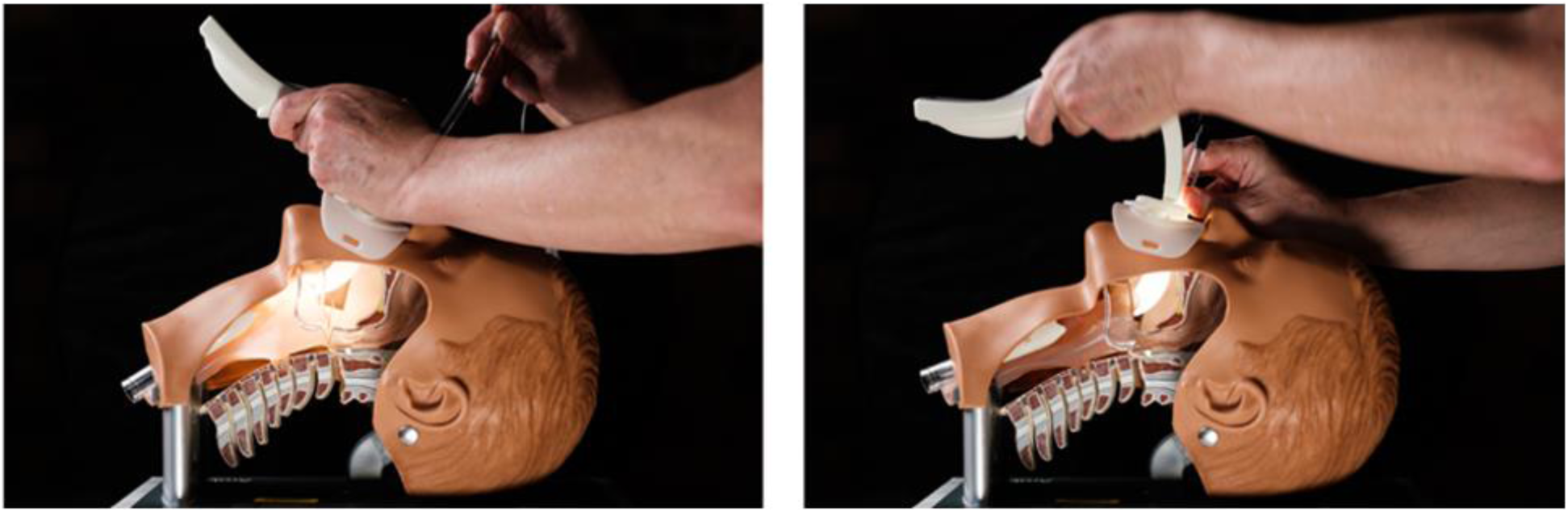
Endotracheal intubation with the aid of the Airway Shield™: Intubation.

### Technique of intubation with the novel device

Endotracheal intubation with the Airway Shield is performed in 3 steps. First, during induction, the device is placed: the working channel is introduced into the mouth of the patient following the palate, until the shield covers the mouth. Second, ETI is performed: as in a standard ETI, the video-laryngoscope blade is introduced first to obtain a view of the larynx, followed by the ETT (endotracheal tube), in this case following the blade through the working channel, towards the larynx. Finally, the Airway Shield is removed: once the ETT is in place and the ventilator is connected, the device is peeled away from the midline and removed. (Figure 3 and Additional File 2: multimedia content).

### Manikin preparation

For this study, we designed two different manikin models to produce aerosols and droplets. To simulate and evaluate the spread of aerosols during the intubation procedure, a resuscitation manikin (Megacode Kelly, Laerdal) was modified by connecting a nebulizer (Aerogen Solo, Aerogen) for inhalation to the reservoir bag of a self-inflating bag connected to the lungs, and powered by a ventilator (Servo U, Maquet), thus permitting the nebulization of an ultraviolet light-sensitive fluid into the simulated airway. Continuous airflow (6L/minute) was applied to the nebulizer to ensure sufficient aerosol visualization. To evaluate the spread of droplets, an atomizer connected by a long tubing to a syringe containing colored fluid (green dye mixed with water) was placed at the level of the manikin’s oropharynx, pointing towards the mouth (Additional File 1: Manikin Preparation).

### Simulated endotracheal intubation

We tested the device for the environmental exposure of both aerosols and droplets in two different scenarios – intubation during cardiopulmonary resuscitation (CPR) and intubation during oxygenation with high flow nasal cannula (HFNC) – and compared the results with those obtained in the same scenarios during conventional intubation. A total of eight simulated intubations were performed by a single operator, with first pass intubation recorded as a dichotomous variable (yes/no). The same sequence of simulations was followed for the scenarios evaluating the spread of droplets. (Further documentation on the complete sequence of events is available in Appendix C and D) During the simulations, photographs were taken at a rate of 6 frames per second to capture data on the spread of aerosols and droplets at three moments of each simulated clinical scenario: just before introduction of the laryngoscope into the manikin’s mouth (corresponding to anesthetic induction); during initial laryngoscopy; and during the introduction of the endotracheal tube. The spread of aerosols and droplets was documented by photographs taken from a fixed angle against a dark background. In order to quantify and compare results, the photograph showing the highest spread of aerosols and droplets was chosen for each scenario. All photographs measured 1920 × 1280 pixels.

### Data analysis

Images from each scenario were selected according to the maximum count of droplets or aerosols in each setting and were converted from color to black and white in order to permit digital analysis. A manual polygonal selection of the areas in which aerosols and droplets were visualized was carried out. Captured data were analyzed with detection and automatic count of aerosols and droplets using ImageJ v2.1.0/1.53c, an open-source package for the processing and analysis of scientific images.

Aerosol and droplets were quantified using pixel counts. Overall results were expressed as means (± SD), with the comparison between groups made using a two-tailed Student’s T-test under the assumption of unequal variances. A P value of 0.05 or less was considered to indicate statistical significance. Statistical analysis of the results was carried out using R version 4.1.0 (© 2021 The R Foundation for Statistical Computing).

## Results

24 images were selected for digital analysis using automatic pixel counts (Figures 4 and 5 and Additional File 3), after previous manual selection of the area in which aerosols or droplets had been detected. Pixel counts demonstrated significant overall reduction of aerosols and droplets during ETI in high-fidelity clinical simulations with Airway Shield compared to intubation without the device (509 (859.96) vs 10168.91 (11600.63); P=0.014), as shown in Table 1. When analyzed by subgroups (Table 2), Airway Shield reduced the spread of aerosols by 12-fold on average (P=0.045). The spread of droplets was reduced by an average of 43-fold, although this result was not statistically significant.

**Table 1:**
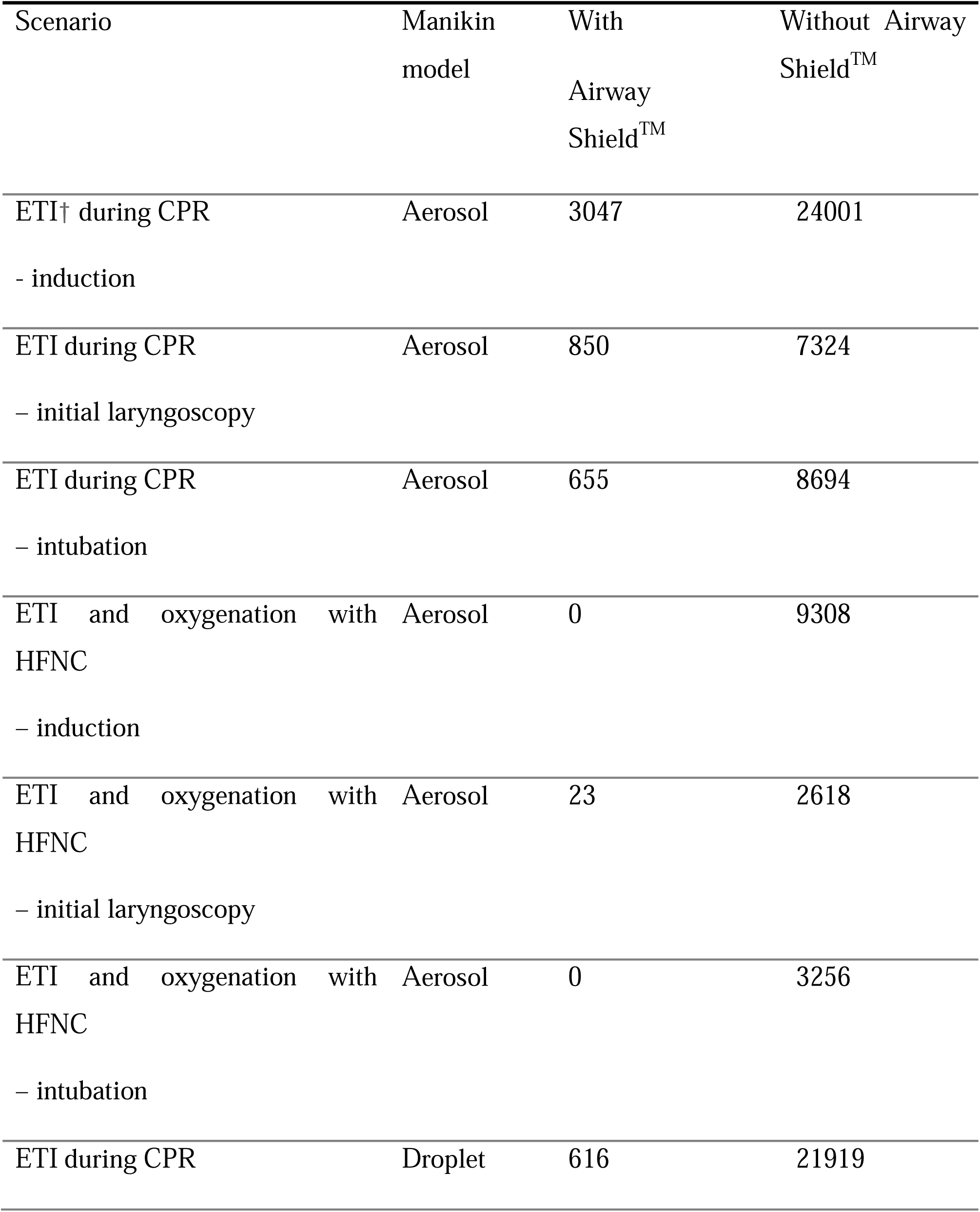
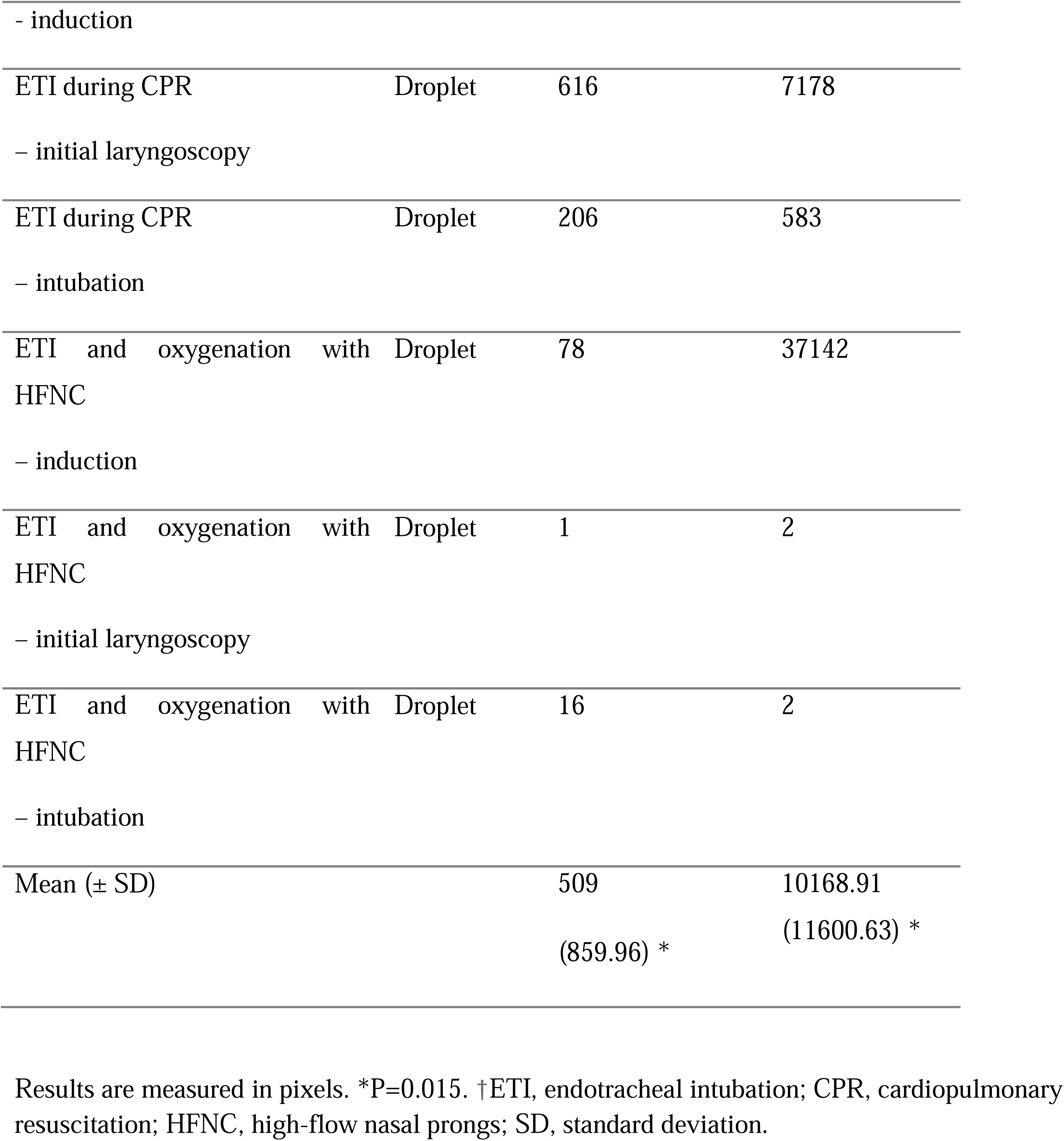
Aerosol and droplet count in high-fidelity clinical simulations of endotracheal intubation (ETI)

**Table 2:**
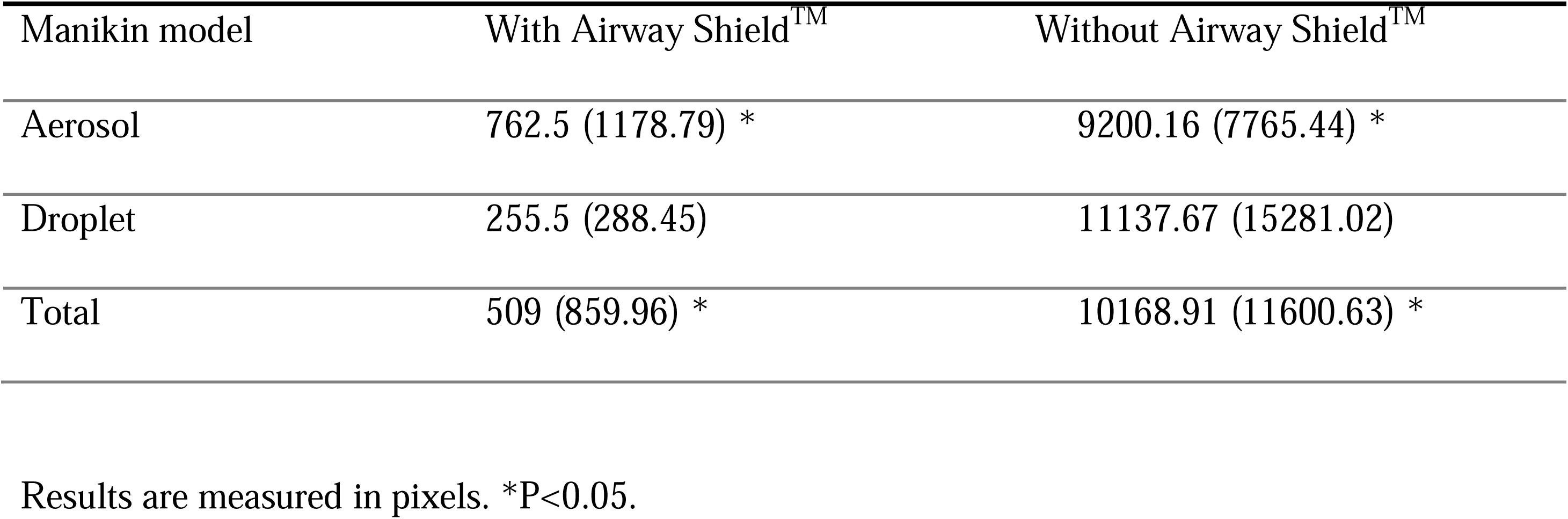
Overall comparison of aerosol and droplet counts

| Manikin model | With Airway Shield<sup>TM</sup> | Without Airway Shield<sup>TM</sup> |
| --- | --- | --- |
| Aerosol | 762.5 (1178.79) * | 9200.16 (7765.44) * |
| Droplet | 255.5 (288.45) | 11137.67 (15281.02) |
| Total | 509 (859.96) * | 10168.91 (11600.63) * |

Results are measured in pixels. \*P<0.05.

**Figure 4:**
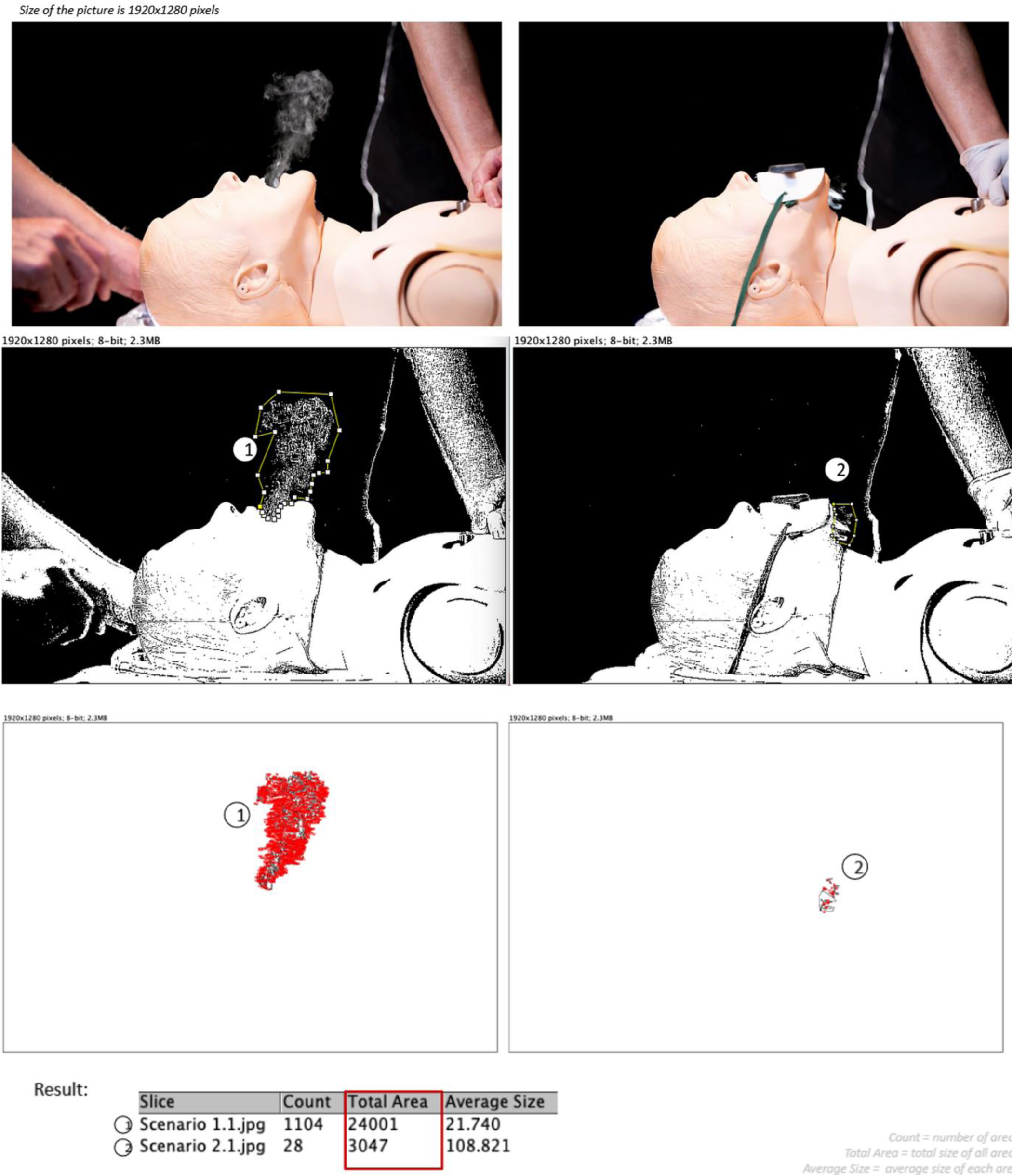
Simulated induction during ETI and CPR in aerosol manikin model.

**Figure 5:**
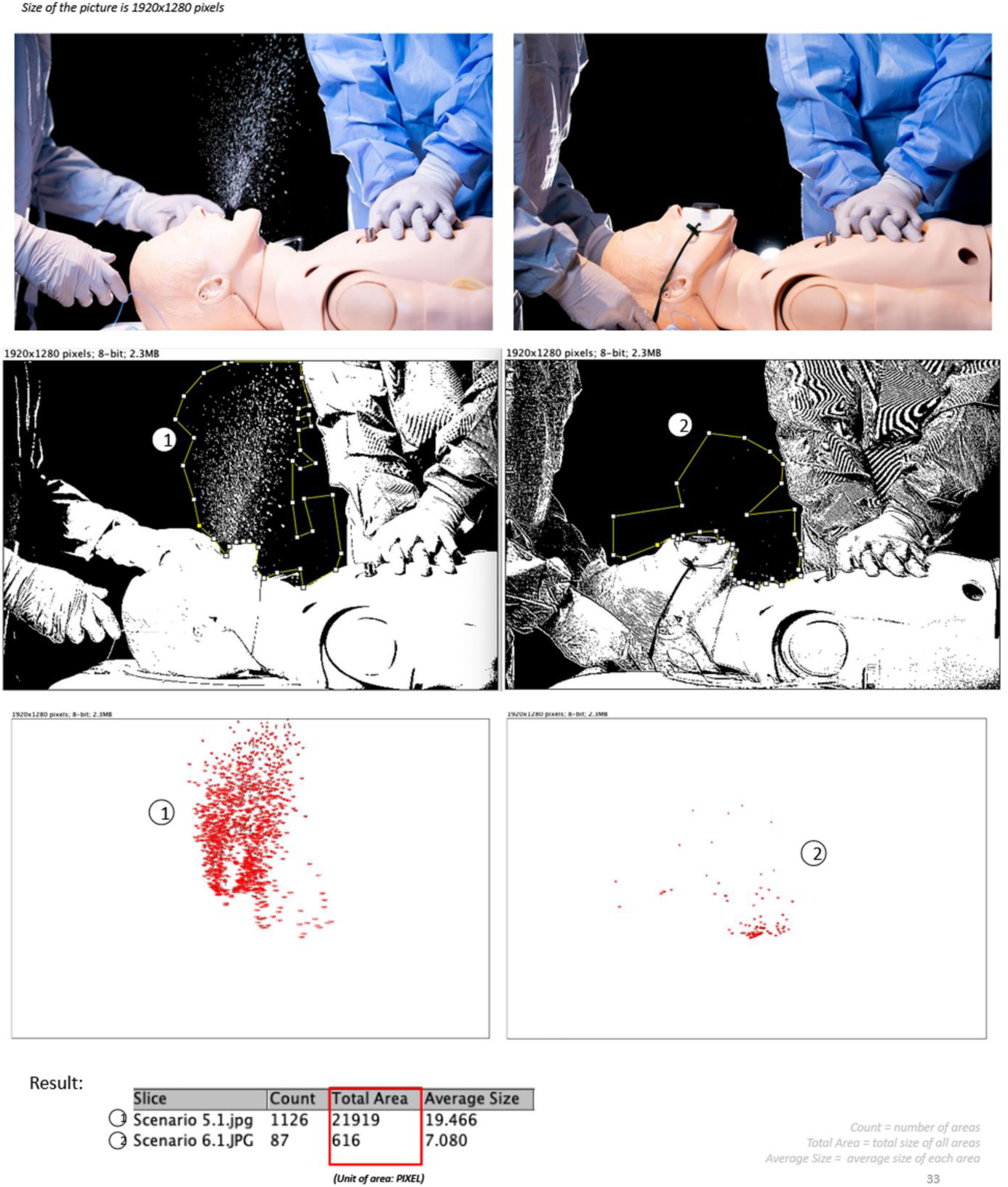
Simulated induction during ETI with HFNC oxygenation in droplet manikin models.

First pass success was achieved in all clinical scenarios, both with and without Airway Shield. The operator did not describe difficulty while carrying out the procedure in any of the simulated intubation scenarios.

Regarding the risk of aerosol and droplet spread during the different moments of the endotracheal intubation procedure, highest counts of airborne particles were observed during simulated induction, before carrying out initial laryngoscopy and intubation (Figure 6). This difference was maintained in both CPR and HFNC scenarios, independently of whether Airway Shield was used.

**Figure 6:**
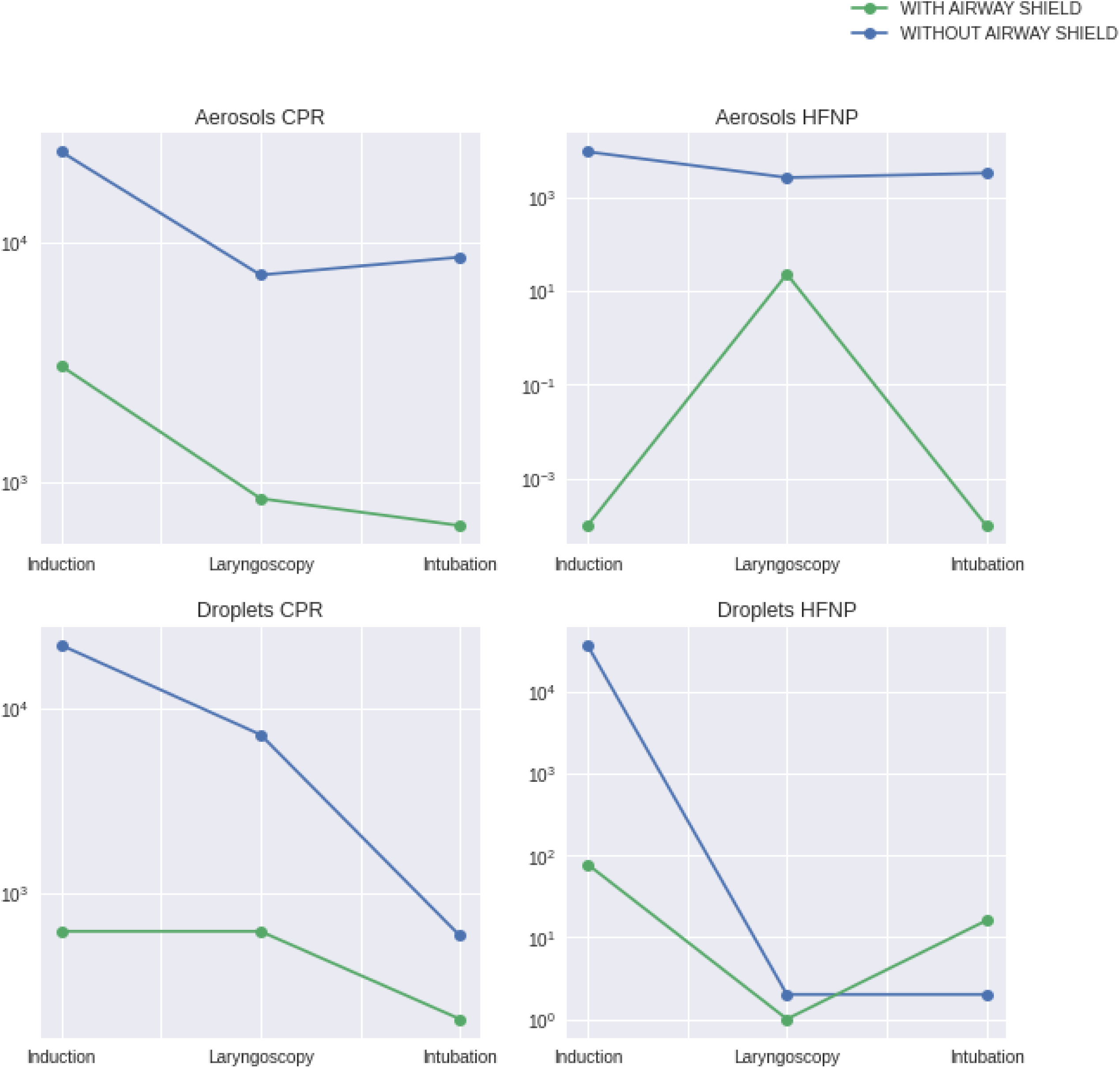
Risk of aerosol and droplet spread during different moments of the intubation procedure. Aerosol and droplet counts, in pixels, are depicted in the logarithmic scale.

## Discussion

The primary objective of our study was to test Airway Shield’s potential for reducing the spread of aerosols and droplets during ETI, as compared to intubation without the device. We simulated clinical scenarios in specifically designed manikin models and used automatic pixel counts as a surrogate marker of the reduction in spread of droplets and aerosols generated by patients in real-life settings. Our results showed that this novel barrier mouthpiece demonstrated significant overall reduction of airborne particles, while permitting a consistent first pass success rate in all simulated intubations. These findings confirm Airway Shield’s effectiveness as a barrier device against aerosols and droplets, while at the same time permitting successful ETI.

Our results offer a novel perspective on barrier devices designed to reduce exposure to airborne particles during intubation. This study is the first to present a specifically designed mouthpiece which demonstrates effective protection from aerosols and droplets while permitting successful ETI. The novelty of this device stems from the fact that it successfully covers the patient’s mouth without impeding first pass intubation.

Regarding the risk of aerosol and droplet spread during the different moments of the endotracheal intubation procedure, our study demonstrated a consistently higher count of airborne particles during induction in all but one scenario. To our knowledge, this study is the first to address the risk of exposure to aerosols and droplets at specific moments of the procedure. Although the number of experiments is too small to permit statistical comparison, we raise the hypothesis that induction may be the moment of highest risk for healthcare professionals carrying out the intubation procedure.

Our study has four main limitations. First, it is a preclinical study in manikin models, using pixel counts as surrogate markers for the aerosols and droplets generated by patients during ETI. Second, the possibility of small variations in the flow of aerosols and droplets produced by the manikins cannot be completely eliminated. Third, the manual polygon selection process may affect reproducibility of the results. Finally, the number of simulated scenarios is relatively small; however, the variations in aerosol and droplet spread are large enough to permit statistically significant conclusions.

## Conclusions

Our study suggests that Airway Shield, a novel barrier mouthpiece, is effective in reducing the spread of aerosols and droplets during ETI, while permitting successful first pass intubation. The results offer a novel perspective on barrier devices and open the door to the possibility of using a mouthpiece to protect healthcare workers during ETI, an aerosol generating procedure. Our findings also generate the hypothesis that the moment of highest risk of exposure to airborne particles is, in fact, during induction, before the actual intubation takes place.

Further research is necessary to confirm these findings, including larger simulation studies and clinical trials to evaluate safety and efficacy of Airway Shield.

## Data Availability

All data produced in the present study are available upon reasonable request to the authors.

## Assistance with the article

The authors would like to thank the ‘Clinical Skills Development Service’ in Brisbane and Clinton Henderson for their contribution setting up the manikins used for the study, Cambell Smyth for his contribution in the CAD design of the device and the drawings presented in this article, Joanne Zuo for her contribution with the digital data analysis, Jose Medrano for his suggestions for the design of the study, and Sergio Lordao and Michael Minoza for their contribution taking the photos and video.

## Presentations

None

## Additional Files

Additional File 1: Manikin Preparation

Additional File 2: Multimedia Content

Additional File 3: Dataset with all 24 shots from the 8 simulated scenarios during CPR and HFNC in droplet and aerosol manikin models performed for the study. Figures with original color photos, black and white photos, and digital pixel counts.

