## Supplemental File 1 for "A Novel Barrier Device and Method for Protection against Airborne Pathogens During Endotracheal Intubation"

Additional File 1: Manikin Preparation

1A. Modified resuscitation manikin to simulate aerosol spread

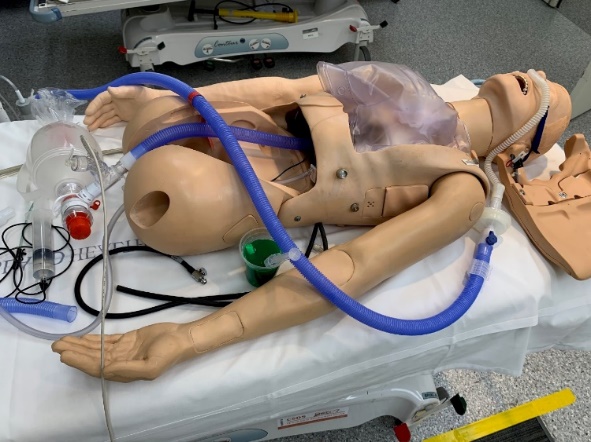

1B. Nebulizer connected to the lungs of a manikin to simulate aerosol spread

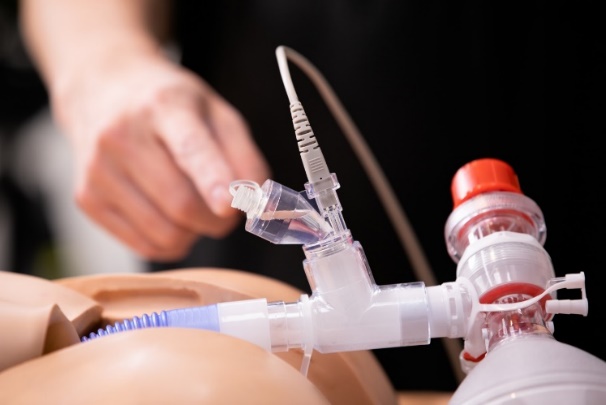

1C. Atomizer placed at the oropharynx of a manikin to simulate droplet spread

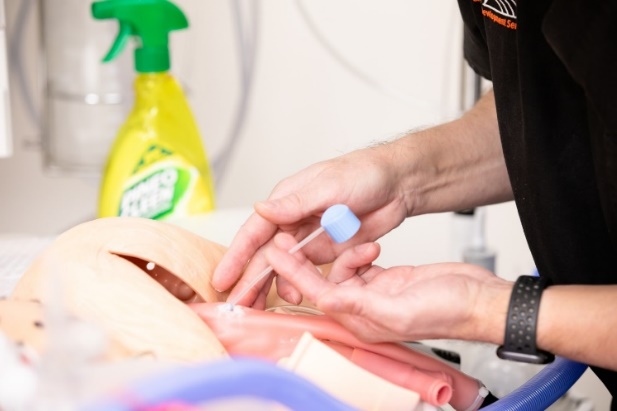
