## Supplemental File 2 for "A Novel Barrier Device and Method for Protection against Airborne Pathogens During Endotracheal Intubation"

**Additional File 2: Multimedia content**

APPENDIX A: Video explainer of how the Airway Shield^TM^ works. <https://vimeo.com/574764379/6b2a1afde7>

APPENDIX B: Video of an endotracheal intubation performed with the aid of the Airway Shield^TM^ in a manikin with sagittal section <https://vimeo.com/581728908/8bd33e0a71>

APPENDIX C: Video with all 8 simulated scenarios during CPR and HFNC in droplet and aerosol manikin models performed for the study. <https://vimeo.com/578354179/600c988f9a>
