## Supplemental File 3 for "A Novel Barrier Device and Method for Protection against Airborne Pathogens During Endotracheal Intubation"

Appendix A
Aerosol and Droplets Simulation Study
Scenarios

### Airway Shield$^{TM}$

#### A Novel Barrier Device and Method for Protection against Airborne Pathogens during Endotracheal Intubation

#### Study Summary

(A simulation study on manikin models)

Evaluation of the effect of the use of the Airway Shield$^{TM}$ to reduce droplet and aerosol spread during chest compressions and intubation

Dr. Julio M Alonso

Clinical Skills Development Service. Brisbane, Australia

### Scenarios

Endotracheal Intubation (ETI) with and without the Airway Shield$^{TM}$

TWO scenarios in each of following settings

1. Aerosol spread during ETI in CPR Scenarios
2. Aerosol spread during ETI with High Flow Nasal Cannula (HFNC) Scenarios
3. Droplet-spread during ETI in CPR Scenarios
4. Droplet-spread during ETI with High Flow Nasal Cannula (HFNC) Scenarios

### Aerosol spread during ETI in CPR Scenario

**(Slides 6 - 17)**

Scenario 1 - Aerosol-spread during ETI in compression-only-CPR WITHOUT the AirwayShield™

1.1 During compression-only-CPR just before ETI

1.2 During compression-only-CPR and laryngoscopy

1.3 During compression-only-CPR and laryngoscopy and intubation

Scenario 2 - Aerosol-spread during ETI in compression-only-CPR WITH the AirwayShield™

2.1 During compression-only-CPR just before ETI

2.2 During compression-only-CPR and laryngoscopy through the 'AirwayShield™'

2.3 During compression-only-CPR and laryngoscopy and intubation through the 'AirwayShield™'

### Aerosol spread during ETI with High Flow Nasal Cannula (HFNC) Scenarios **(Slides 18 - 29)**

Scenario 3 - Aerosol-spread during ETI with HFNC and WITHOUT the AirwayShield$^{TM}$

3.1 During intubation with HFNC O2 just before ETI

3.2 During intubation with HFNC O2 and laryngoscopy

3.3 During intubation with HFNC O2 and laryngoscopy and intubation

Scenario 4 - Aerosol-spread during ETI WITH HFNC and the AirwayShield$^{TM}$

4.1 During intubation with HFNC O2 just before ETI

4.2 During intubation with HFNC O2 and laryngoscopy through the 'AirwayShield$^{TM}$'

4.3 During intubation with HFNC O2 and laryngoscopy and intubation 'AirwayShield$^{TM}$'

### Droplet-spread during ETI in CPR Scenarios

**(Slides 30 - 41)**

Scenario 5 - Droplet-spread during ETI in compression-only-CPR WITHOUT the AirwayShield™

5.1 During compression-only-CPR just before ETI

5.2 During compression-only-CPR and laryngoscopy

5.3 During compression-only-CPR and laryngoscopy and intubation

Scenario 6 - Droplet-spread during ETI in compression-only-CPR WITH the AirwayShield™

6.1 During compression-only-CPR just before ETI

6.2 During compression-only-CPR and laryngoscopy through the 'AirwayShield™'

6.3 During compression-only-CPR and laryngoscopy and intubation through the 'AirwayShield™'

### Droplet-spread during ETI with HFNC Scenarios

**(Slides 42 - 53)**

Scenario 7 - Droplet-spread during ETI with HFNC WITHOUT the AirwayShield™

7.1 During intubation with HFNC O2 just before ETI

7.2 During intubation with HFNC O2 and laryngoscopy

7.3 During intubation with HFNC O2 and laryngoscopy and intubation

Scenario 8 - Droplet-spread during ETI WITH HFNC and AirwayShield™

8.1 During intubation with HFNC O2 just before ETI

8.2 During intubation with HFNC O2 and laryngoscopy through the 'AirwayShield™'

8.3 During intubation with HFNC O2 and laryngoscopy and intubation through the 'AirwayShield™'

### Scenario 1.1 VS 2.1

*Size of the picture is 1920x1280 pixels*

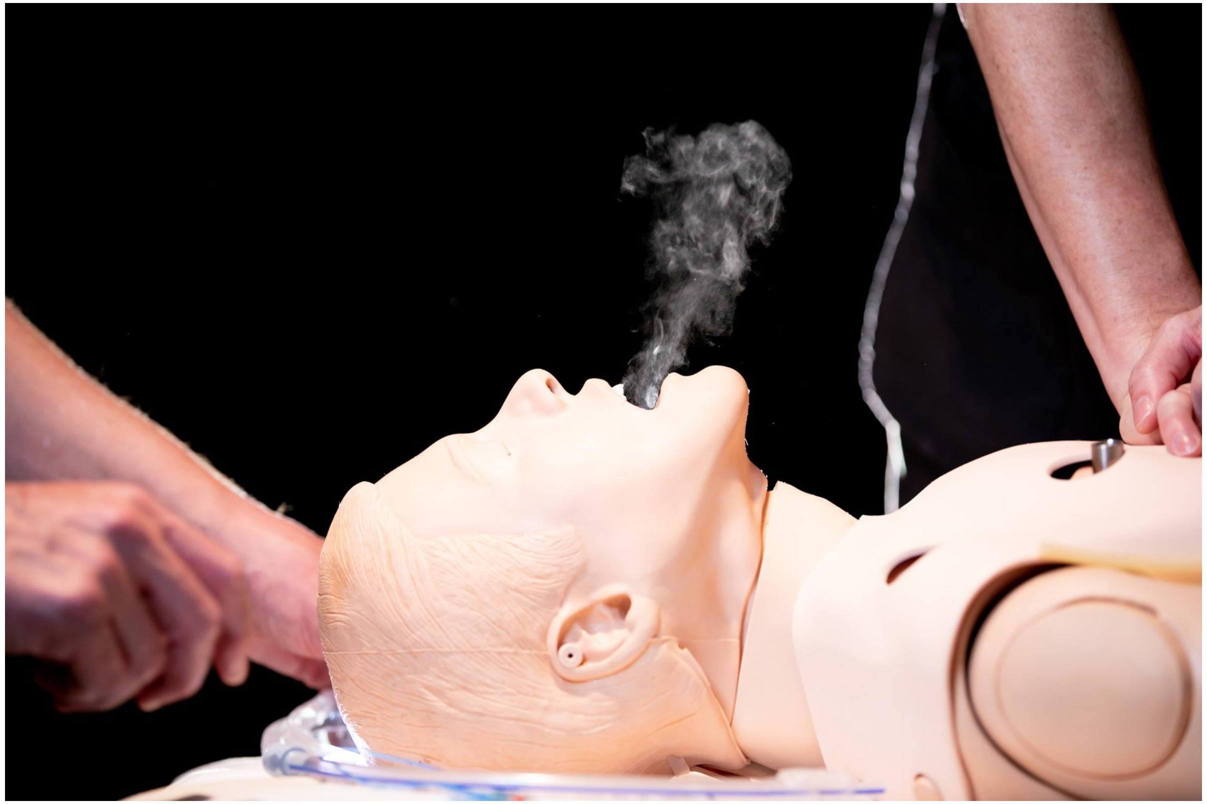

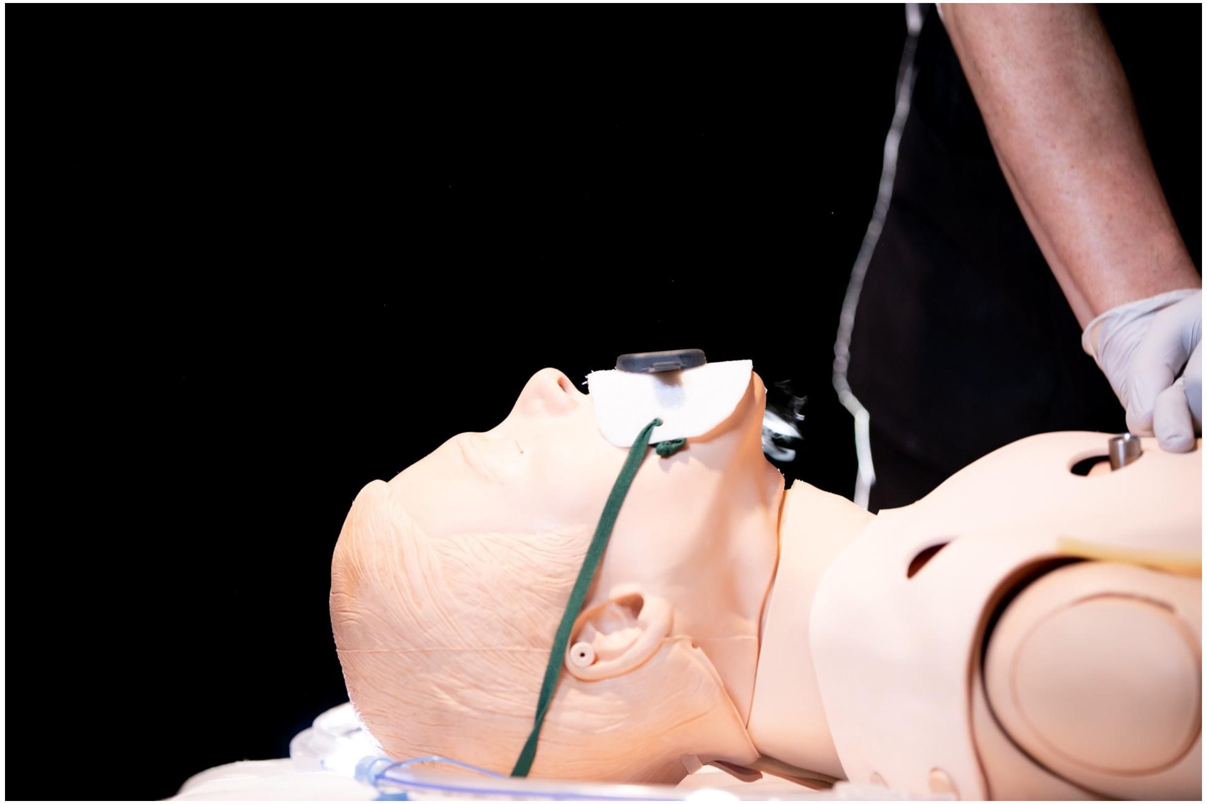

### Scenario 1.1 VS 2.1

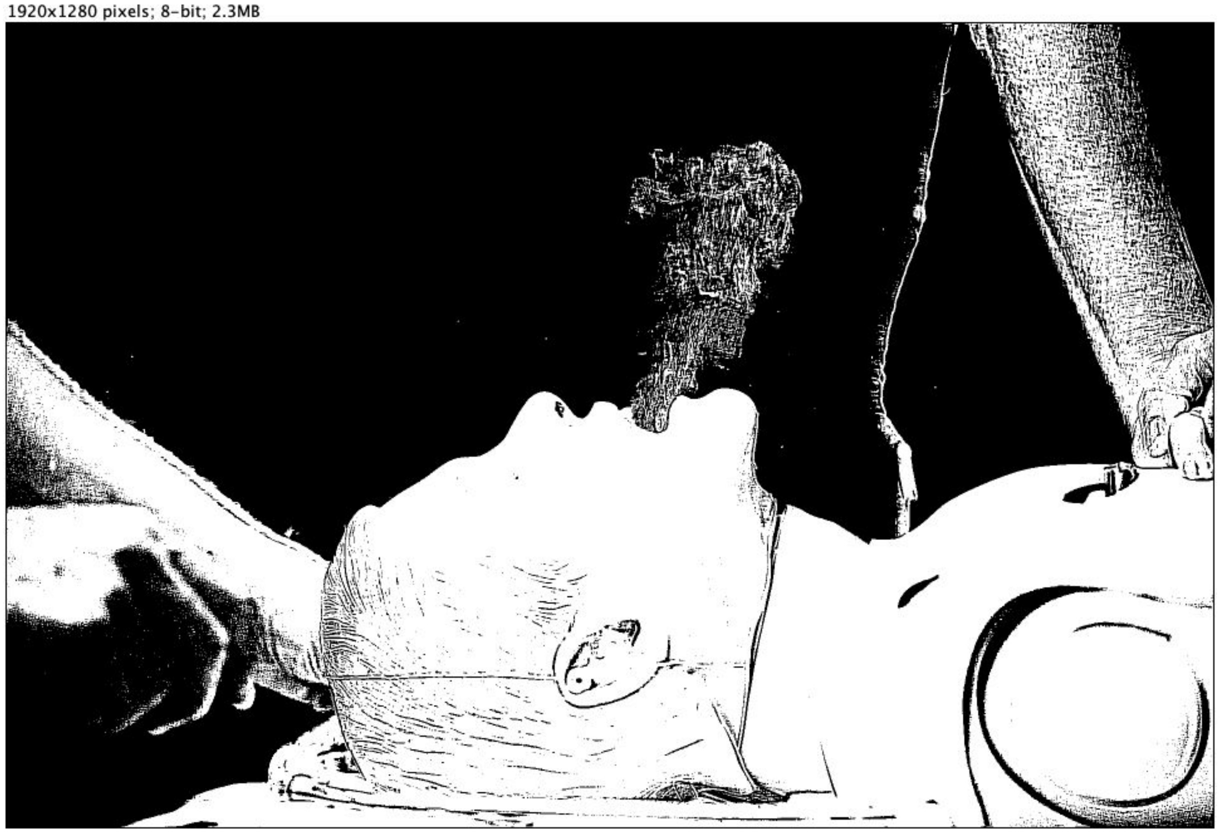


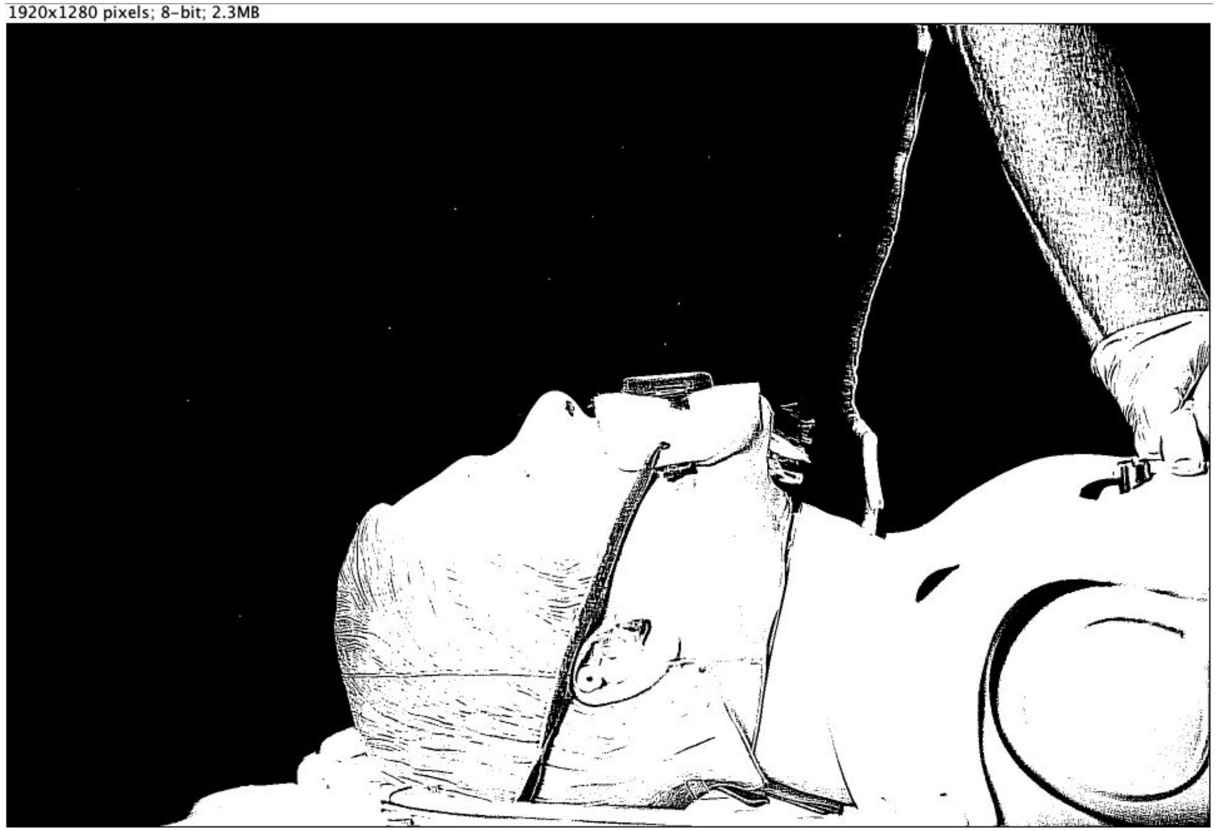

### Scenario 1.1 VS 2.1

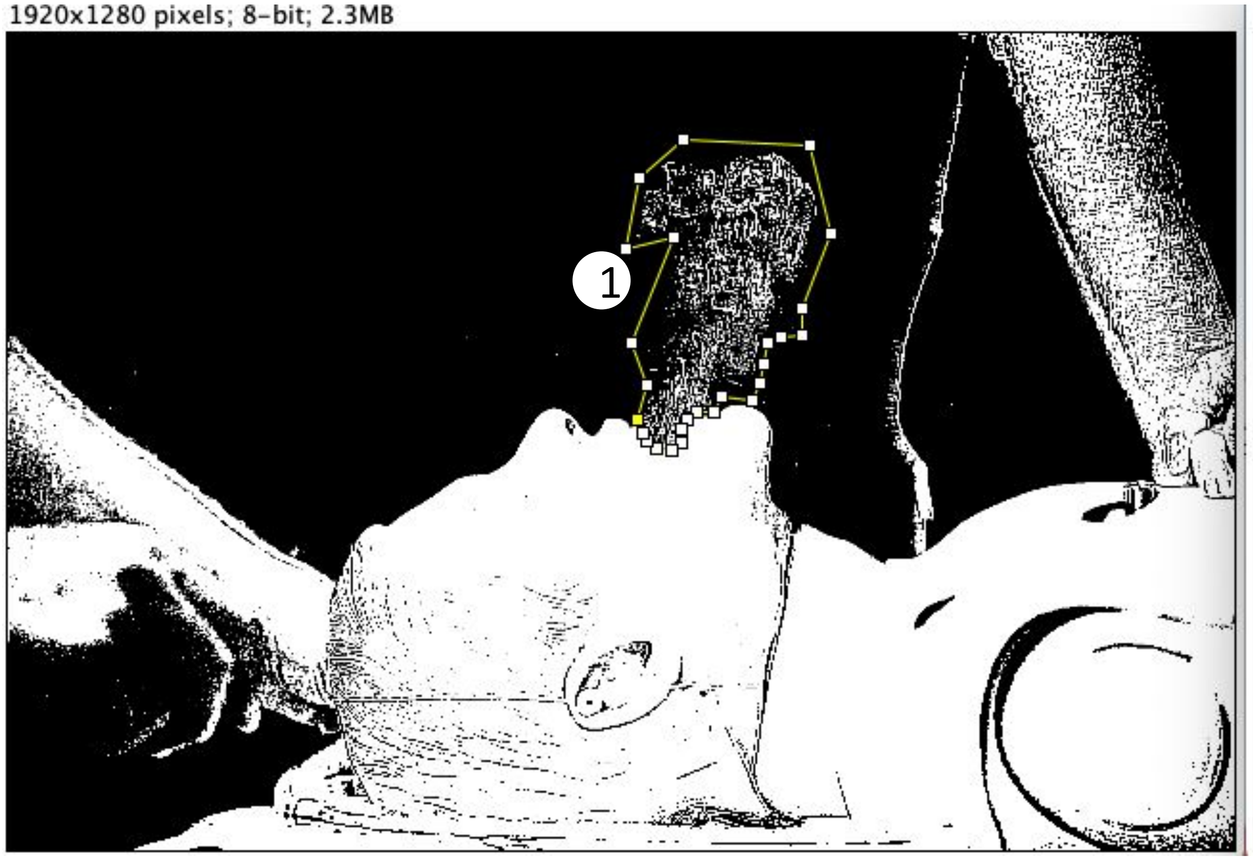


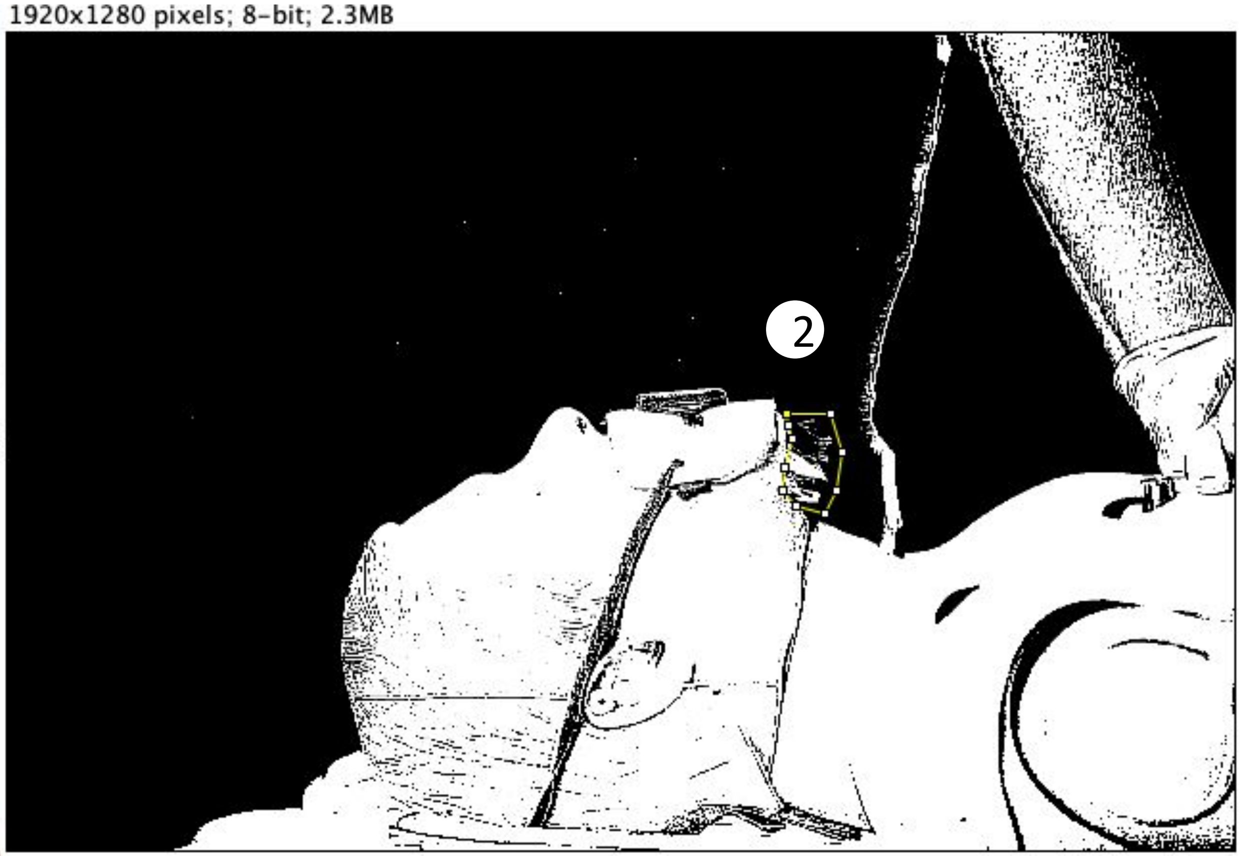

### Scenario 1.1 VS 2.1

1920x1280 pixels; 8-bit; 2.3MB

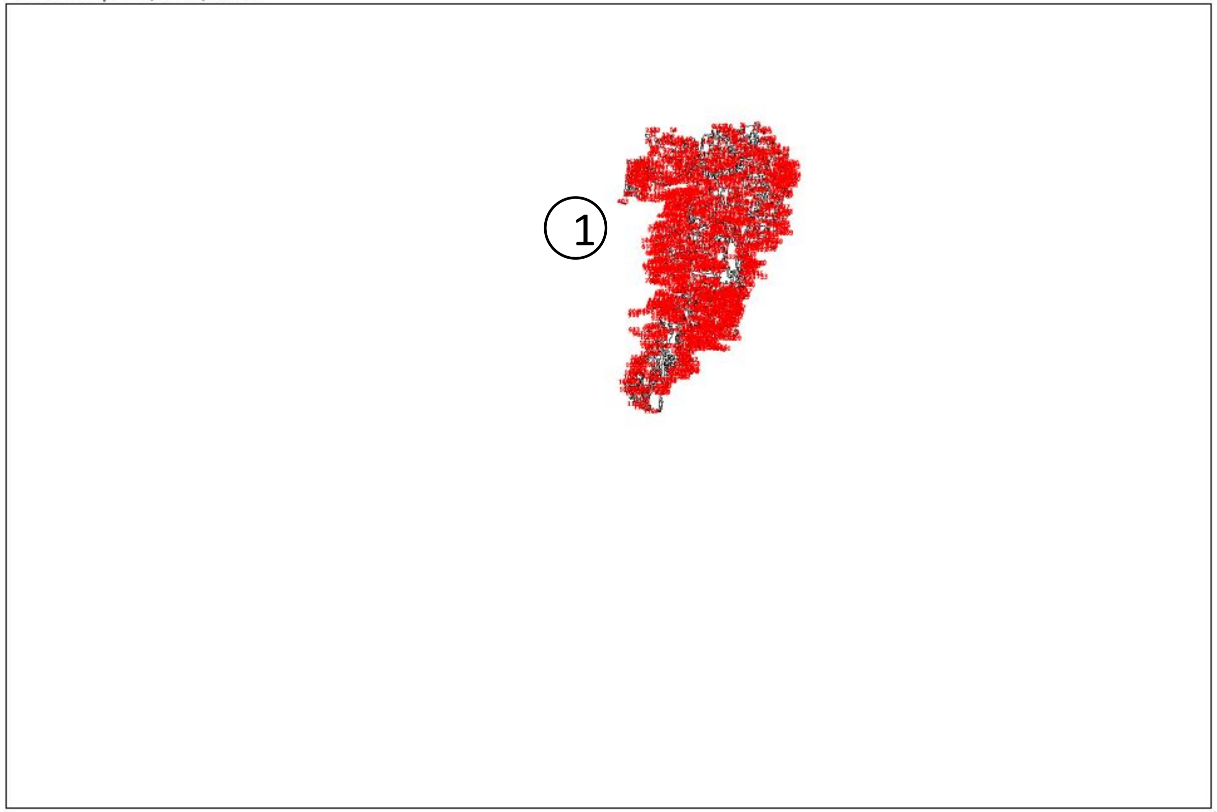


1920x1280 pixels; 8-bit; 2.3MB

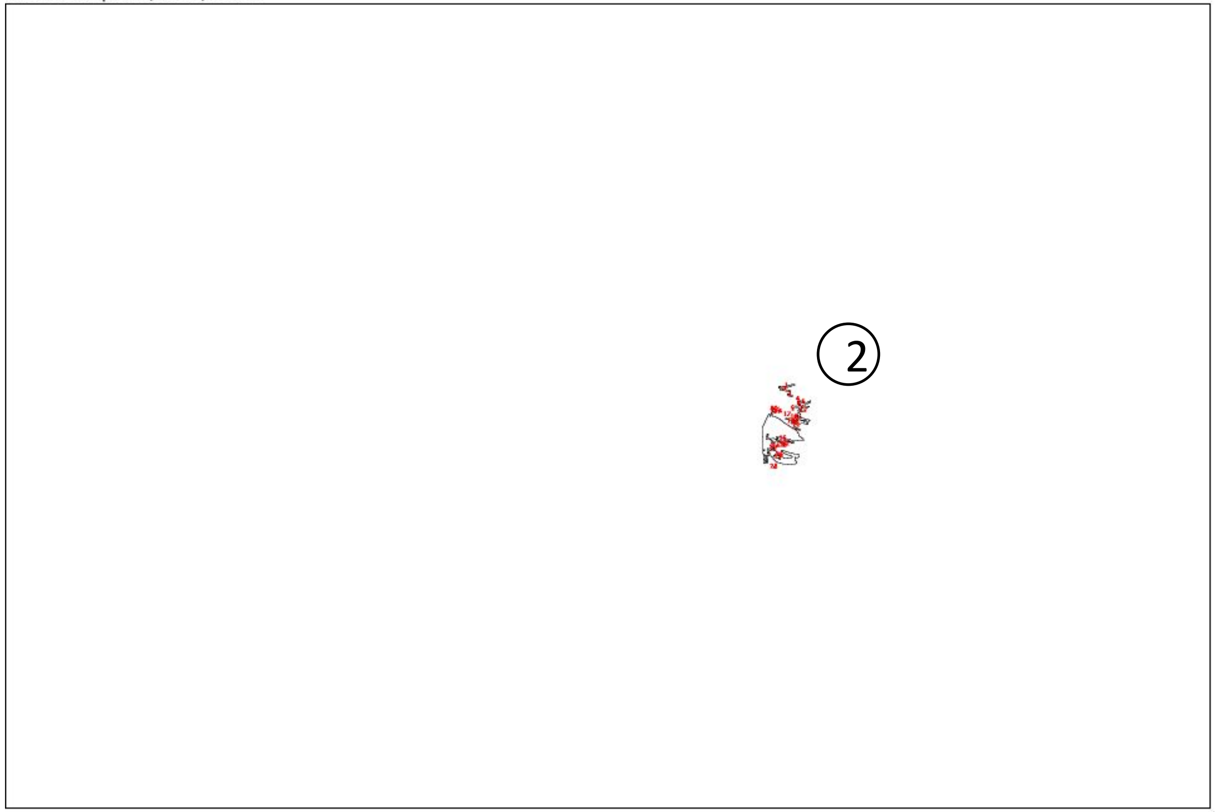


Result:

| | Slice | Count | Total Area | Average Size |
|---|---|---|---|---|
| ① | Scenario 1.1.jpg | 1104 | 24001 | 21.740 |
| ② | Scenario 2.1.jpg | 28 | 3047 | 108.821 |

*Count = number of areas*
*Total Area = total size of all areas*
*Average Size = average size of each area*

*Unit of area: PIXEL*

### Scenario 1.2 VS 2.2

*Size of the picture is 1920x1280 pixels*

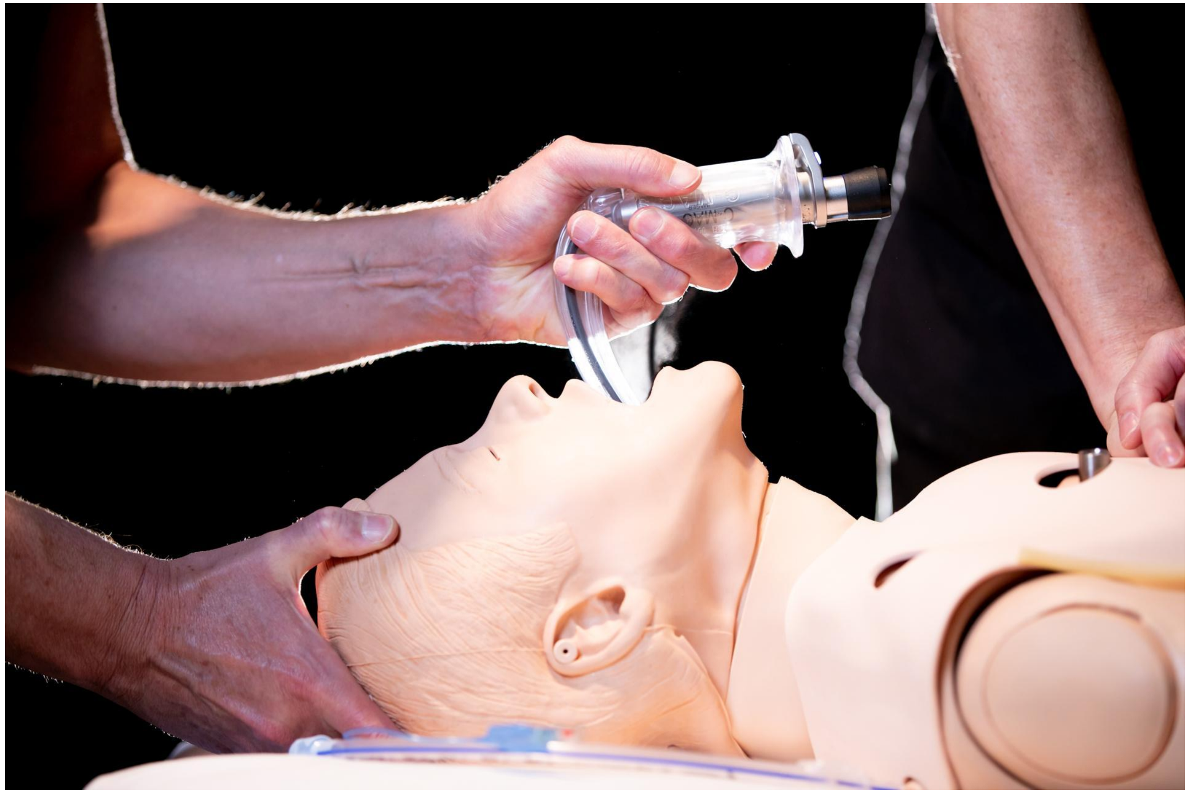

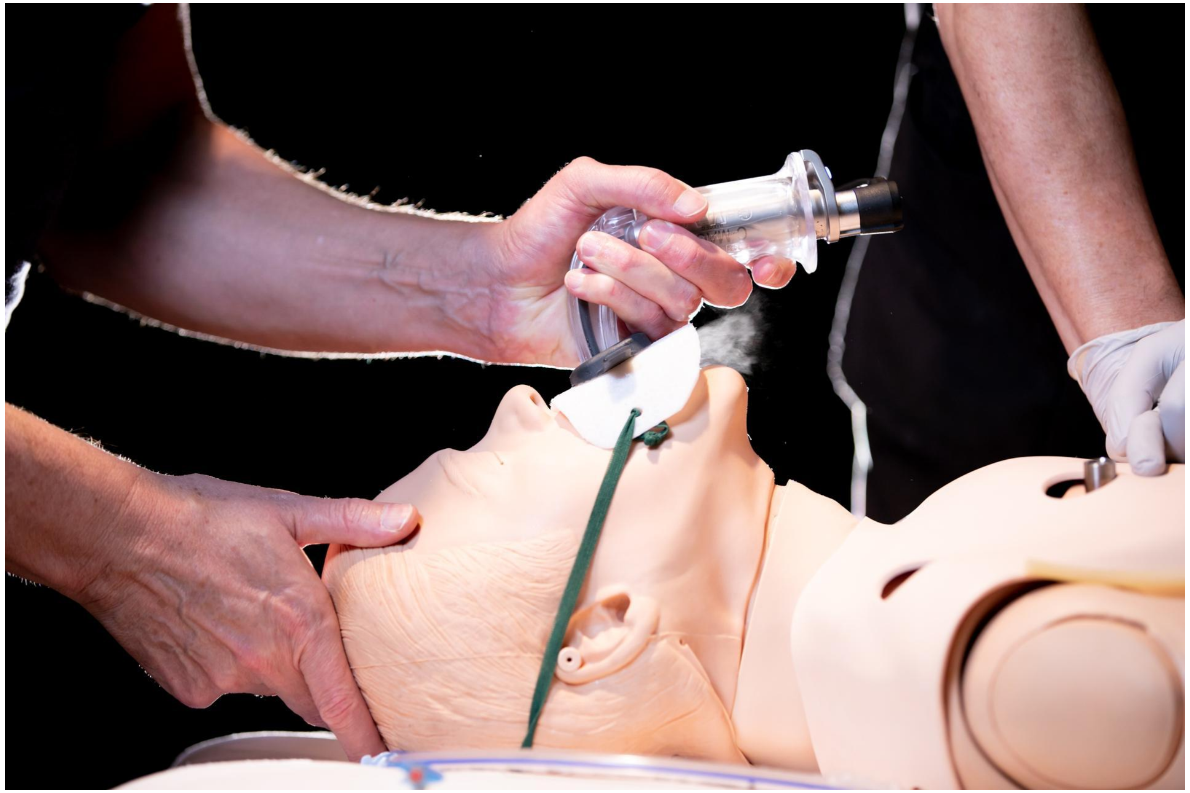

### Scenario 1.2 VS 2.2

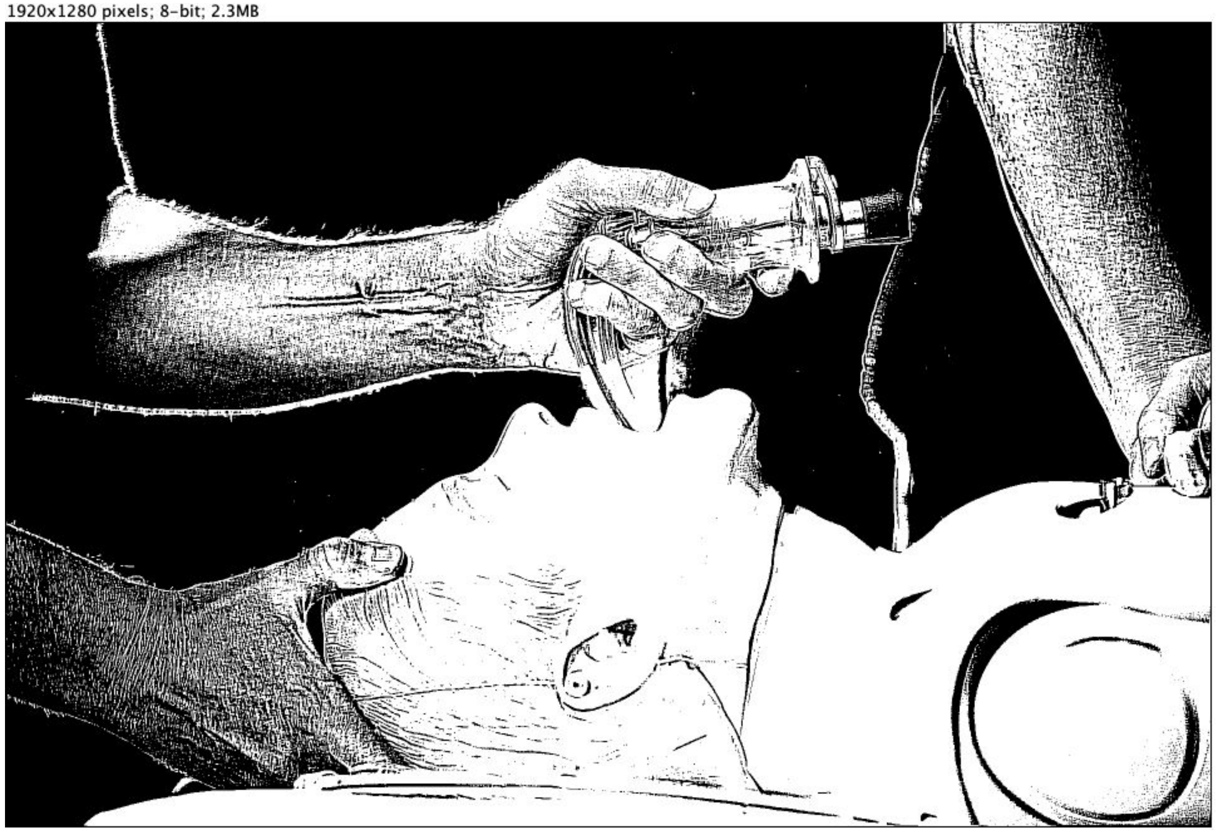


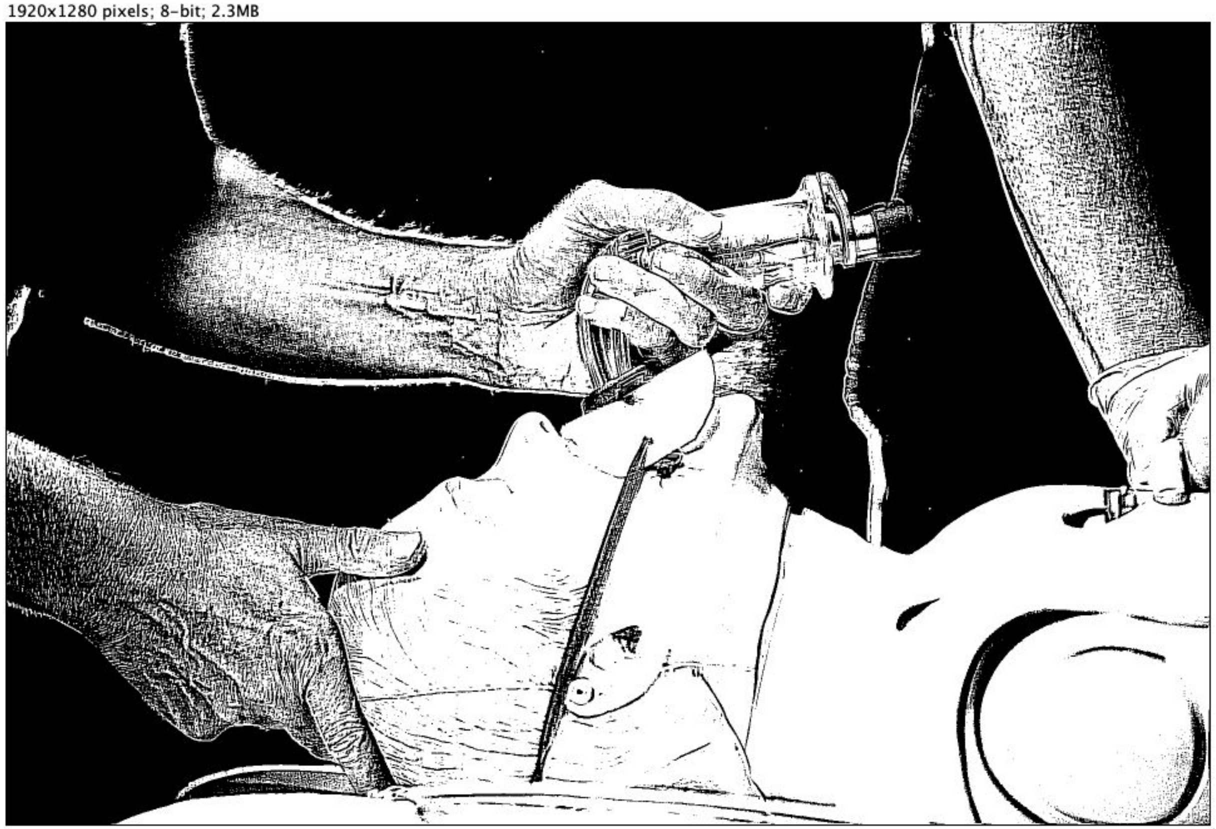

Scenario 1.2 VS 2.2

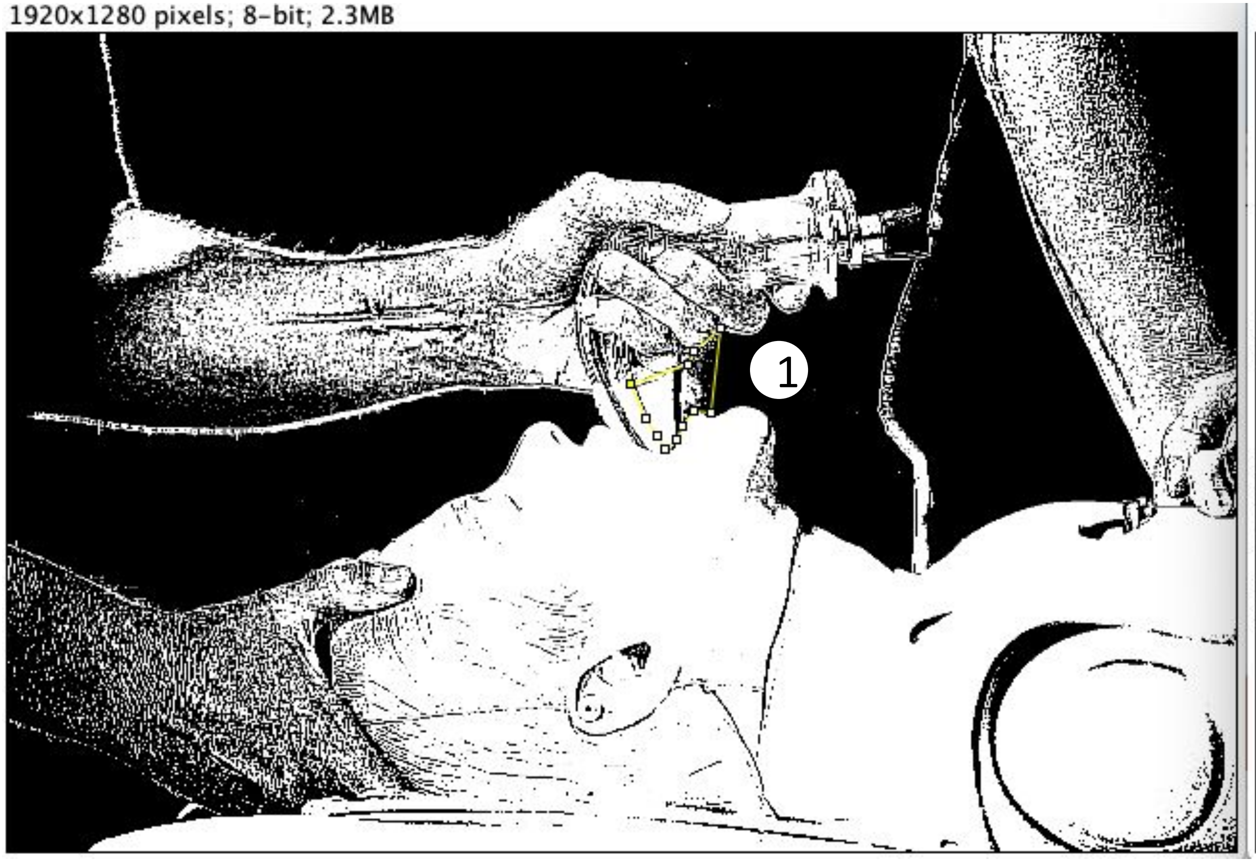


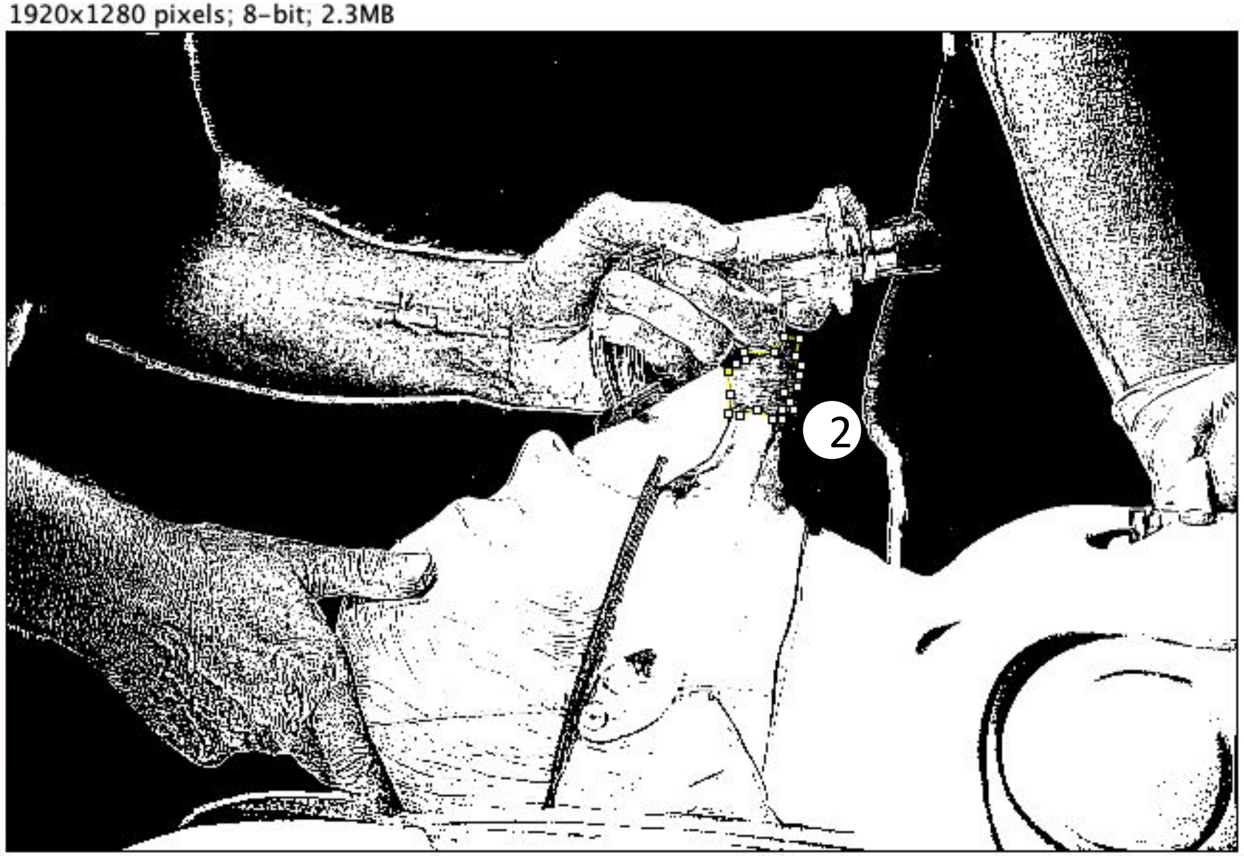

### Scenario 1.2 VS 2.2

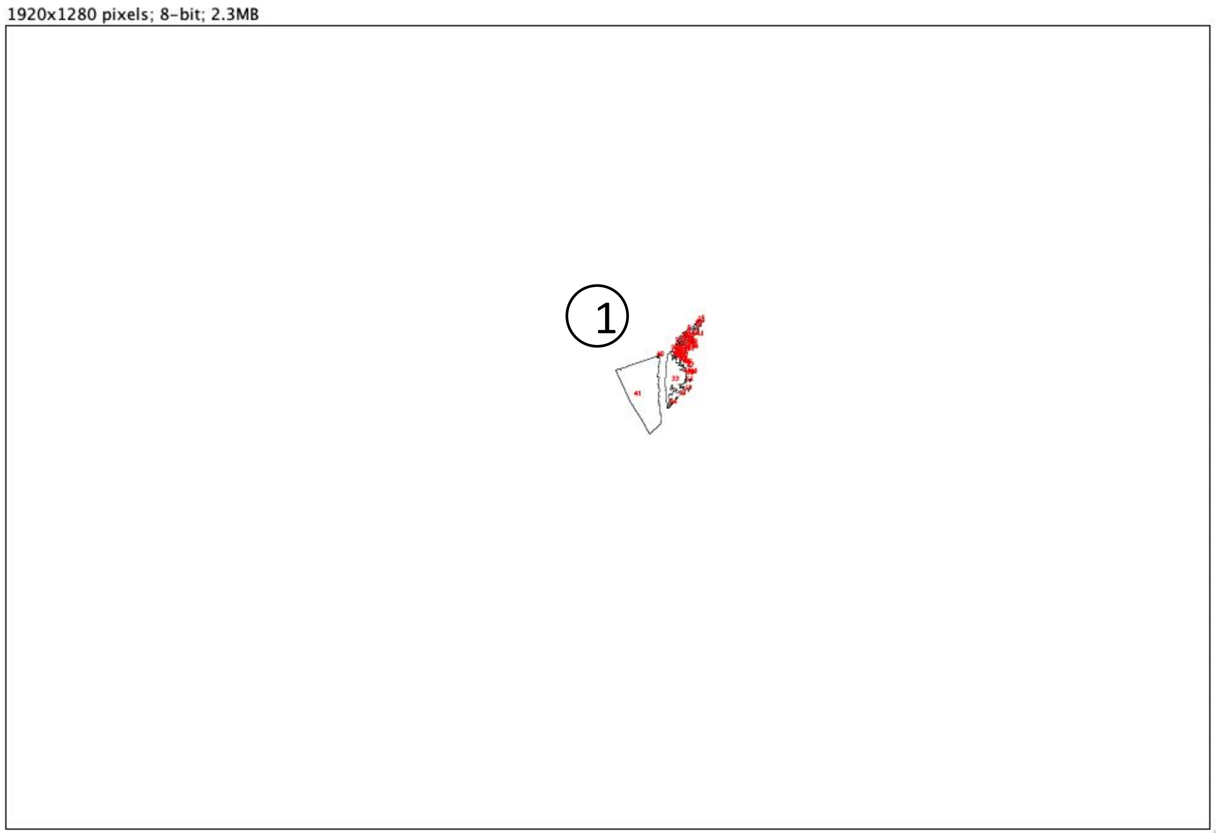


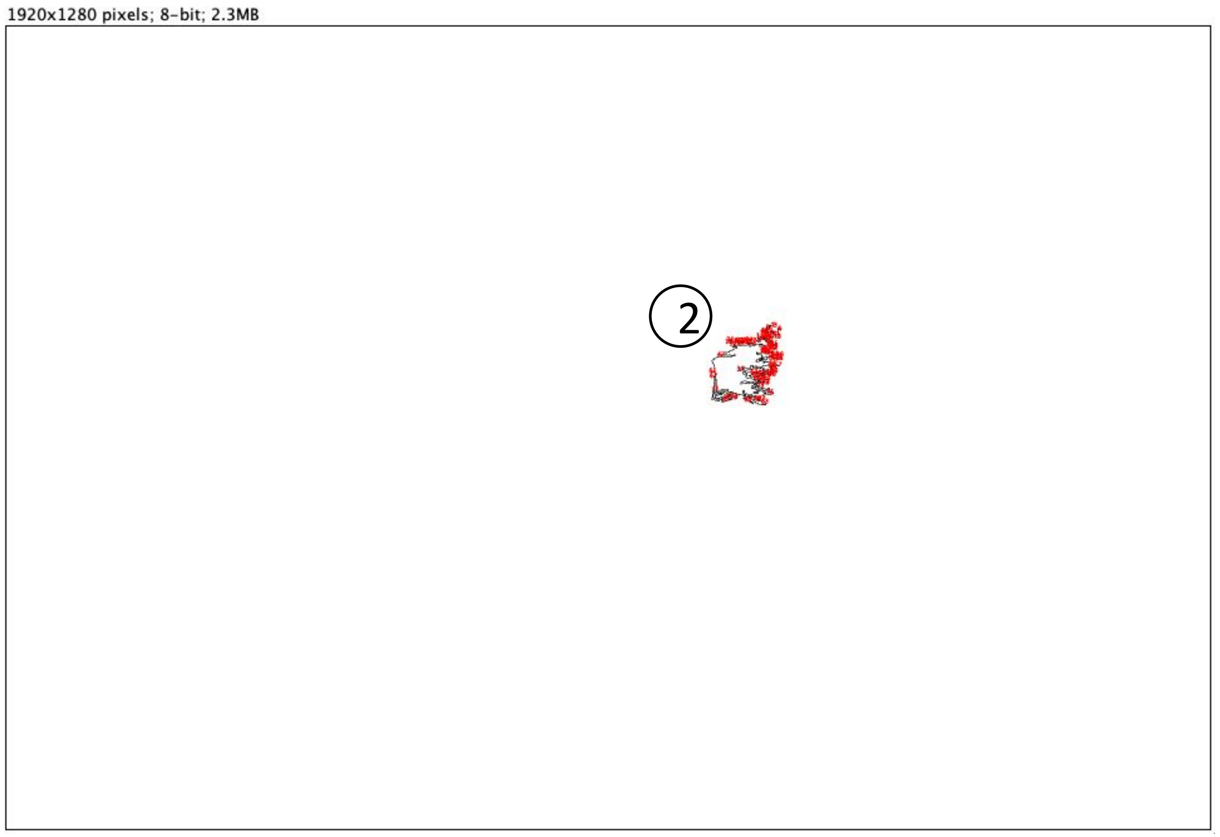


Result:

| | Slice | Count | Total Area | Average Size |
|---|---|---|---|---|
| ① | Scenario 1.2.jpg | 54 | 7324 | 135.630 |
| ② | Scenario 2.2.jpg | 82 | 850 | 10.366 |

***(Unit of area: PIXEL)***

*Count = number of areas*
*Total Area = total size of all areas*
*Average Size = average size of each area*

*Unit of area: PIXEL*

### Scenario 1.3 VS 2.3

*Size of the picture is 1920x1280 pixels*

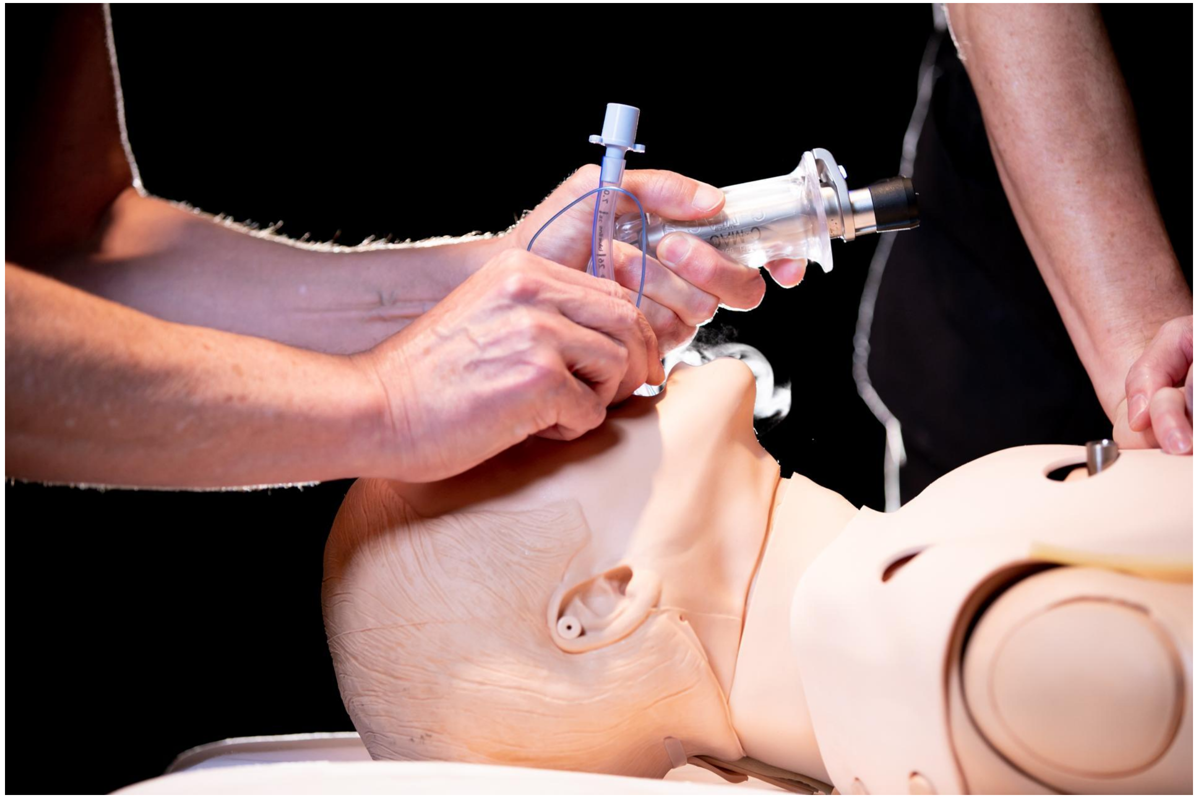

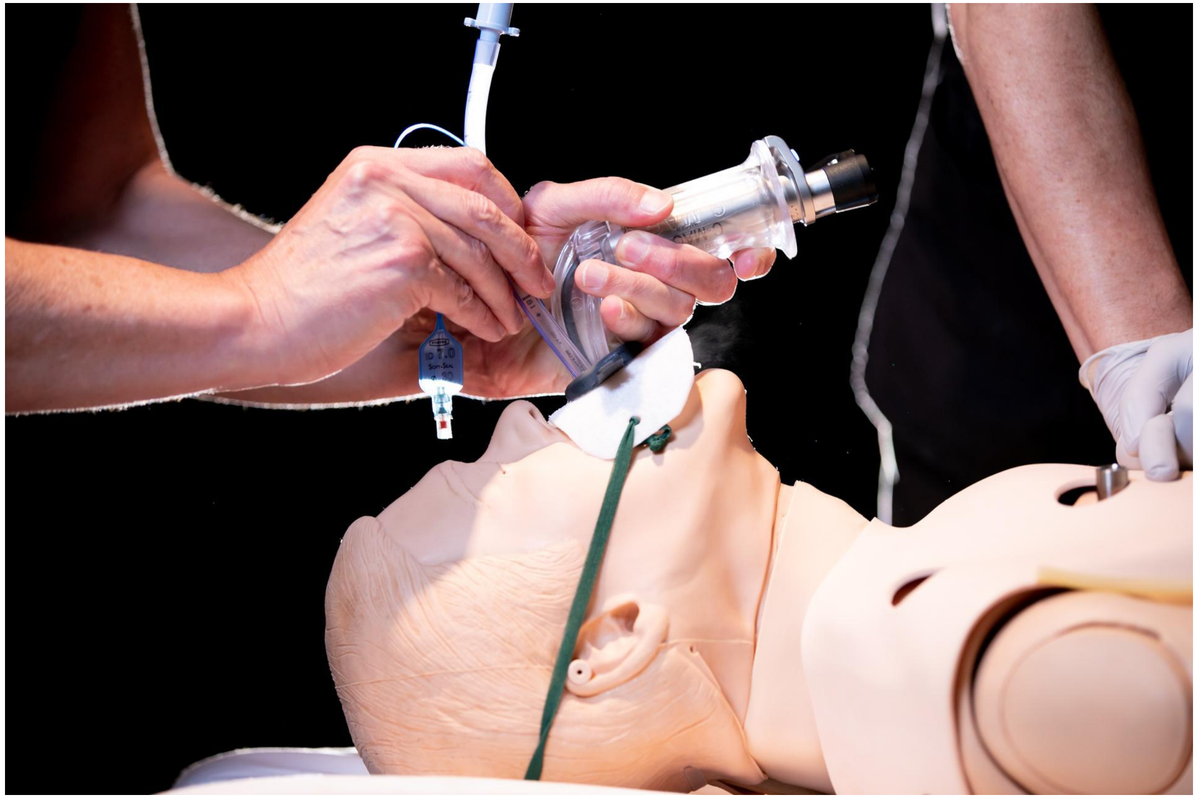

Scenario 1.3 VS 2.3

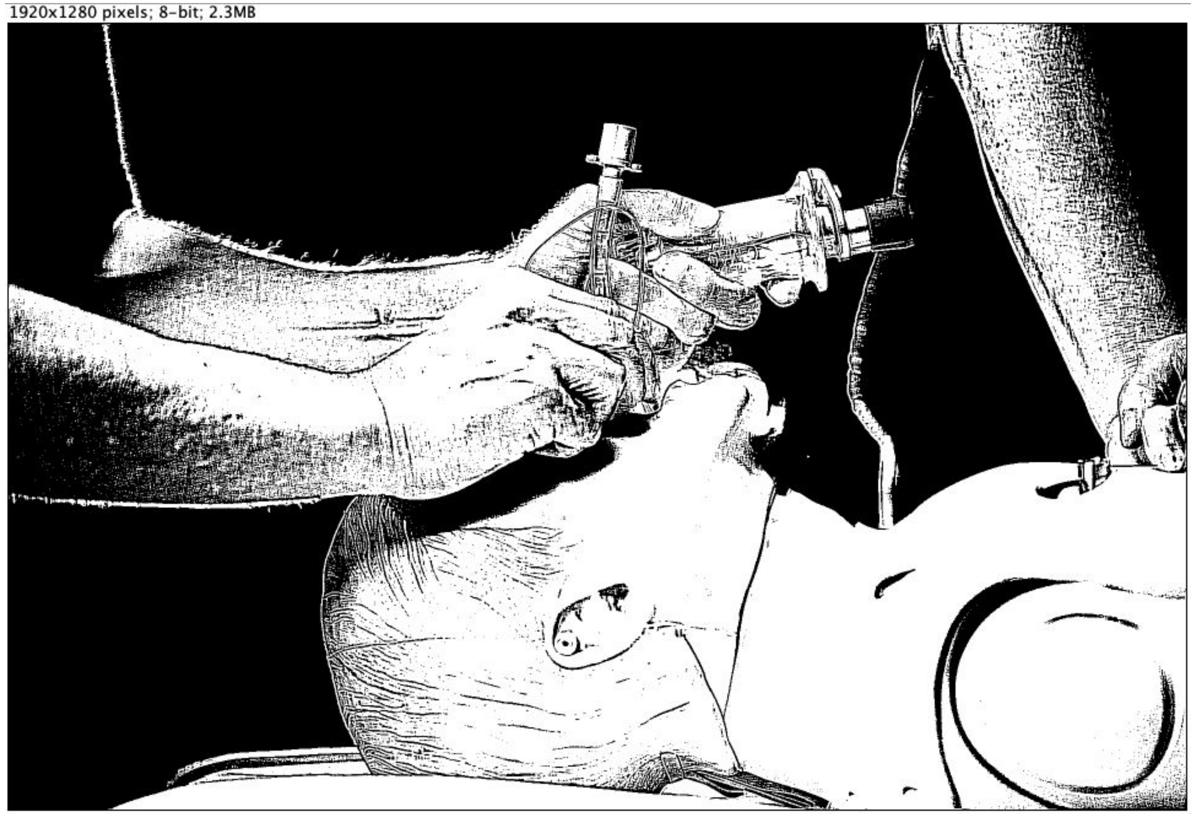


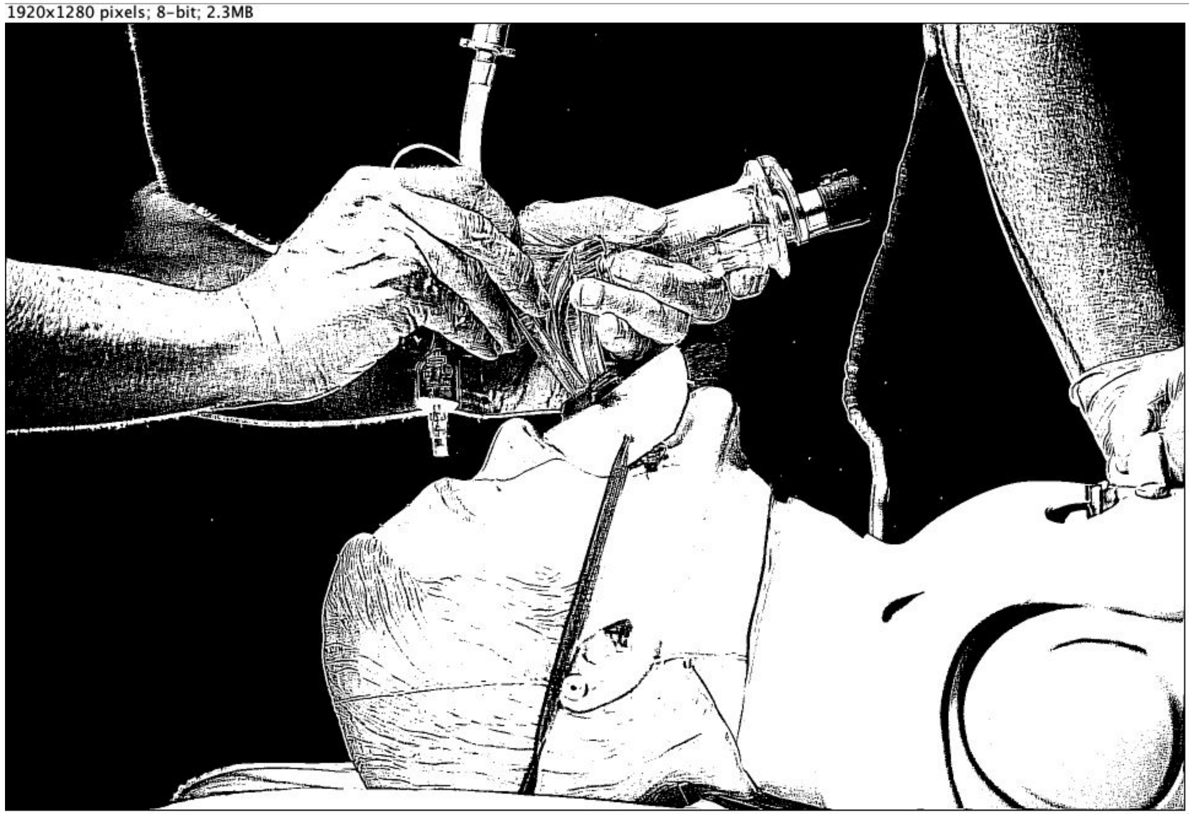

### Scenario 1.3 VS 2.3

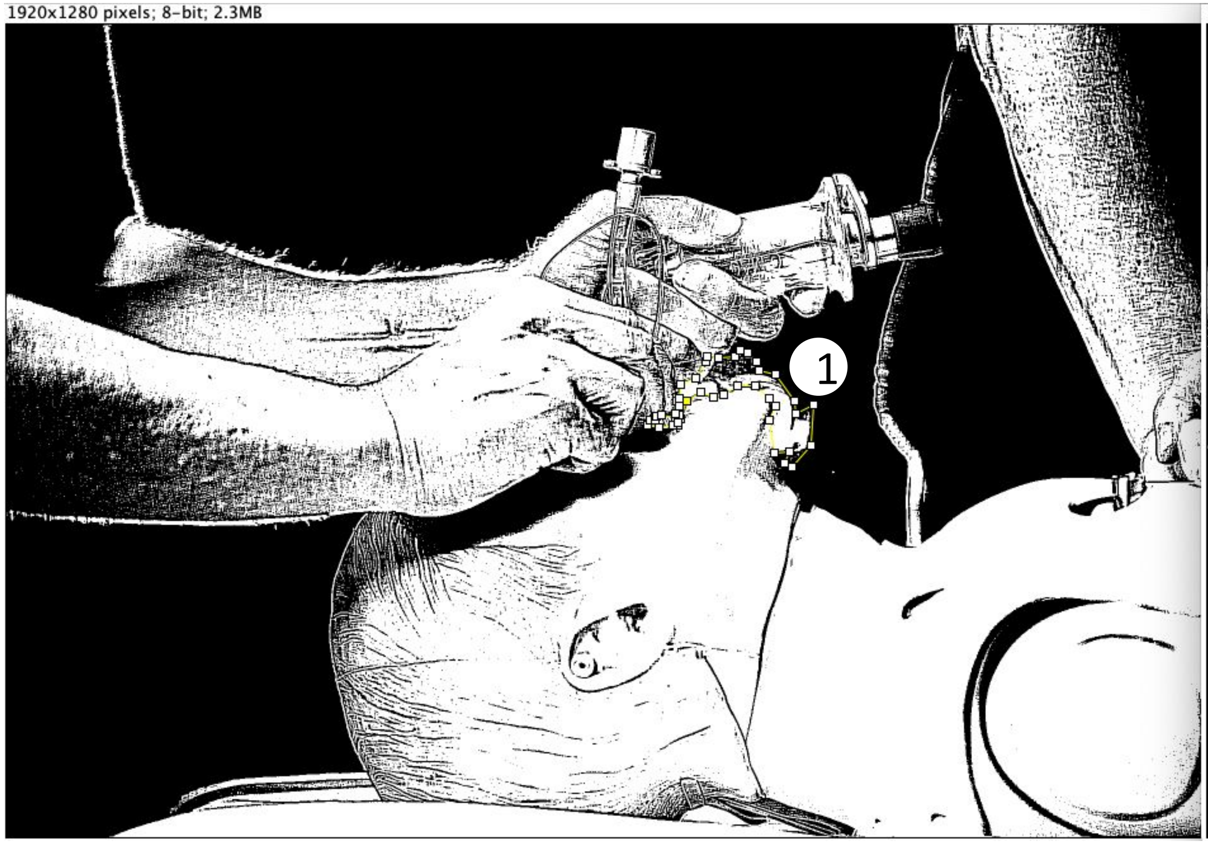


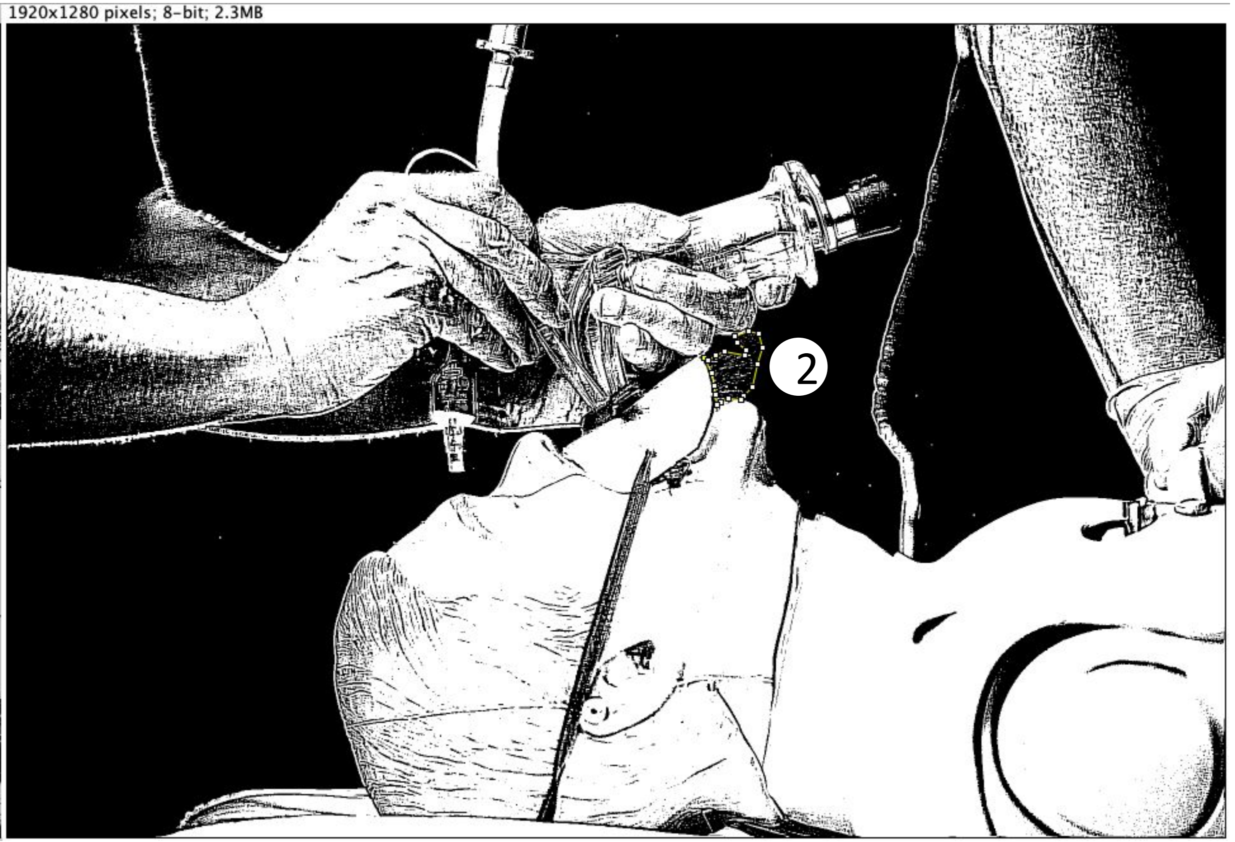

### Scenario 1.3 VS 2.3

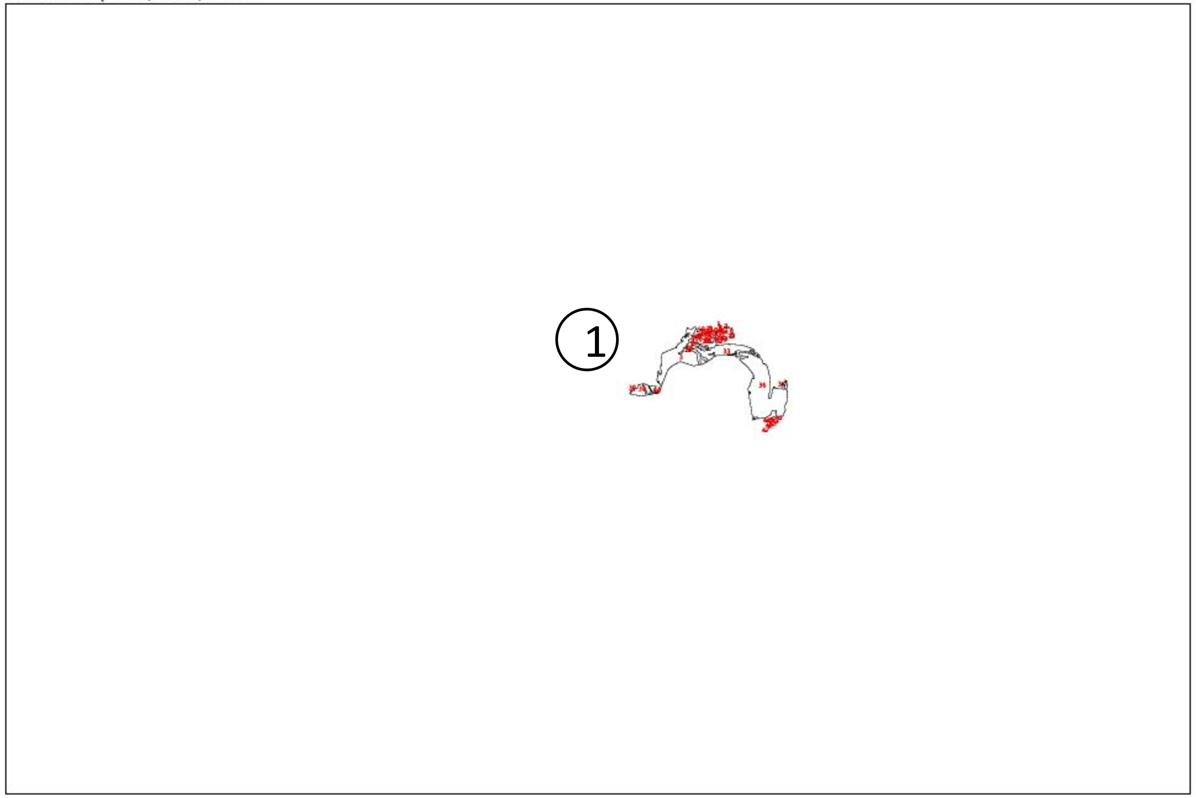


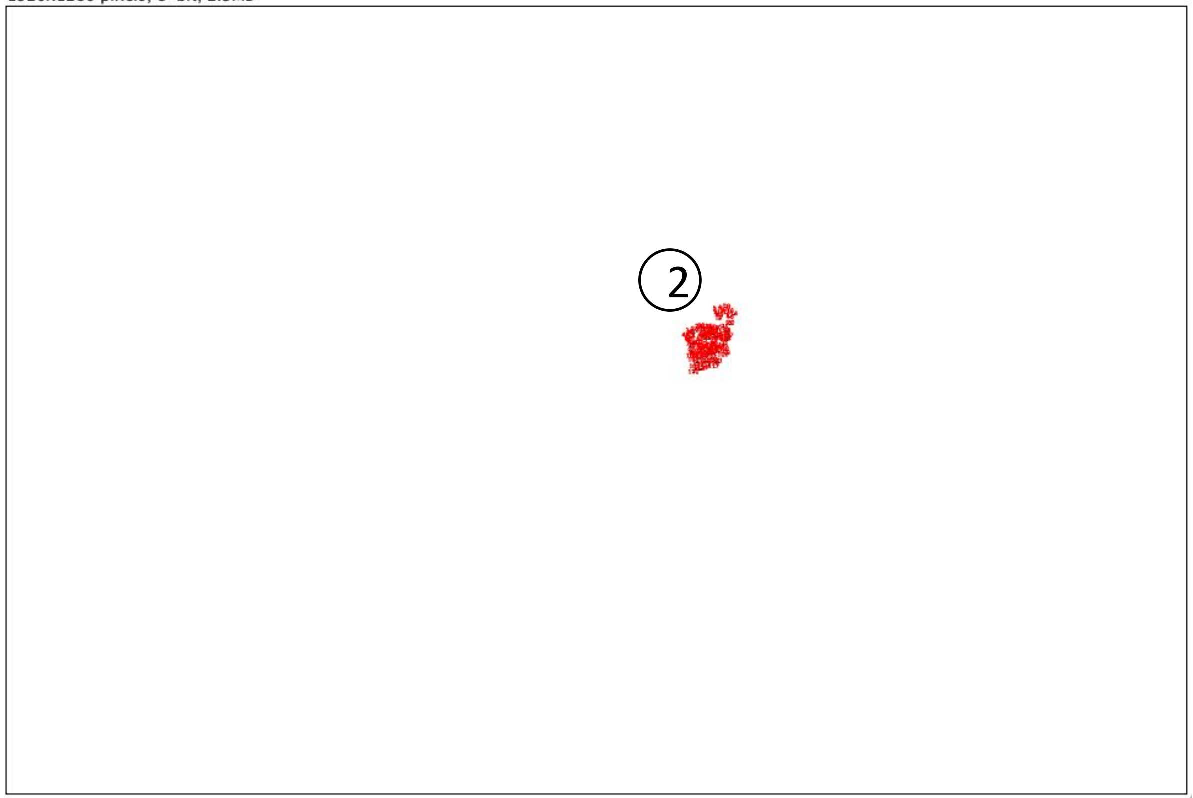


Result:

| | Slice | Count | Total Area | Average Size |
|---|---|---|---|---|
| ① | Scenario 1.3.jpg | 47 | 8694 | 184.979 |
| ② | Scenario 2.3.jpg | 120 | 655 | 5.458 |

***(Unit of area: PIXEL)***

*Count = number of areas*
*Total Area = total size of all areas*
*Average Size = average size of each area*

*Unit of area: PIXEL*

### Scenario 3.1 VS 4.1

*Size of the picture is 1920x1280 pixels*

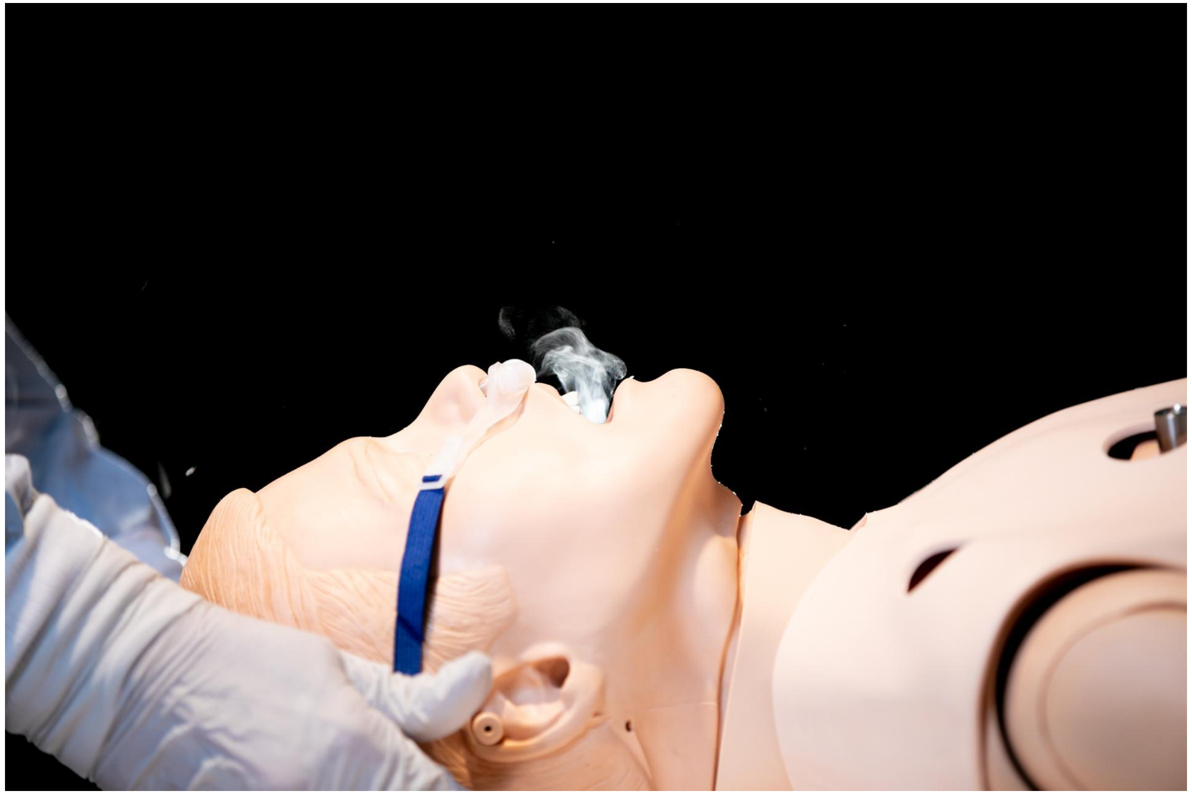

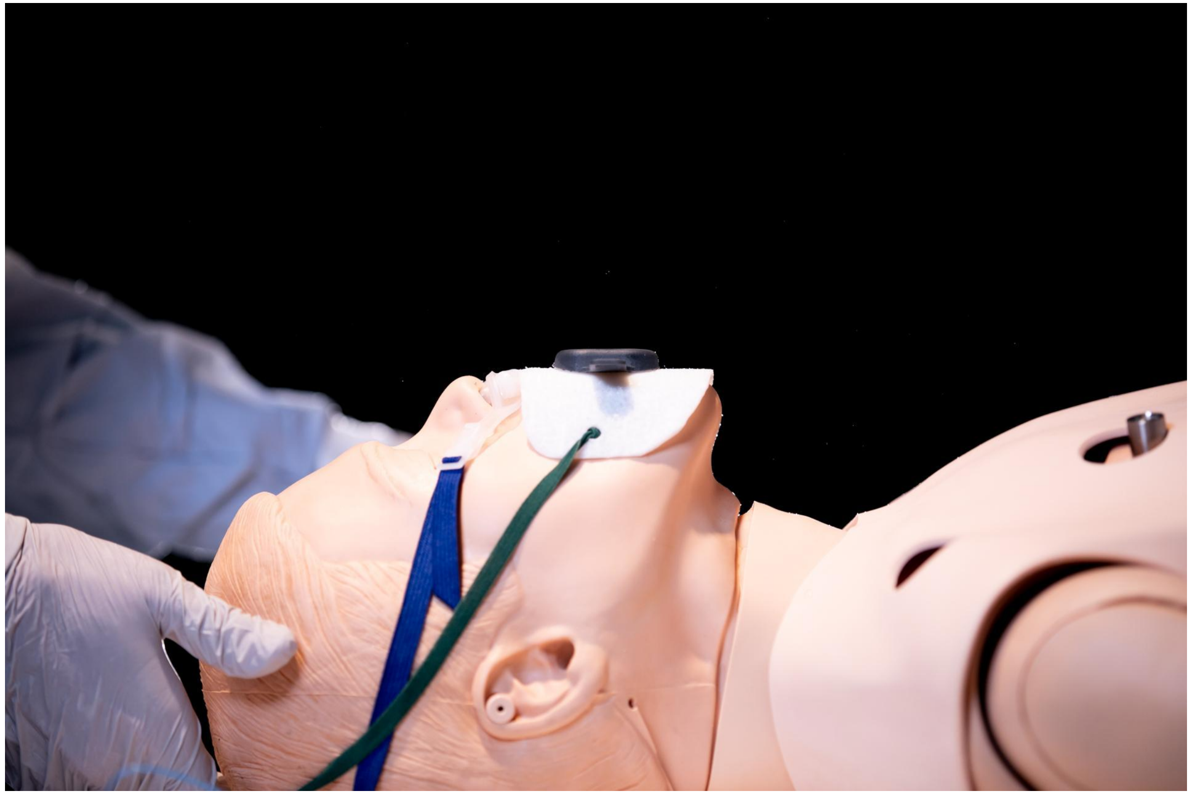

### Scenario 3.1 VS 4.1

1920x1280 pixels; 8-bit; 2.3MB

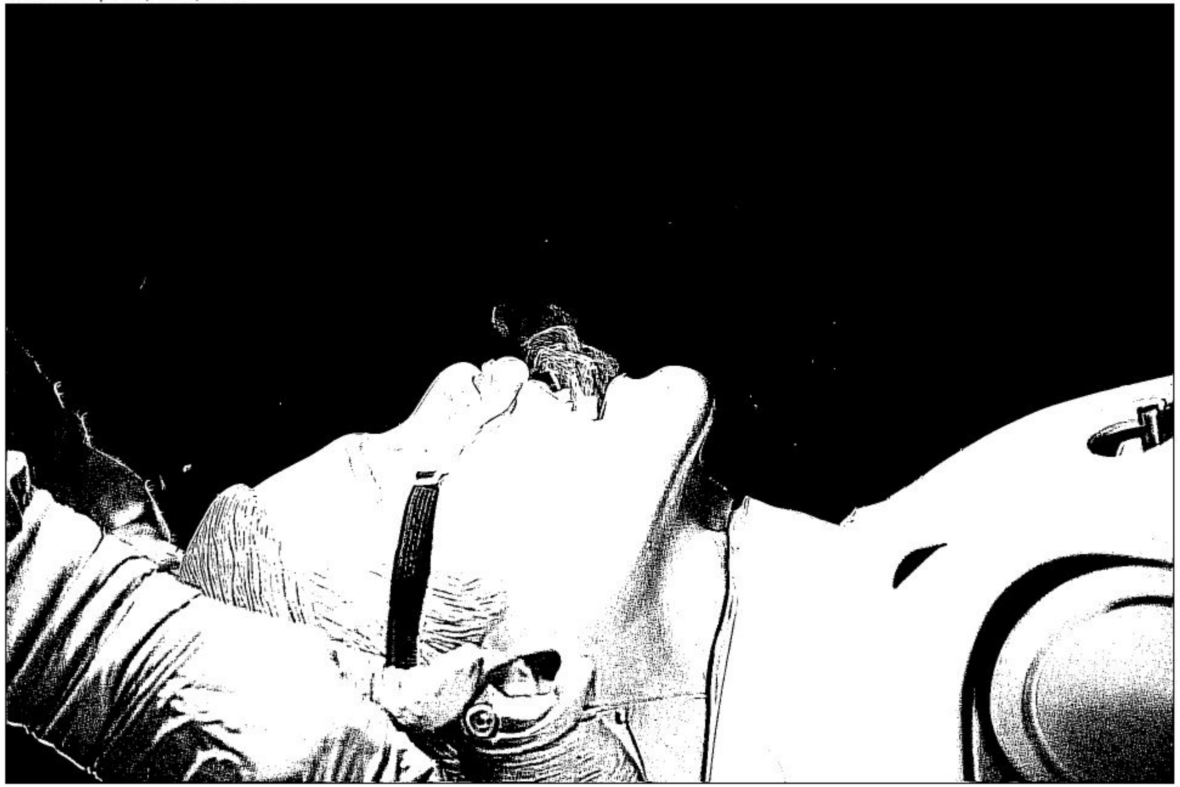

1920x1280 pixels; 8-bit; 2.3MB

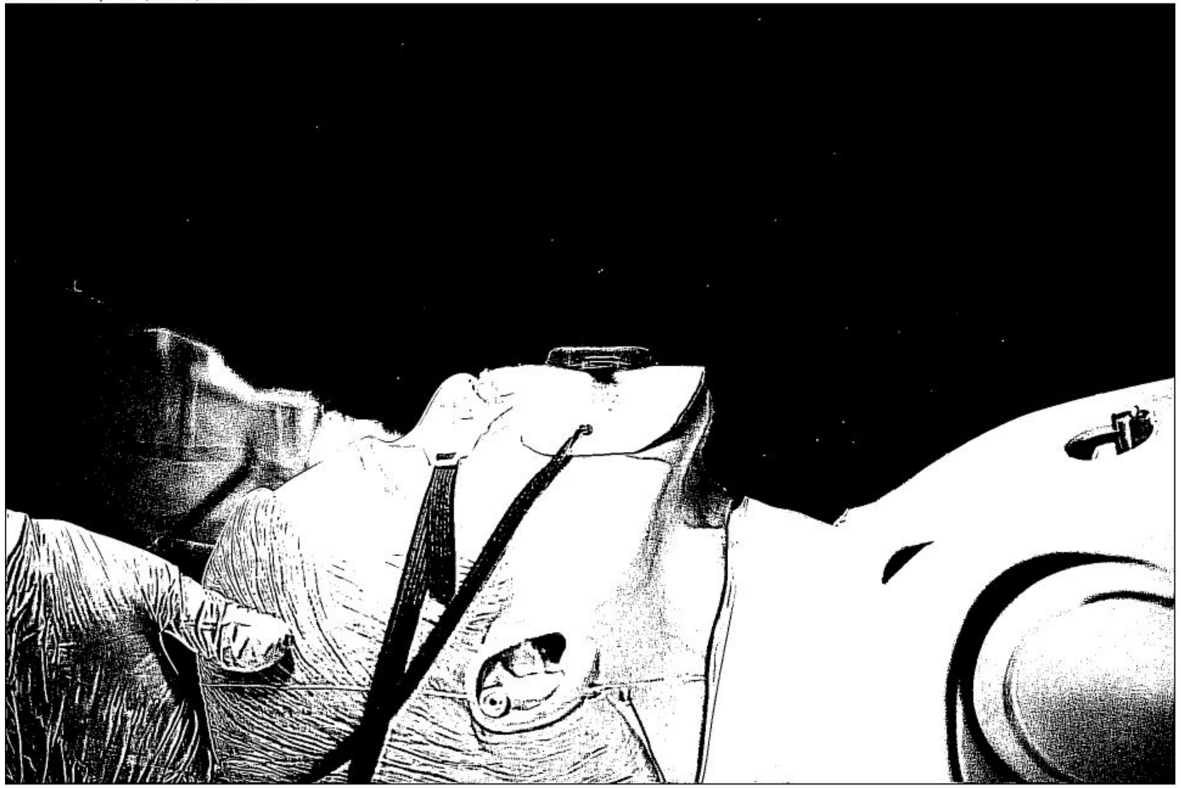

Scenario 3.1 VS 4.1

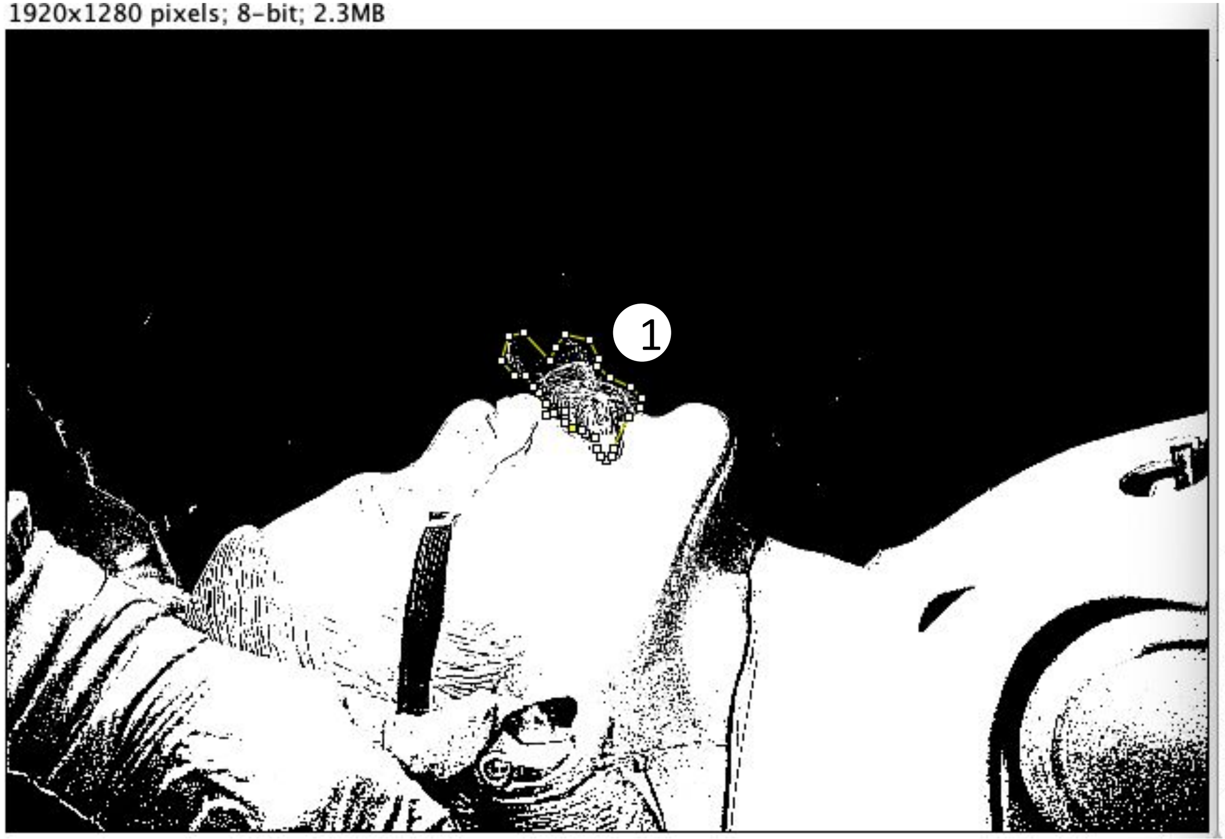


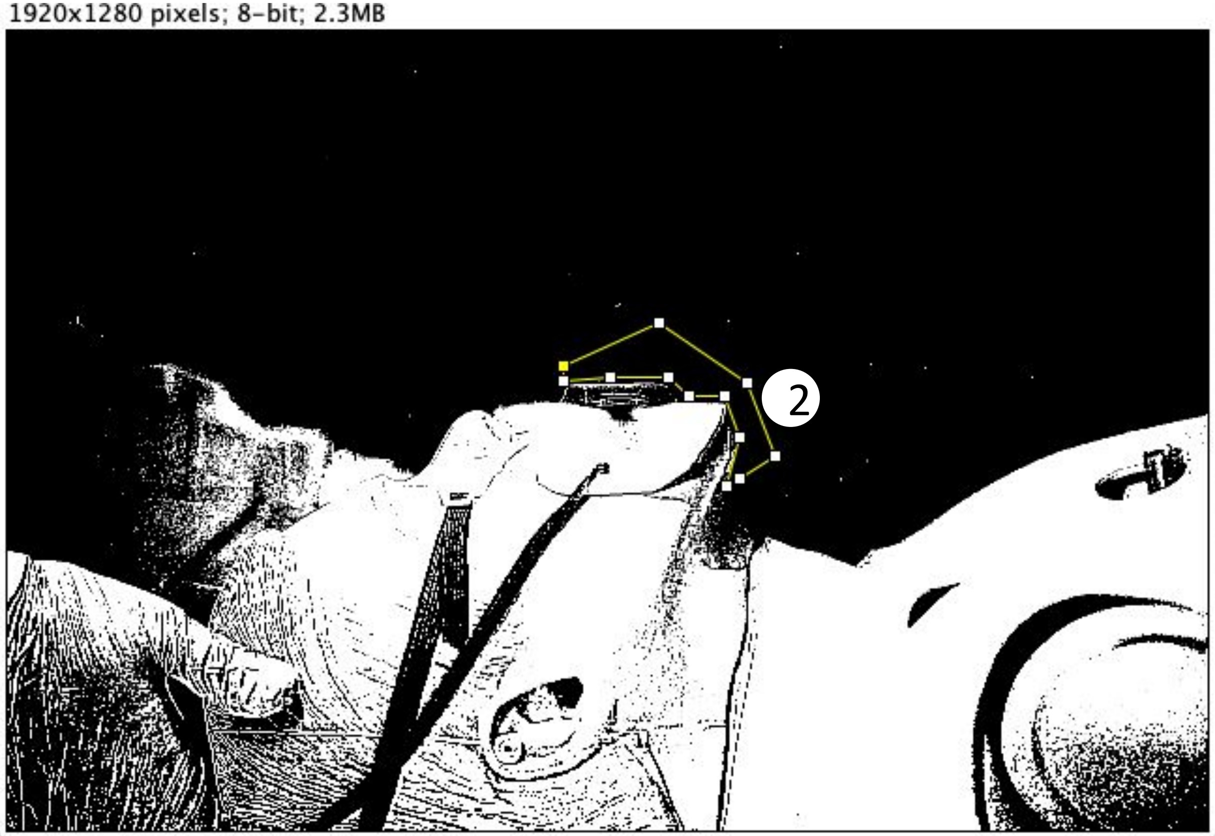

### Scenario 3.1 VS 4.1

1920x1280 pixels; 8-bit; 2.3MB

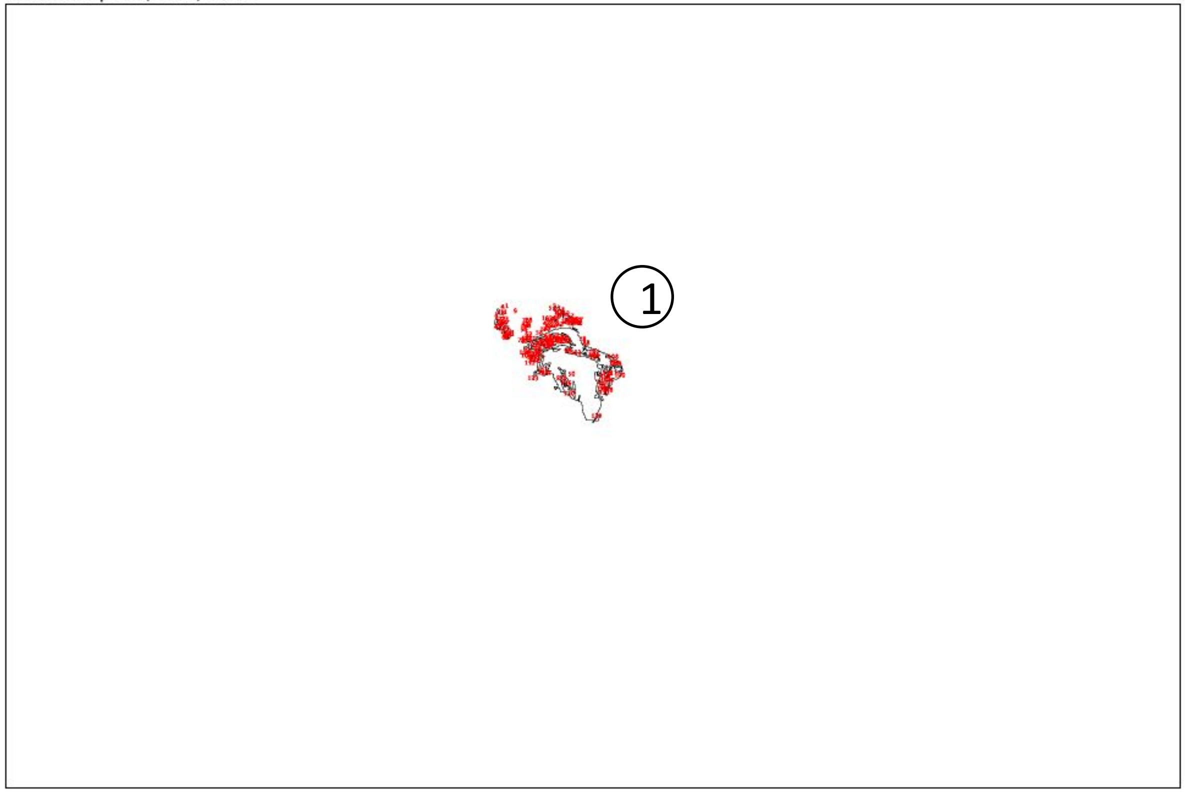


1920x1280 pixels; 8-bit; 2.3MB

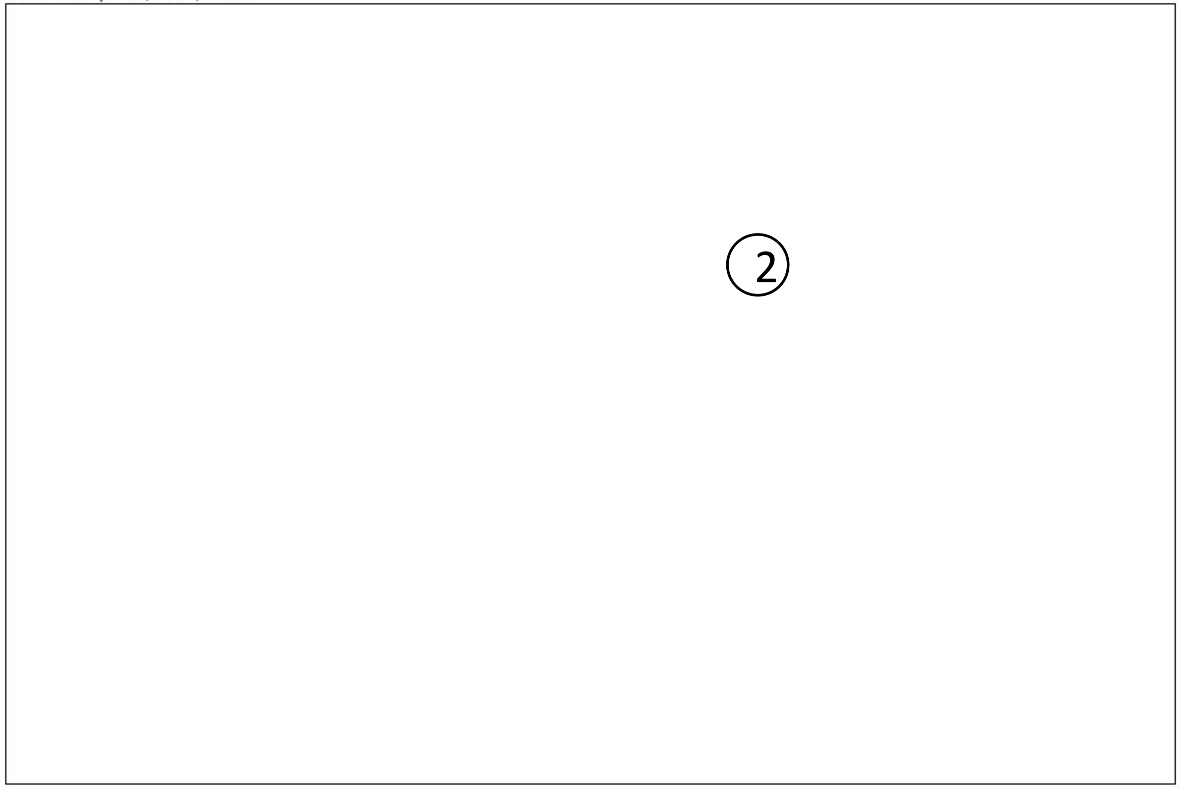


Result:

| | Slice | Count | Total Area | Average Size |
|---|---|---|---|---|
| ① | Scenario 3.1.jpg | 132 | 9308 | 70.515 |
| ② | Scenario 4.1.jpg | 0 | 0 | NaN |

***(Unit of area: PIXEL)***

*Count = number of areas*
*Total Area = total size of all areas*
*Average Size = average size of each area*

*Unit of area: PIXEL*

### Scenario 3.2 VS 4.2

*Size of the picture is 1920x1280 pixels*

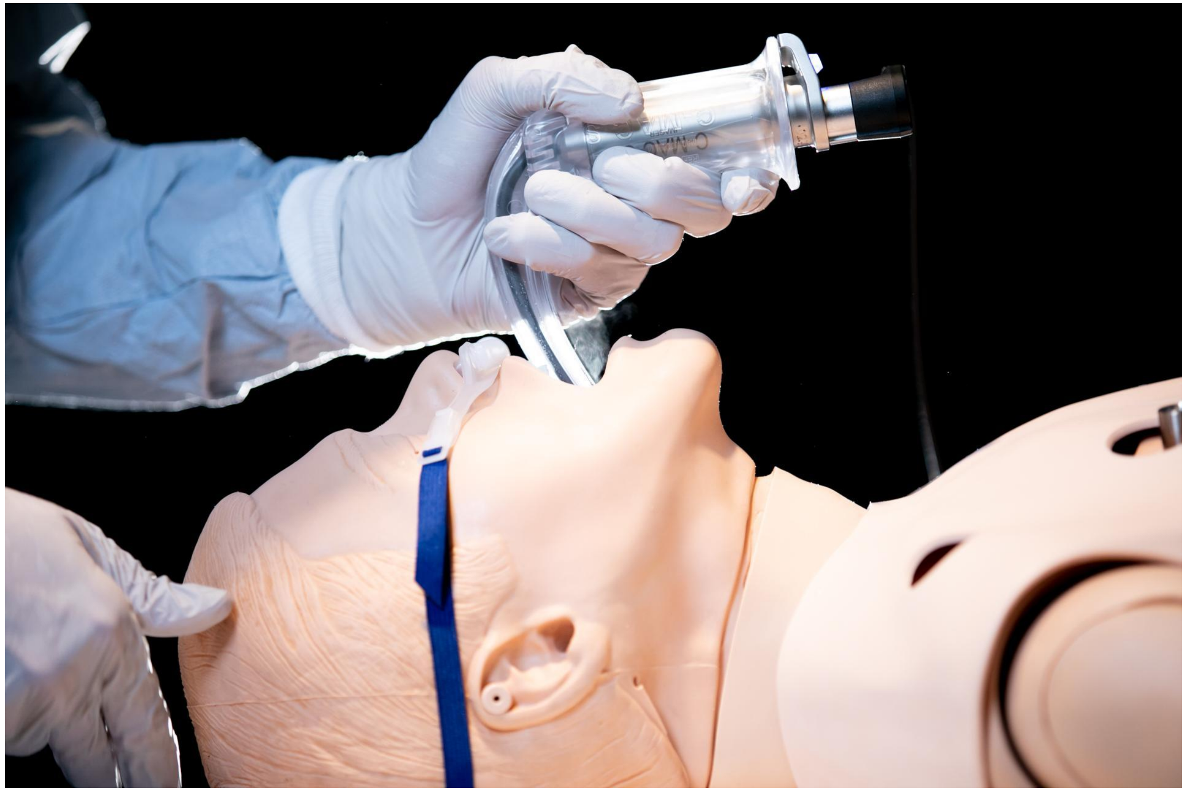

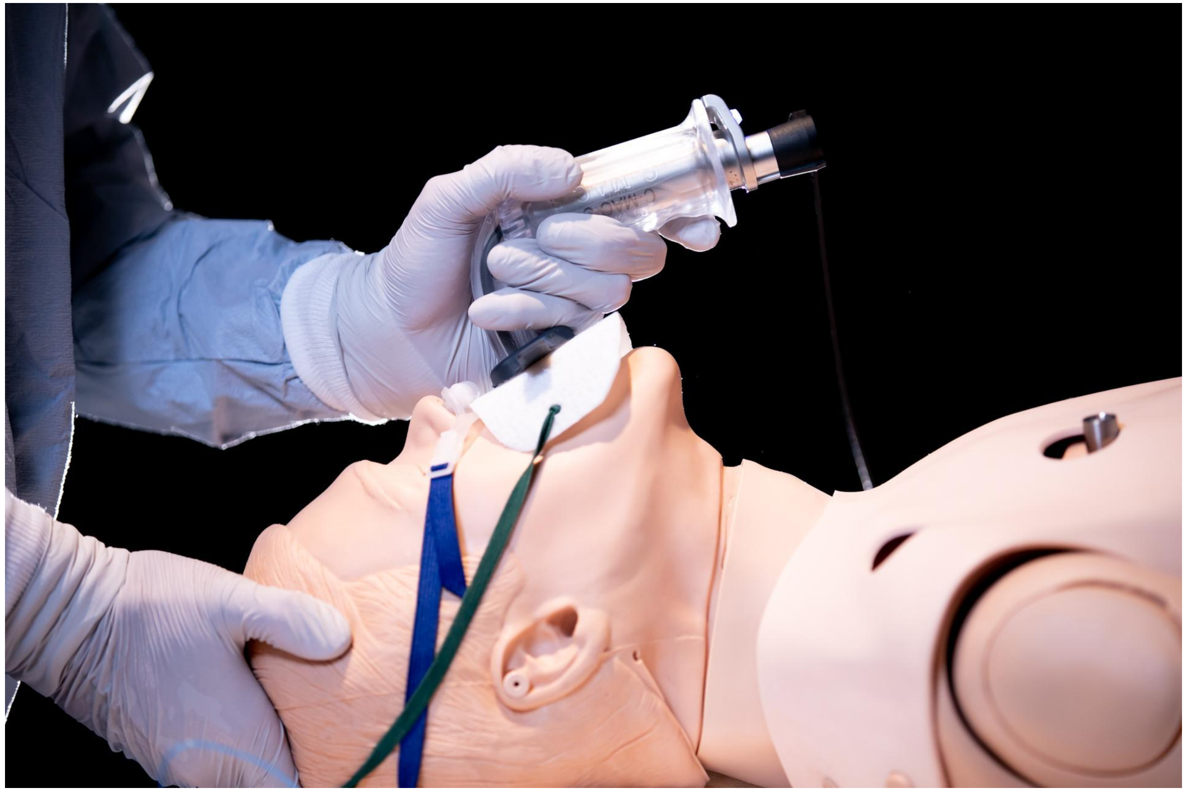

### Scenario 3.2 VS 4.2

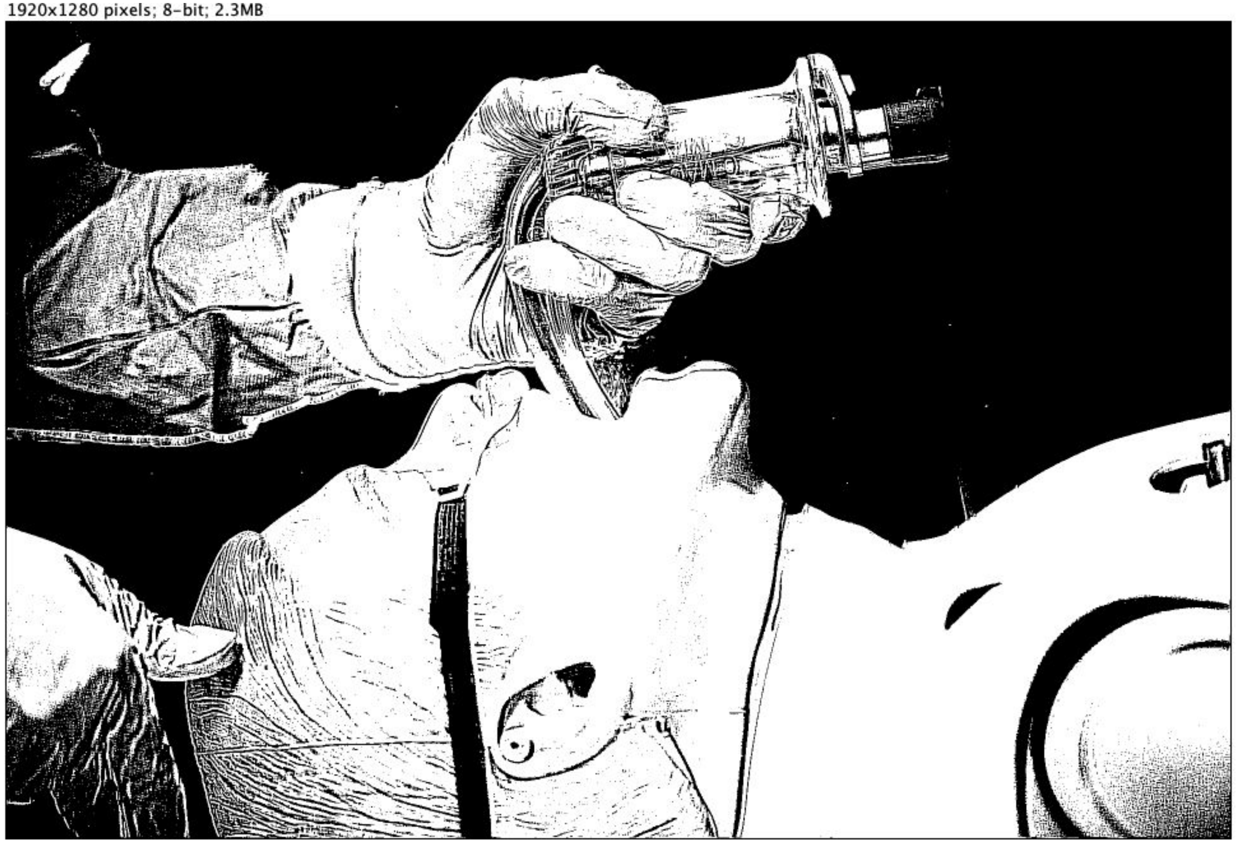


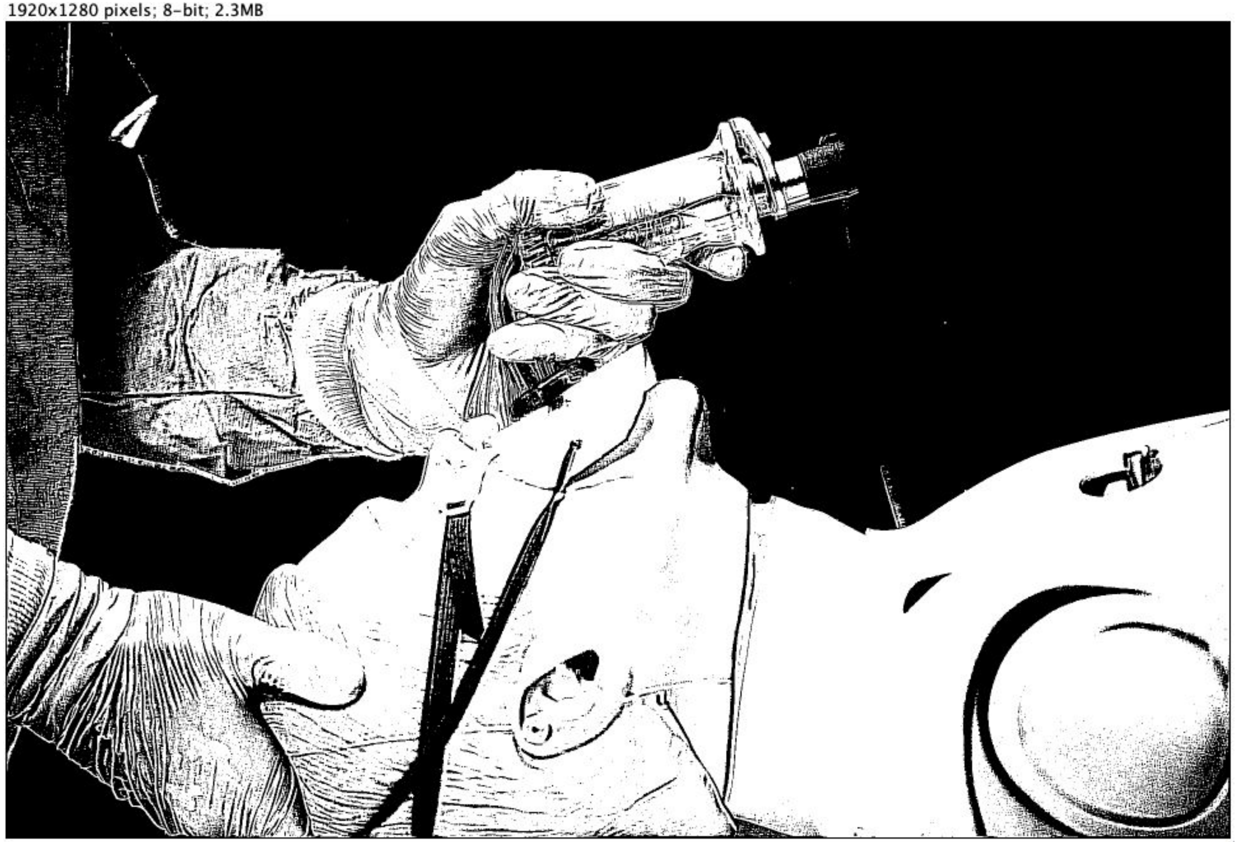

Scenario 3.2 VS 4.2

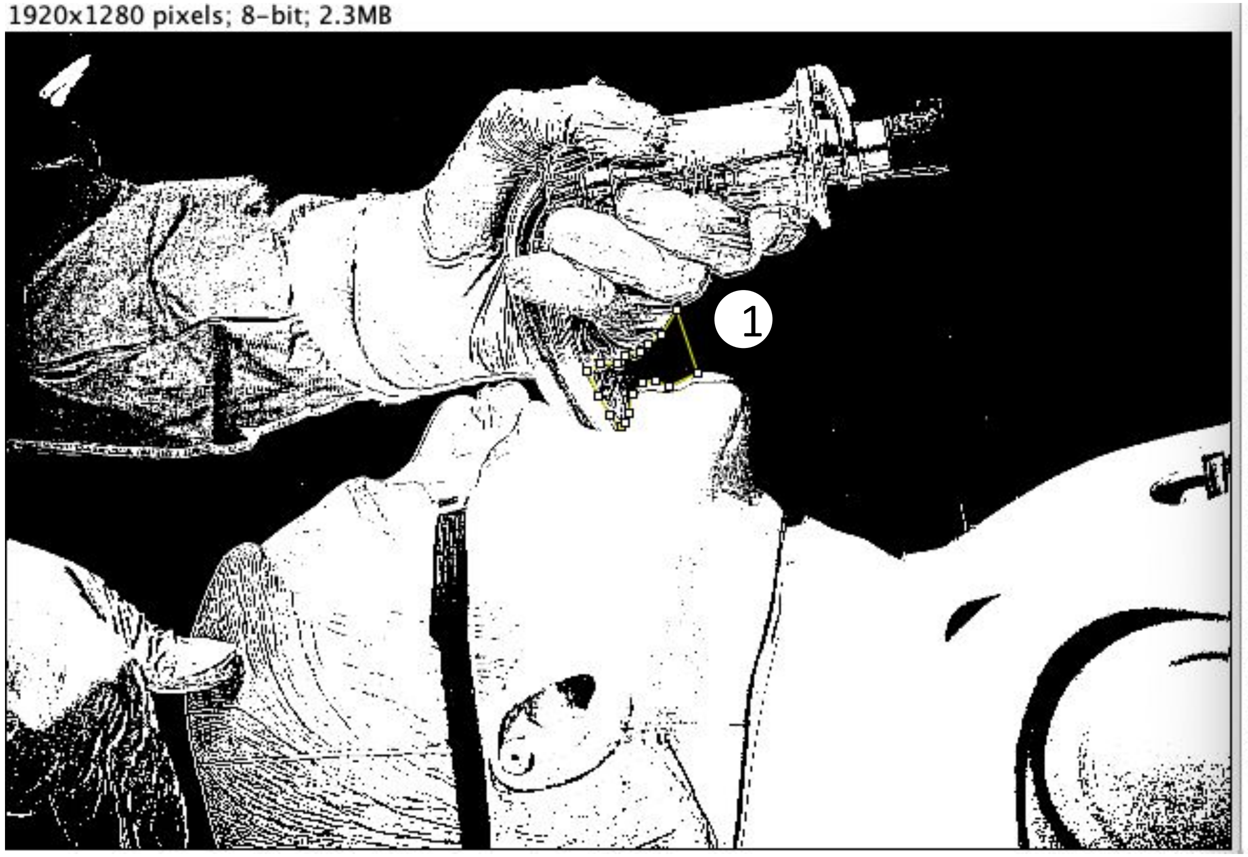


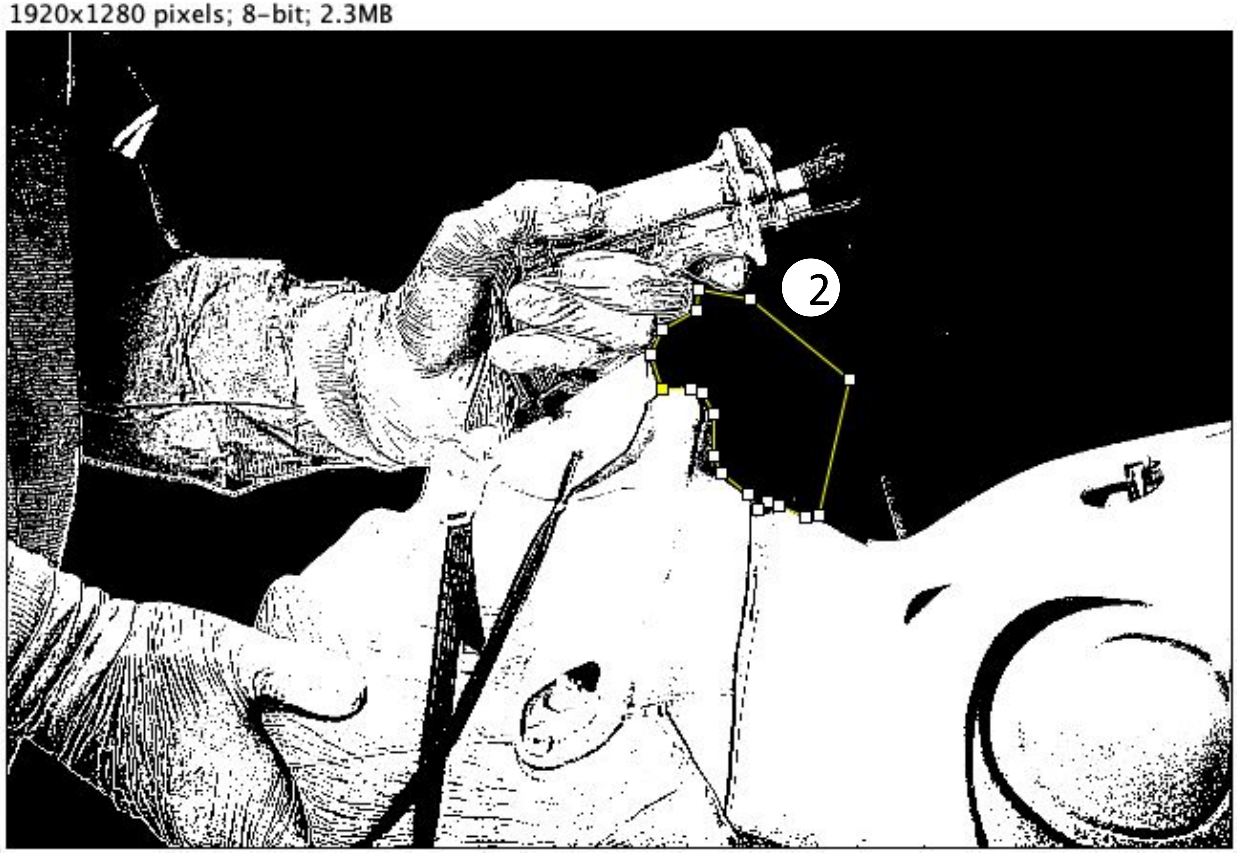

### Scenario 3.2 VS 4.2

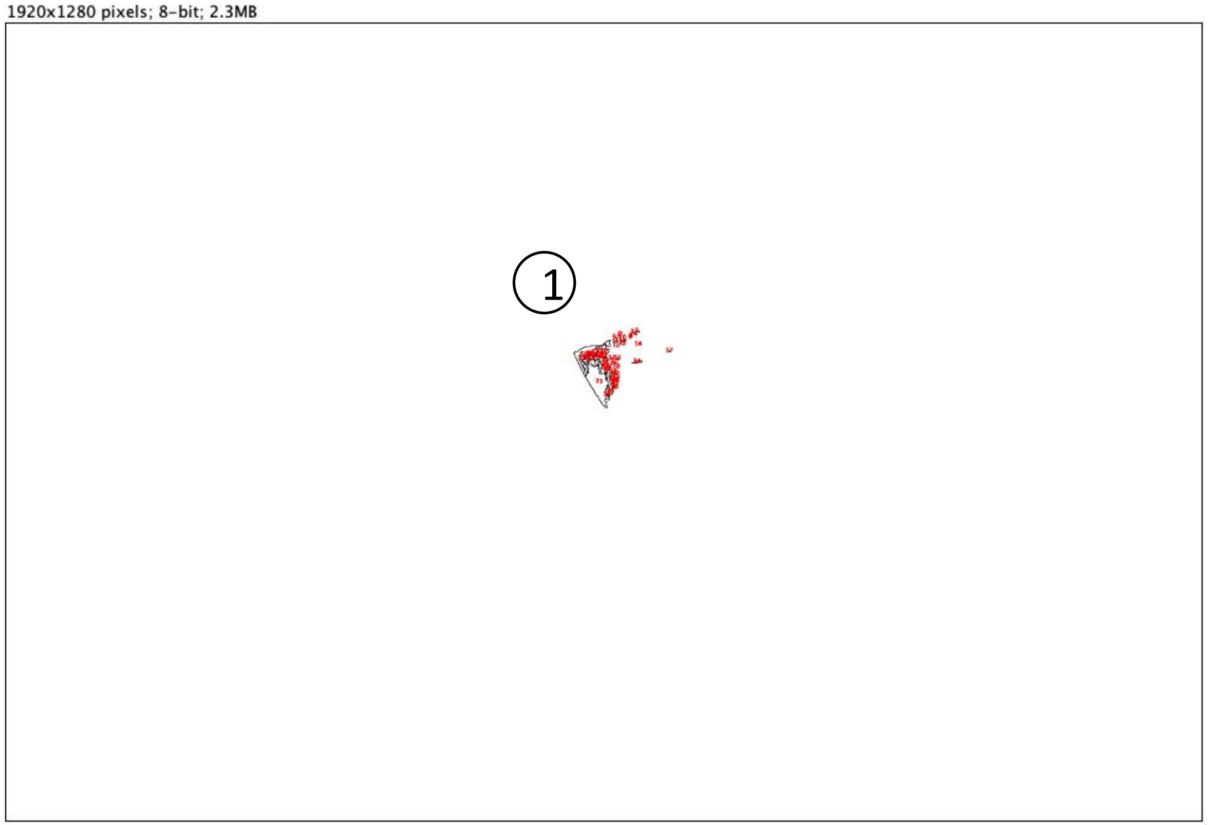


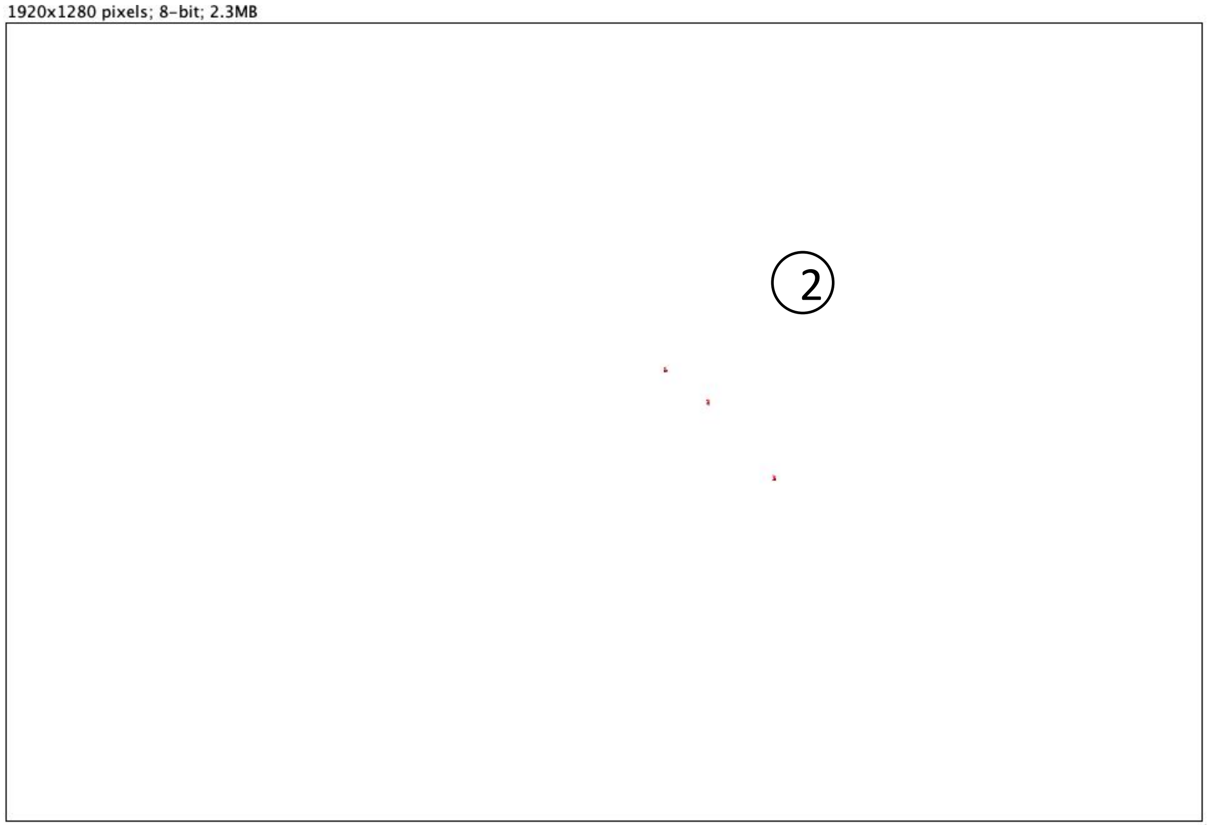


Result:

| | Slice | Count | Total Area | Average Size |
|---|---|---|---|---|
| 1 | Scenario 3.2.jpg | 50 | 2618 | 52.360 |
| 2 | Scenario 4.2.jpg | 3 | 23 | 7.667 |

***(Unit of area: PIXEL)***

*Count = number of areas*
*Total Area = total size of all areas*
*Average Size = average size of each area*

*Unit of area: PIXEL*

### Scenario 3.3 VS 4.3

*Size of the picture is 1920x1280 pixels*

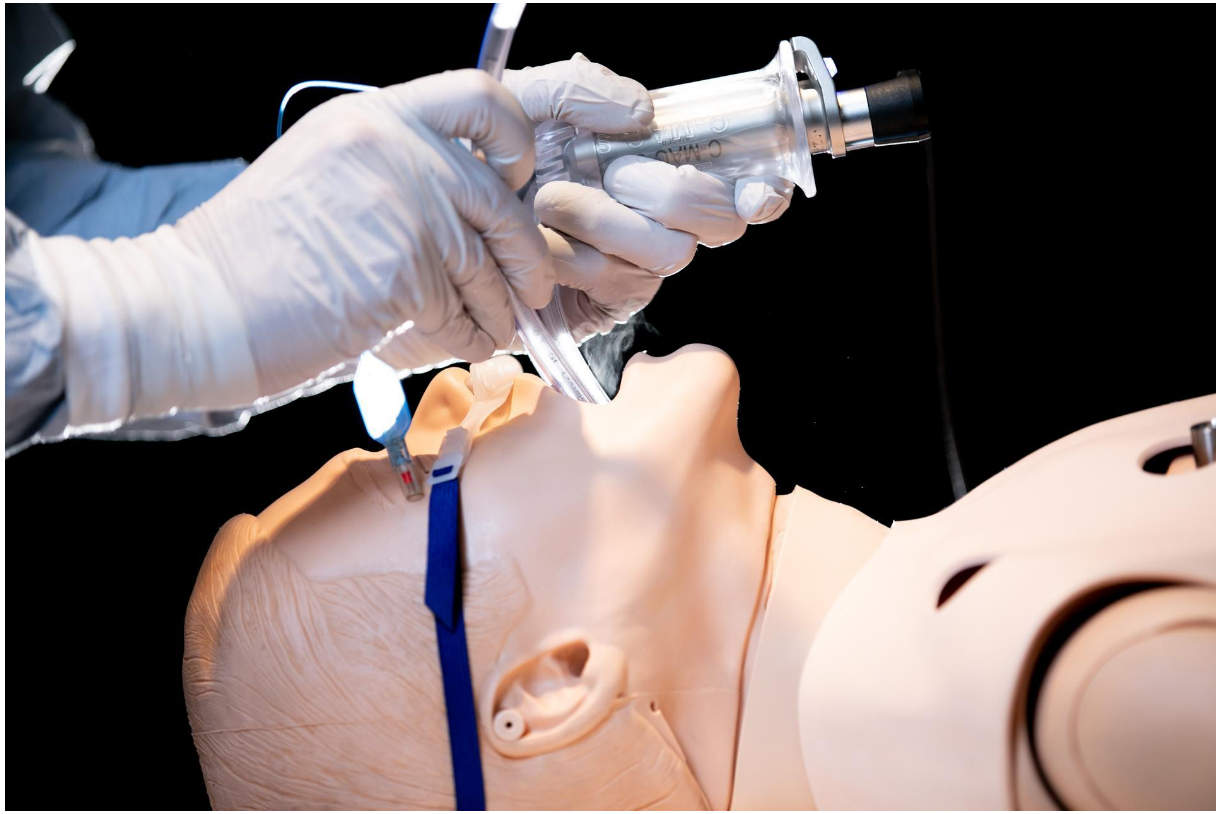

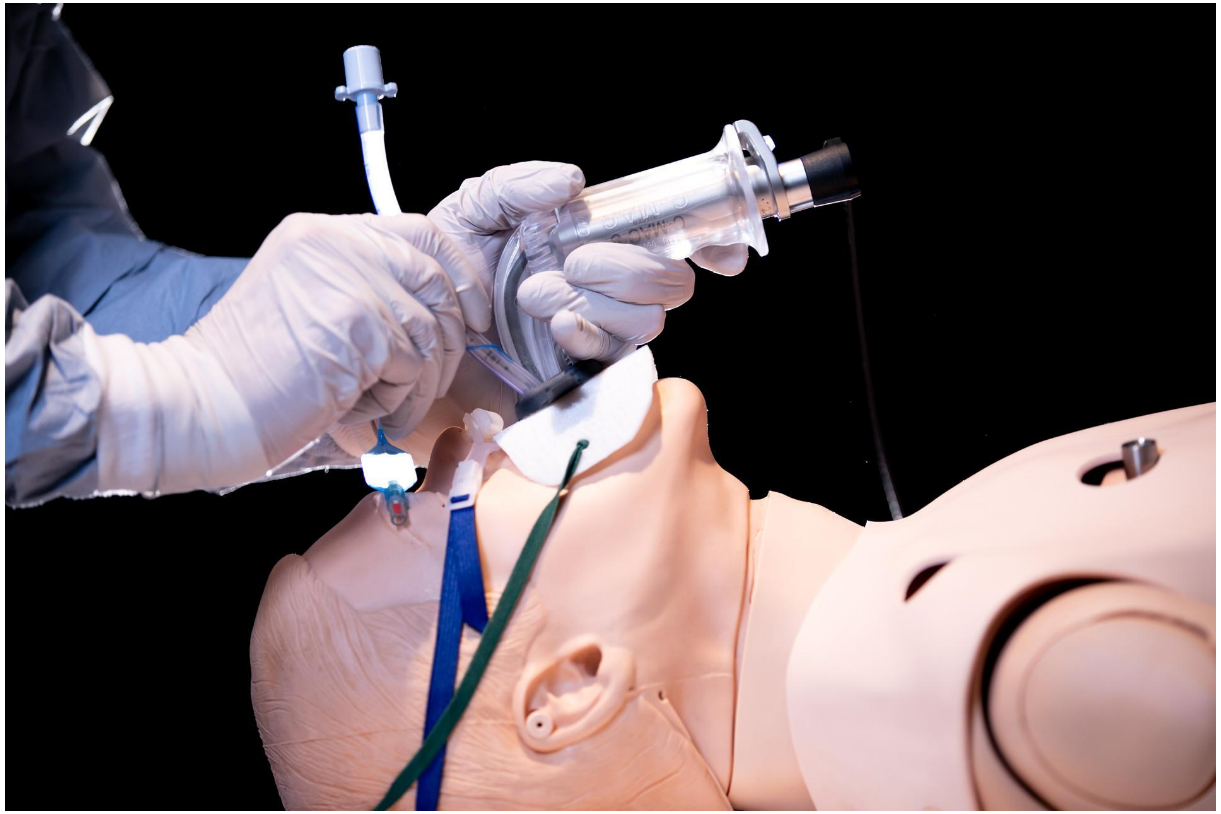

Scenario 3.3 VS 4.3

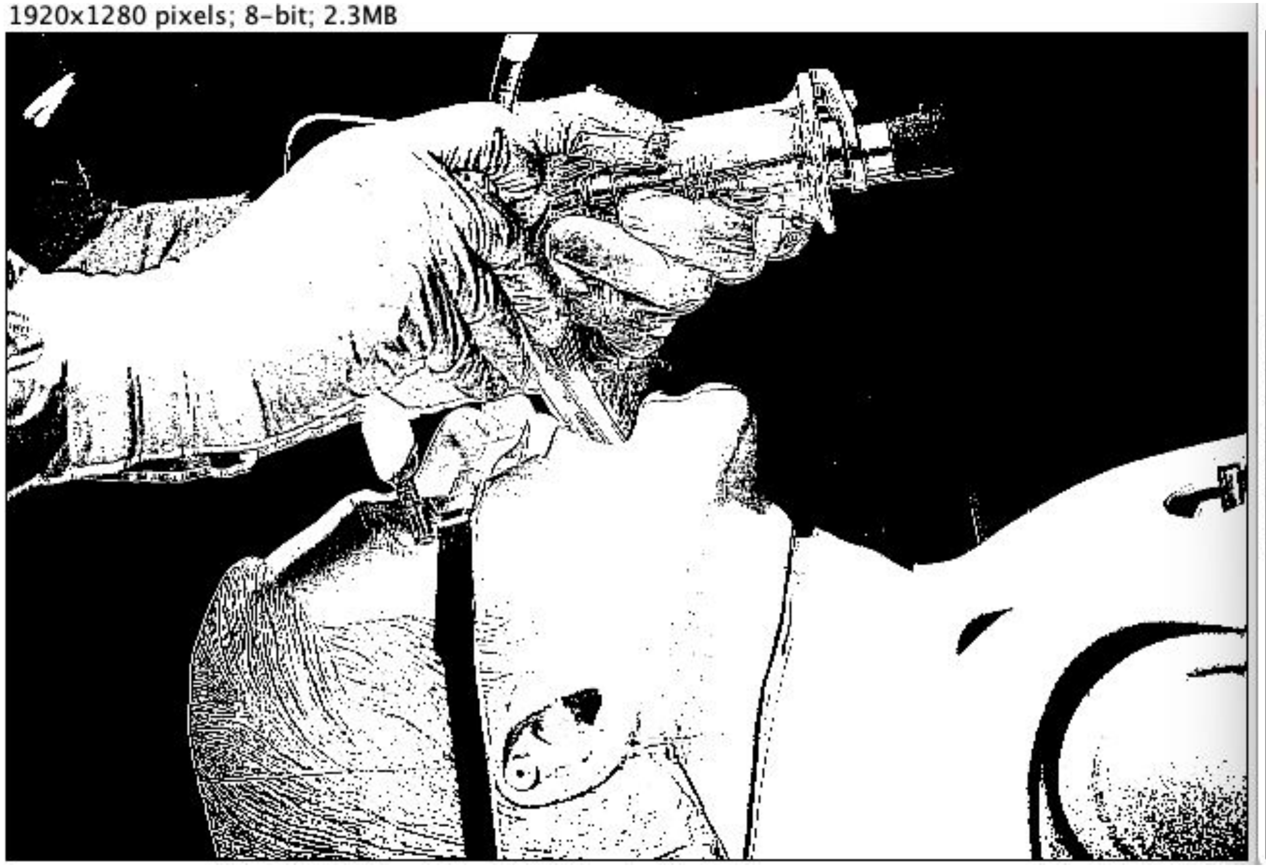


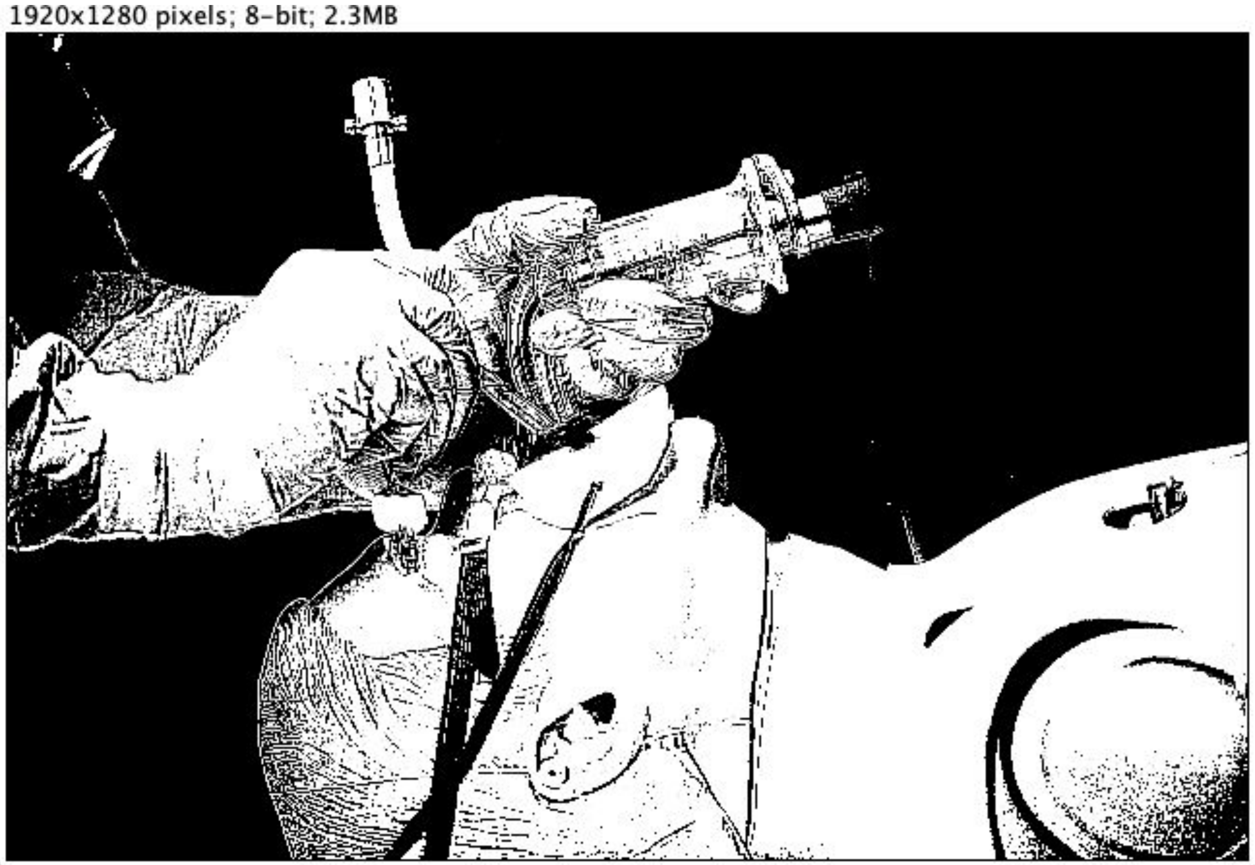

Scenario 3.3 VS 4.3

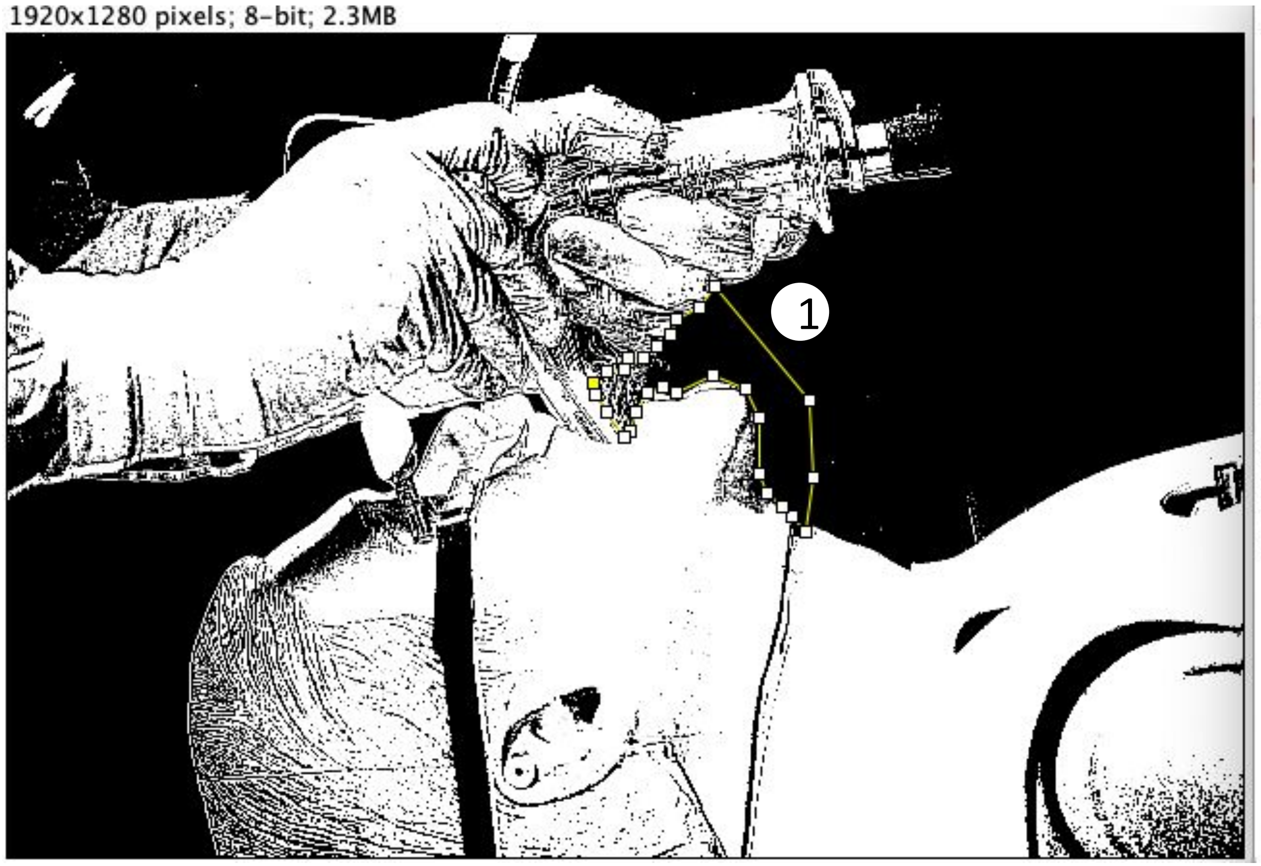


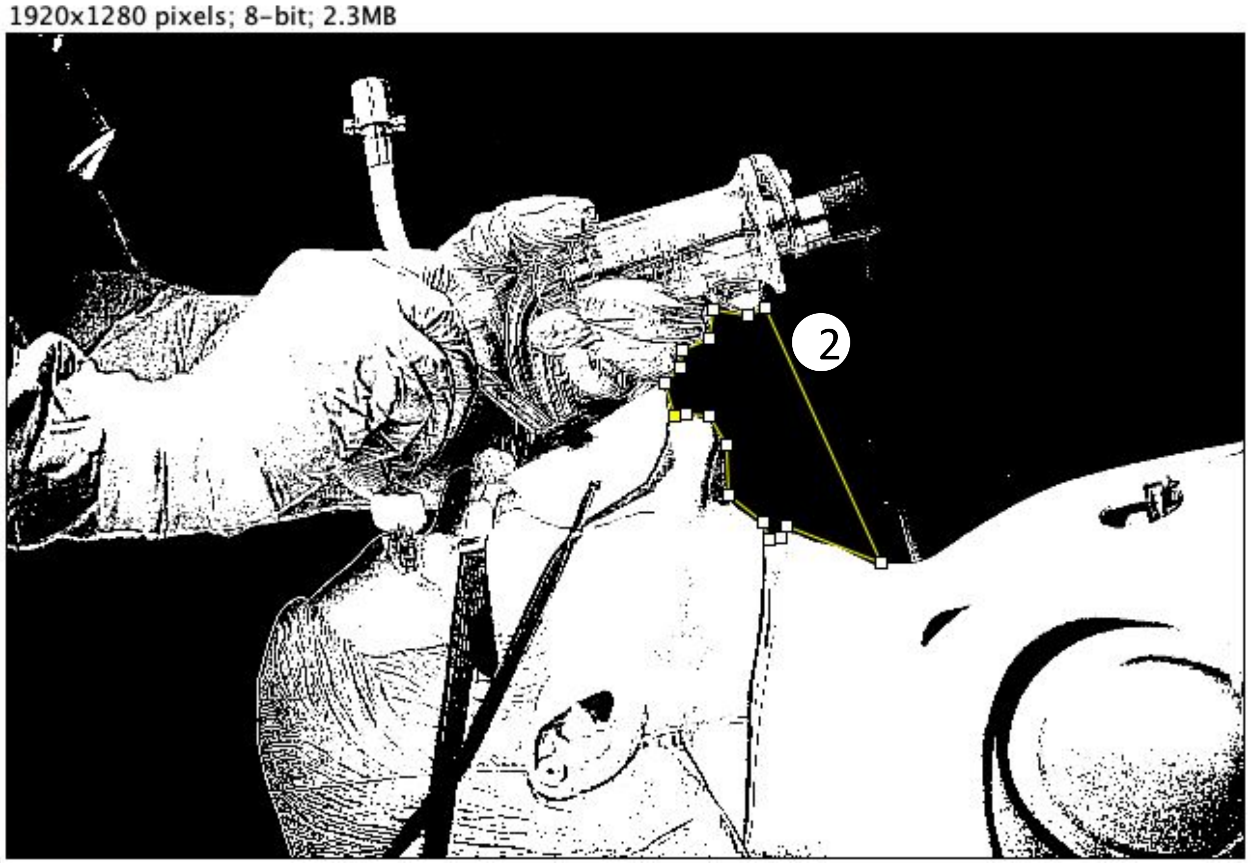

### Scenario 3.3 VS 4.3

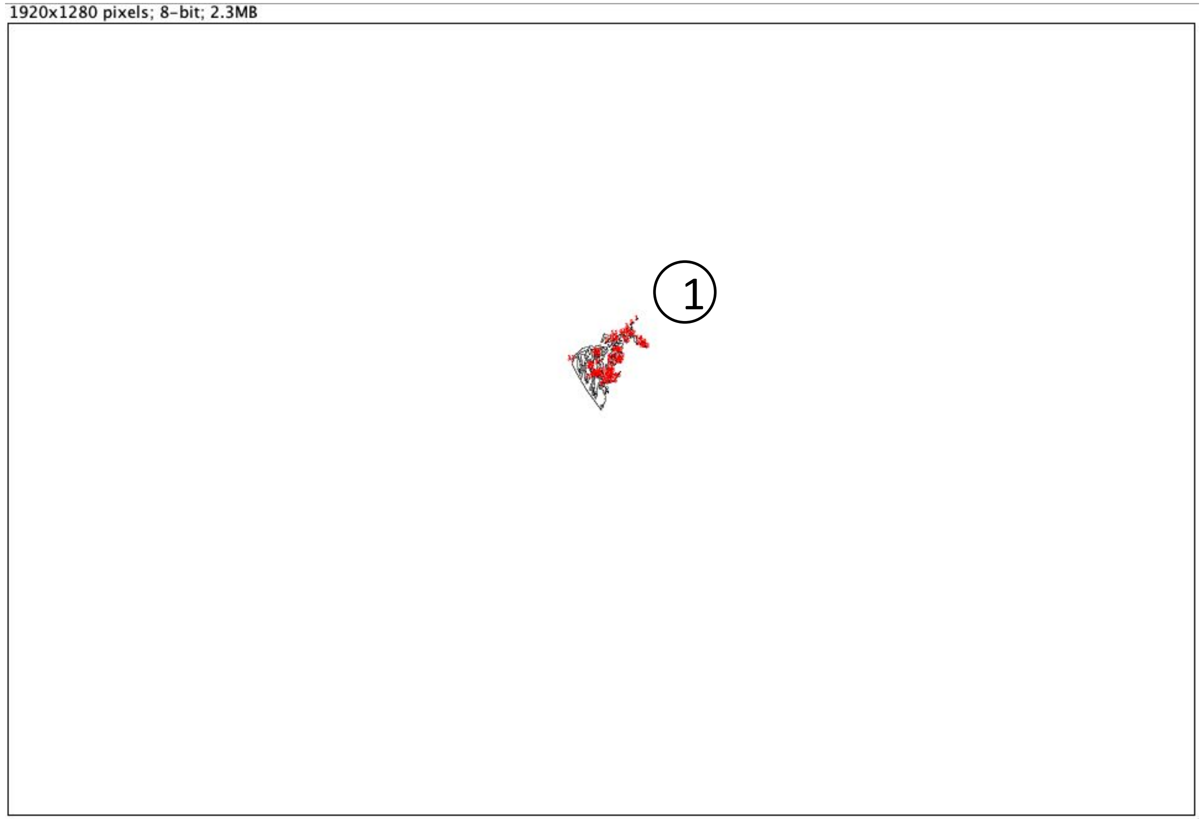


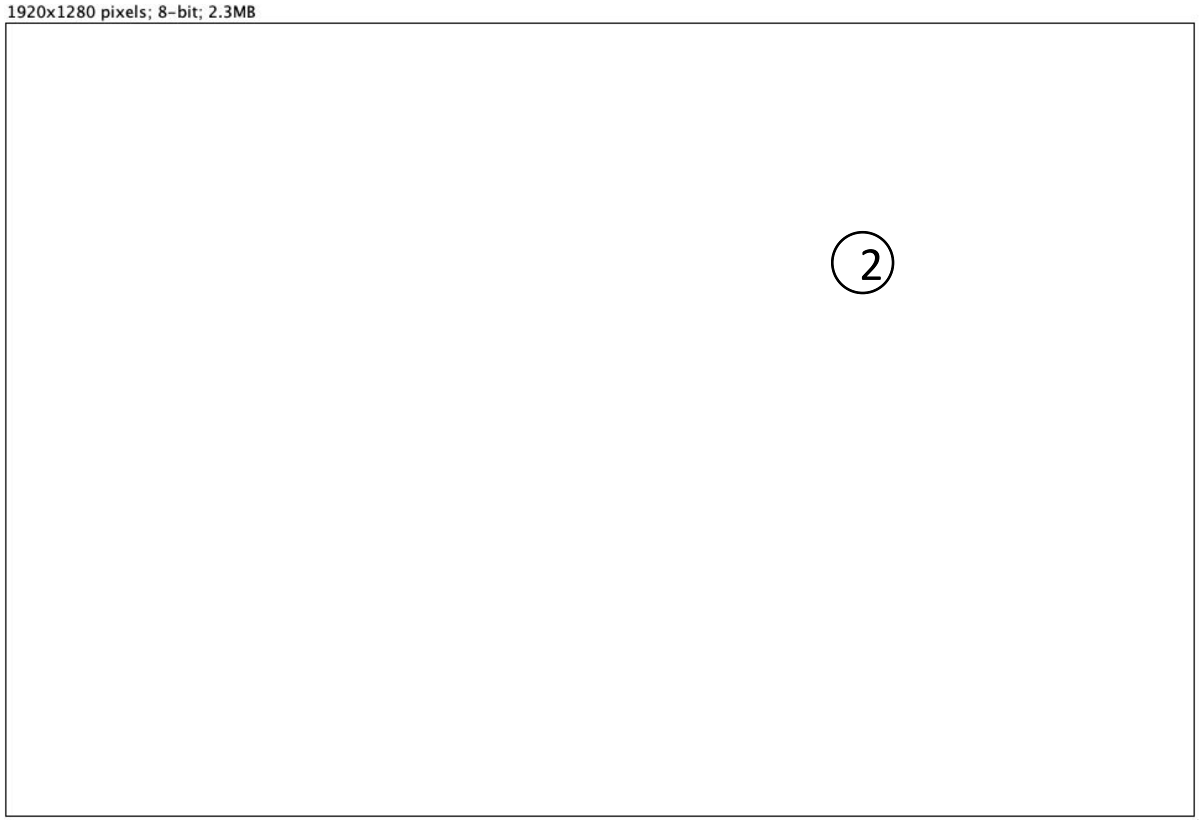


Result:

| | Slice | Count | Total Area | Average Size |
|---|---|---|---|---|
| ① | Scenario 3.3.jpg | 57 | 3256 | 57.123 |
| ② | Scenario 4.3.jpg | 0 | 0 | NaN |

***(Unit of area: PIXEL)***

*Count = number of areas*
*Total Area = total size of all areas*
*Average Size = average size of each area*

*Unit of area: PIXEL*

### Scenario 5.1 VS 6.1

*Size of the picture is 1920x1280 pixels*

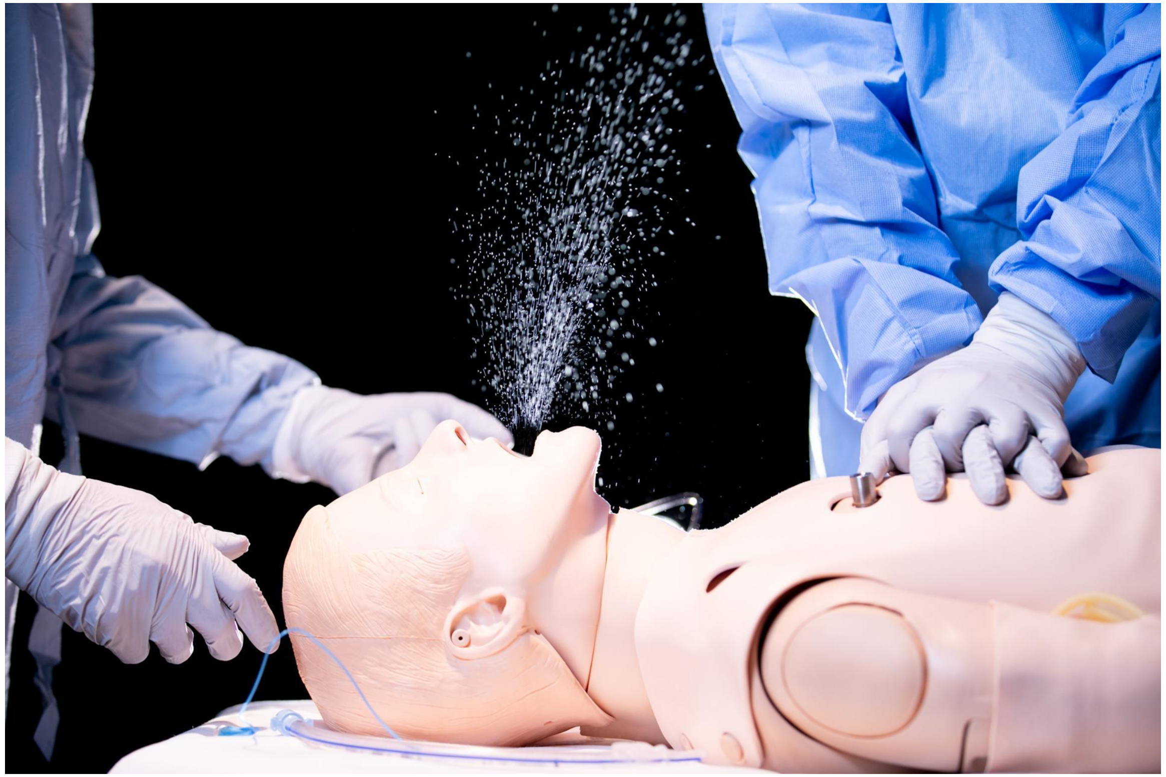

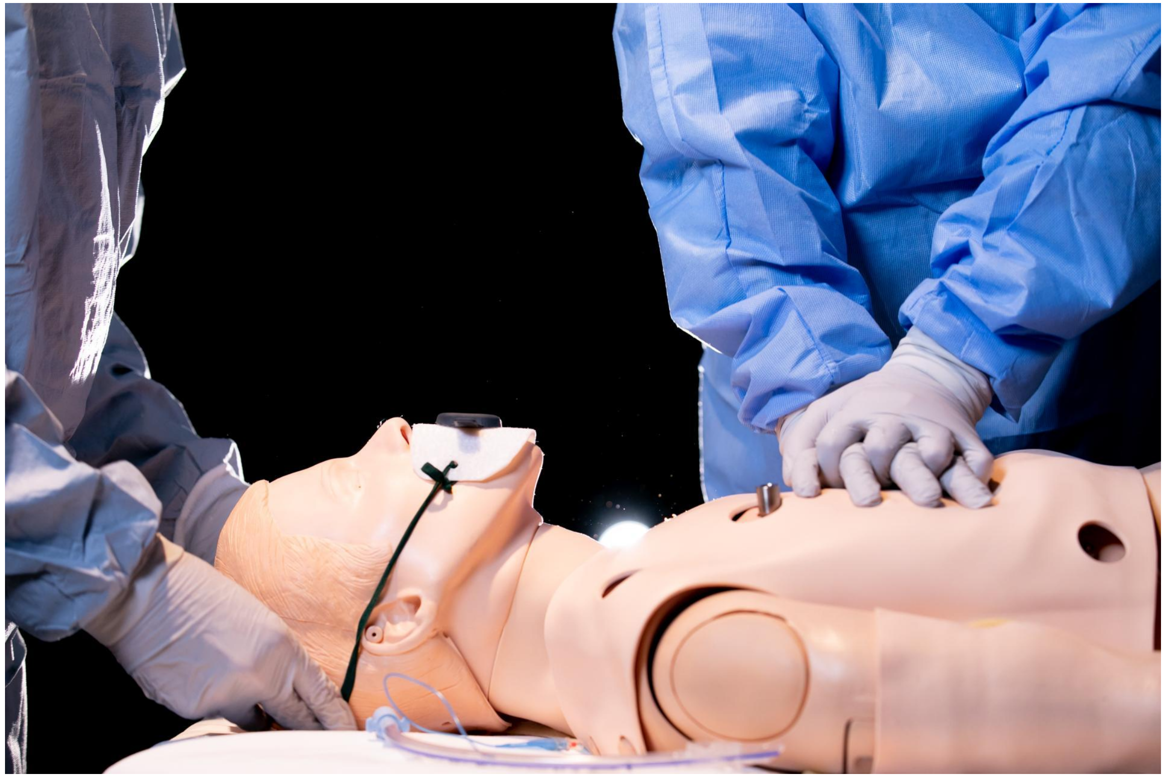

Scenario 5.1 VS 6.1

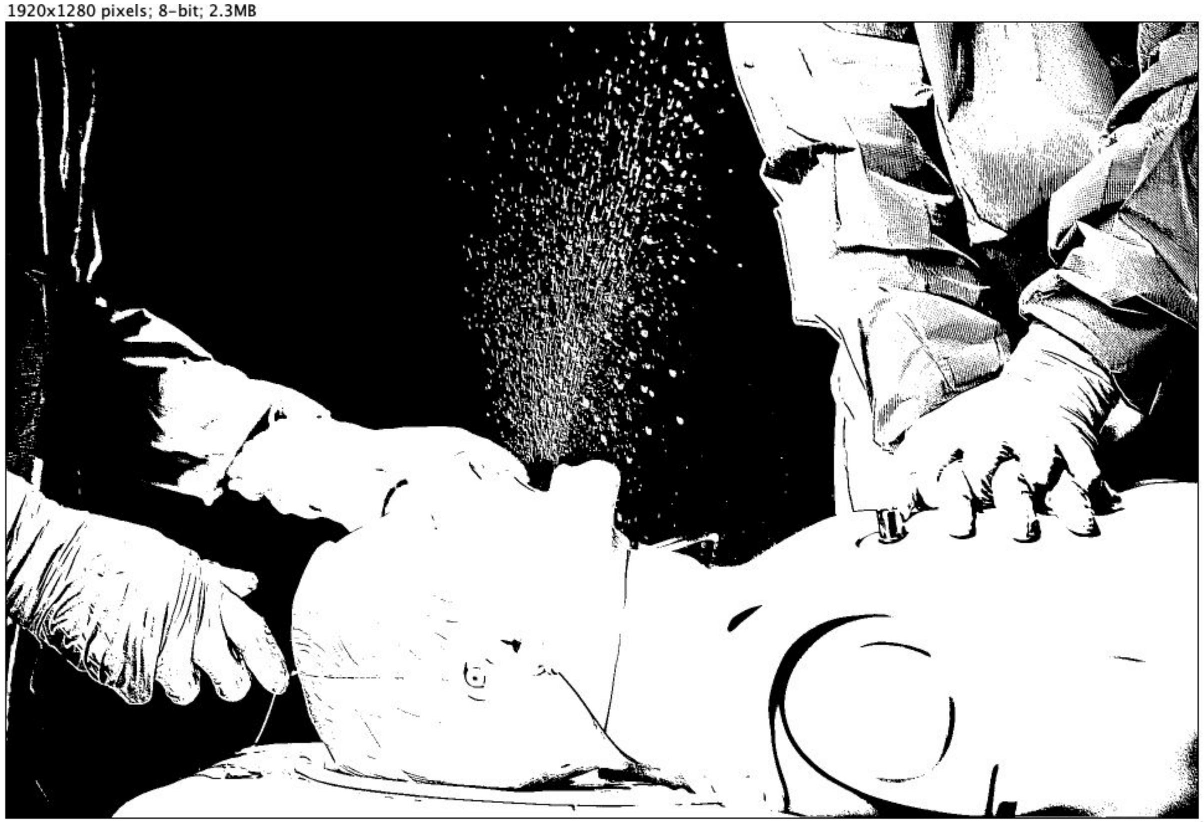


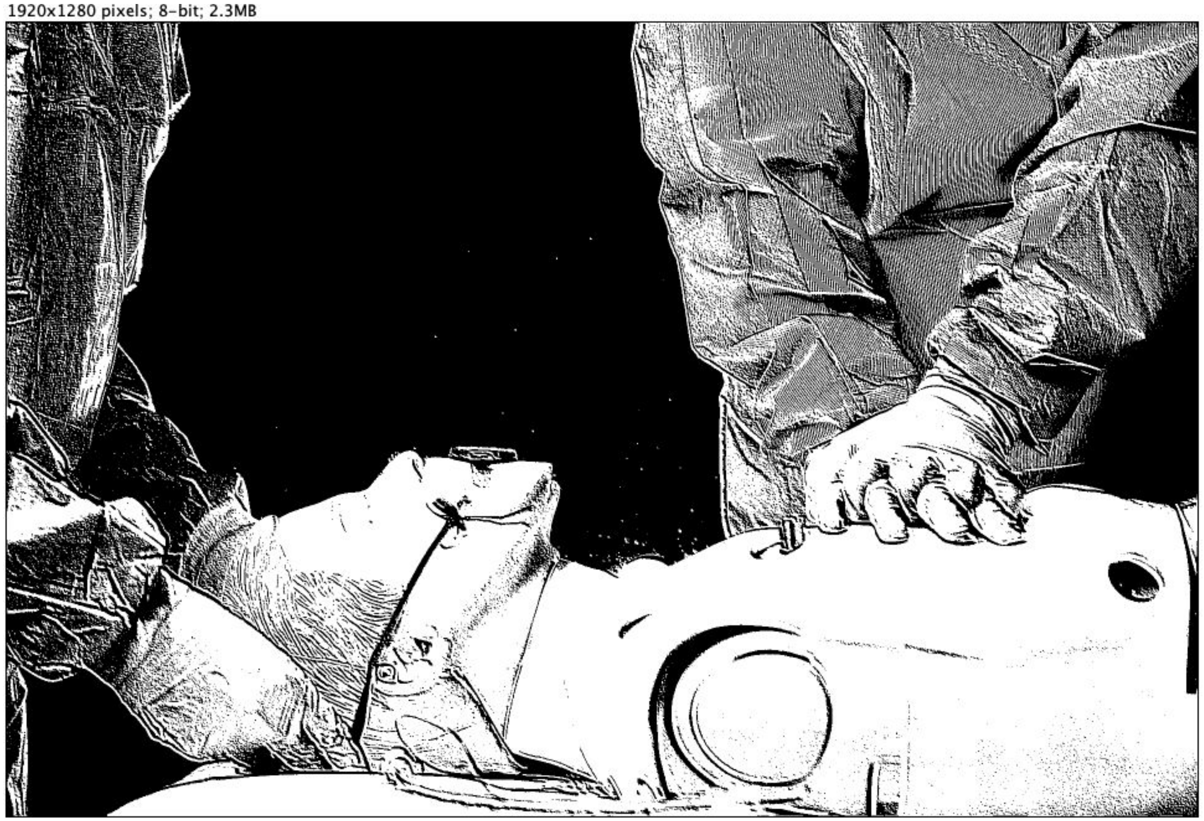

Scenario 5.1 VS 6.1

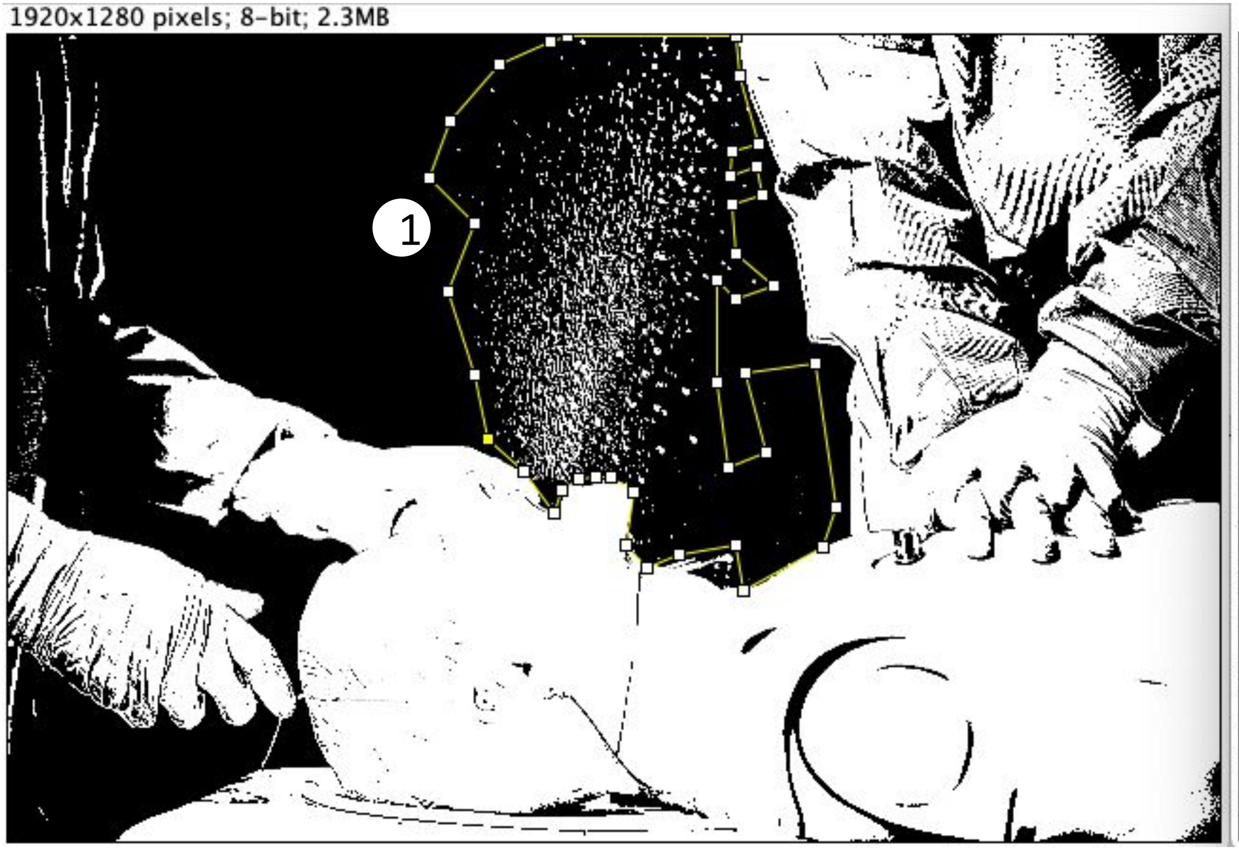


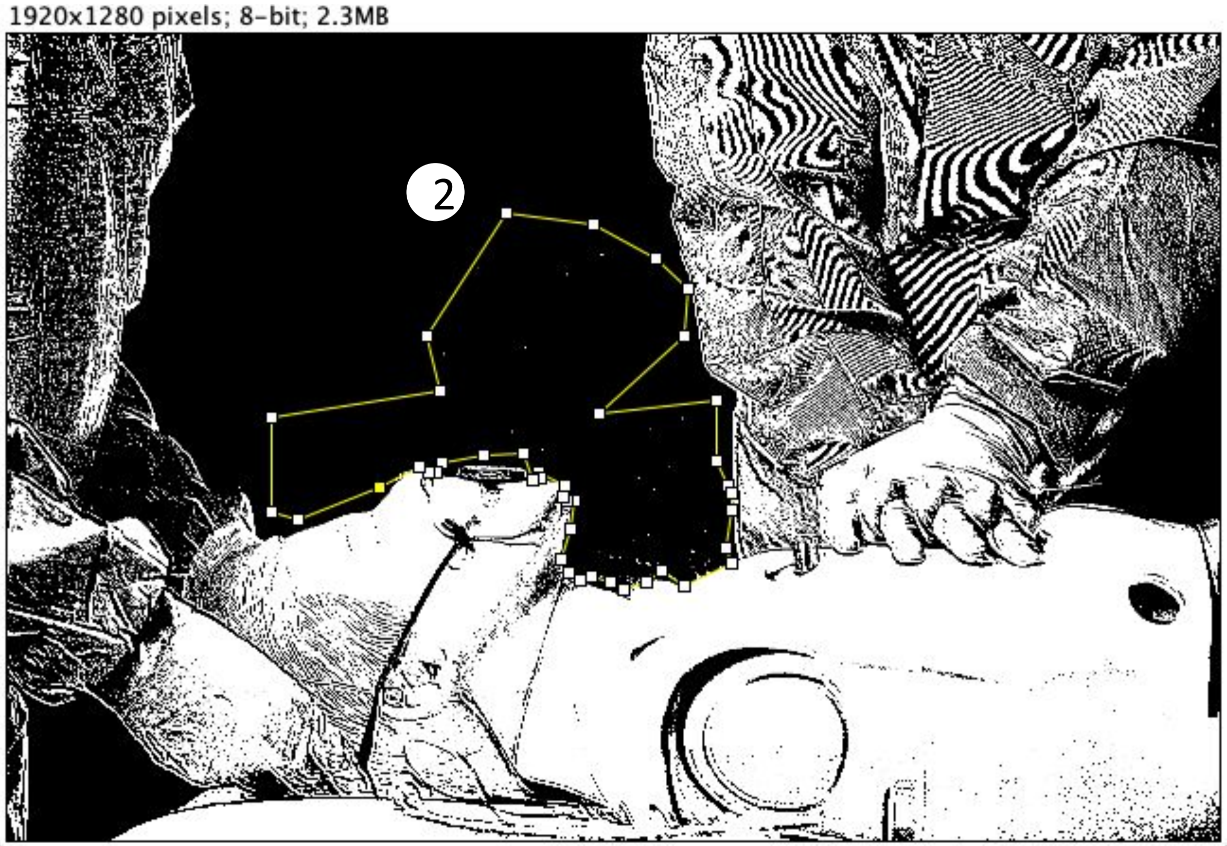

#### Scenario 5.1 VS 6.1

1920x1280 pixels; 8-bit; 2.3MB

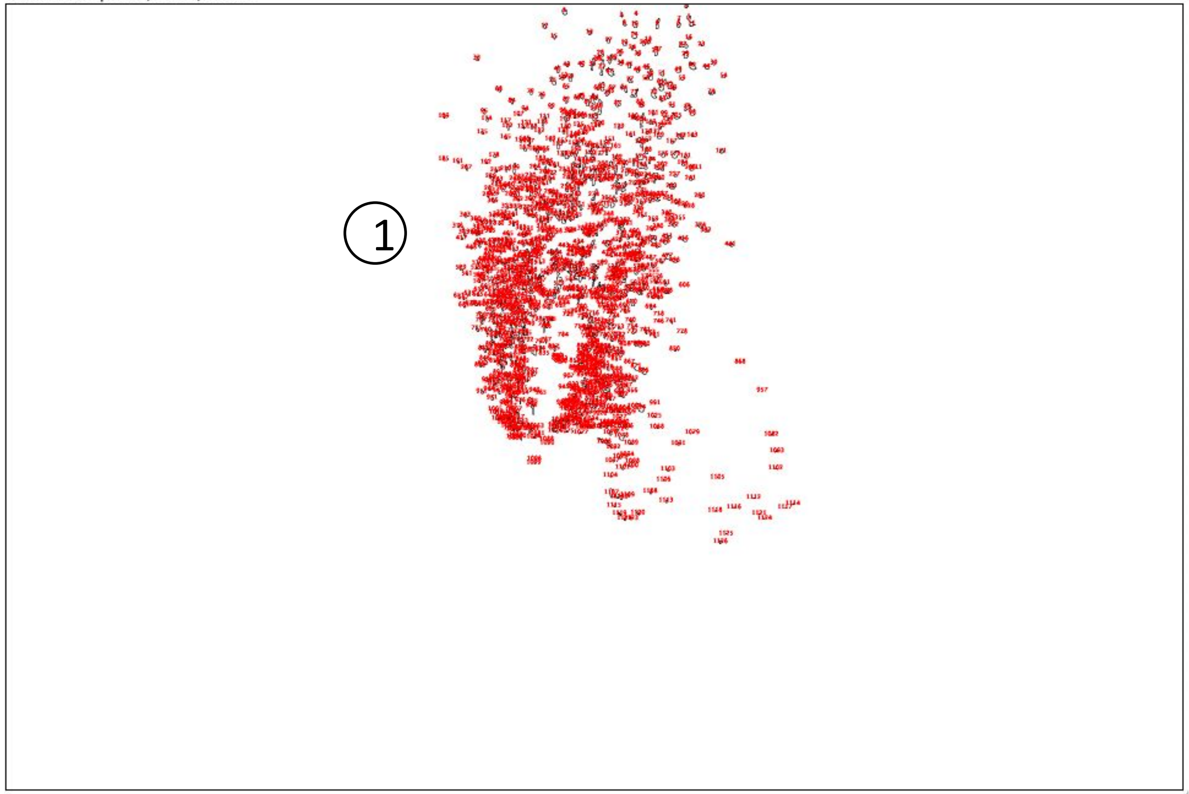

1920x1280 pixels; 8-bit; 2.3MB

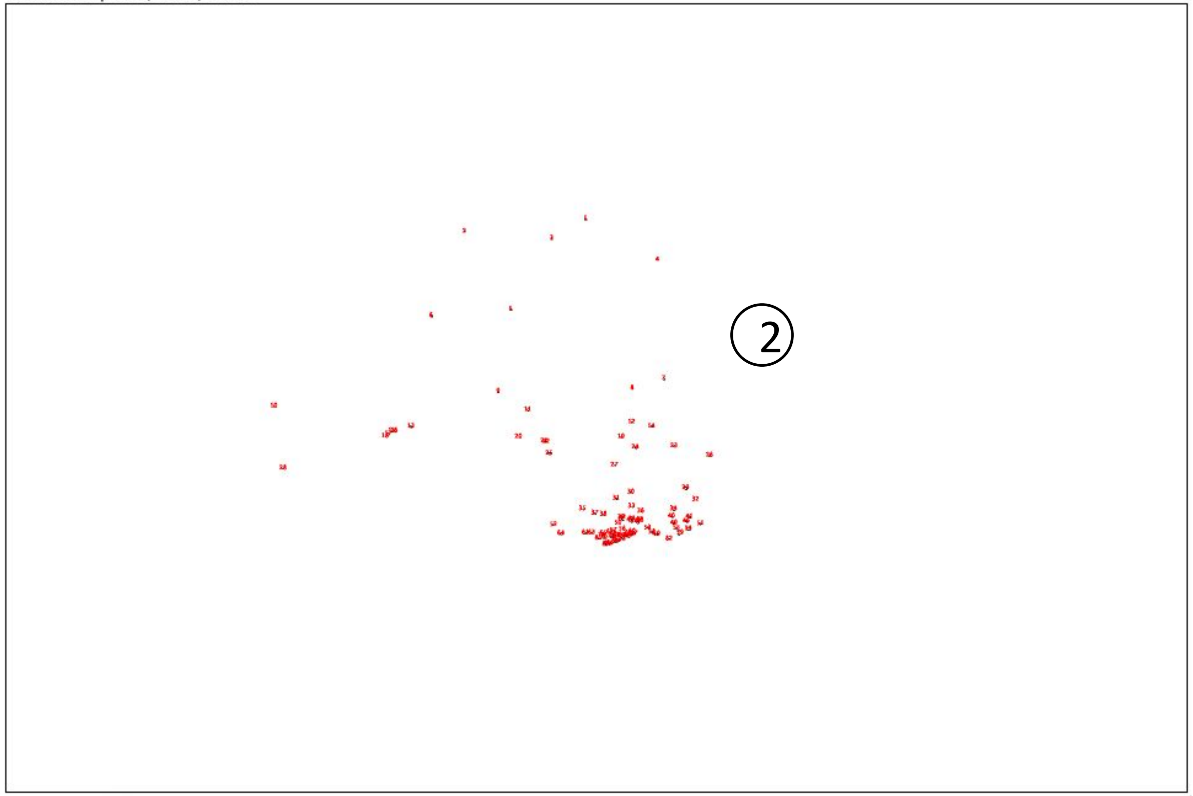

Result:

| | Slice | Count | Total Area | Average Size |
|---|---|---|---|---|
| 1 | Scenario 5.1.jpg | 1126 | 21919 | 19.466 |
| 2 | Scenario 6.1.JPG | 87 | 616 | 7.080 |

***(Unit of area: PIXEL)***

*Count = number of areas*
*Total Area = total size of all areas*
*Average Size = average size of each area*

*Unit of area: PIXEL*

### Scenario 5.2 VS 6.2

*Size of the picture is 1920x1280 pixels*

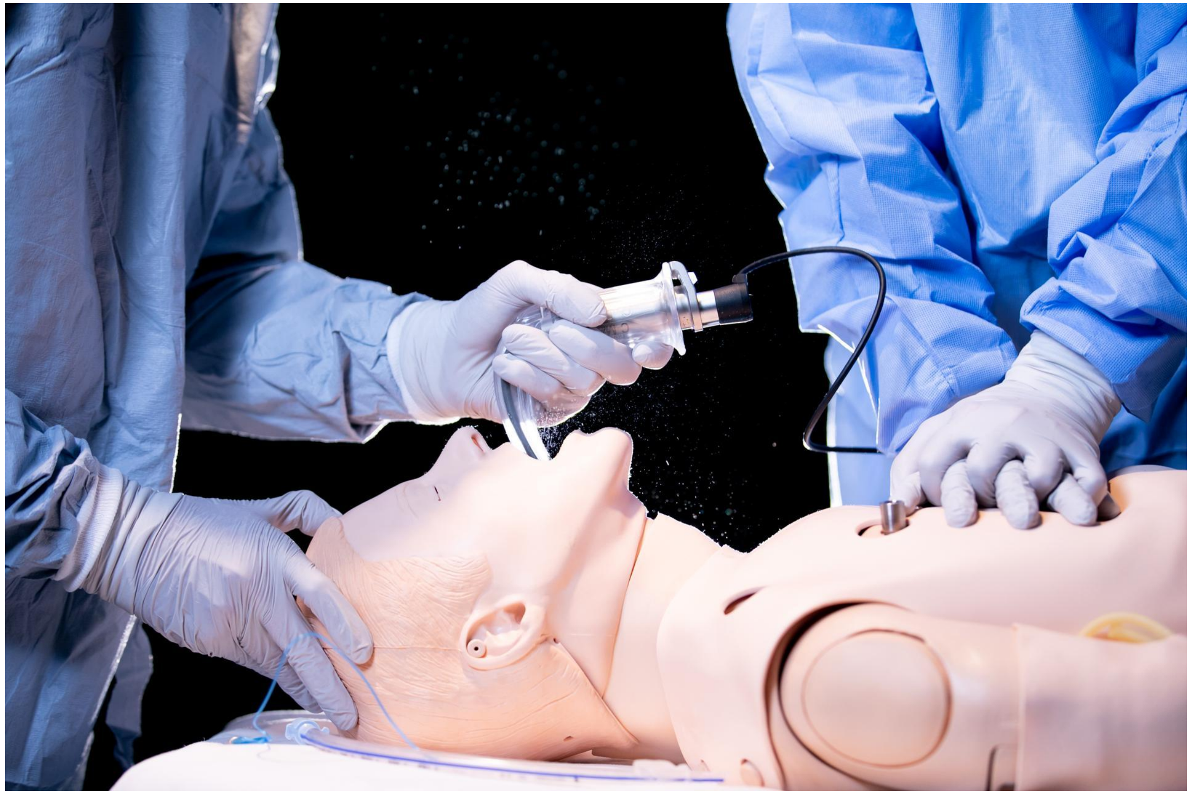

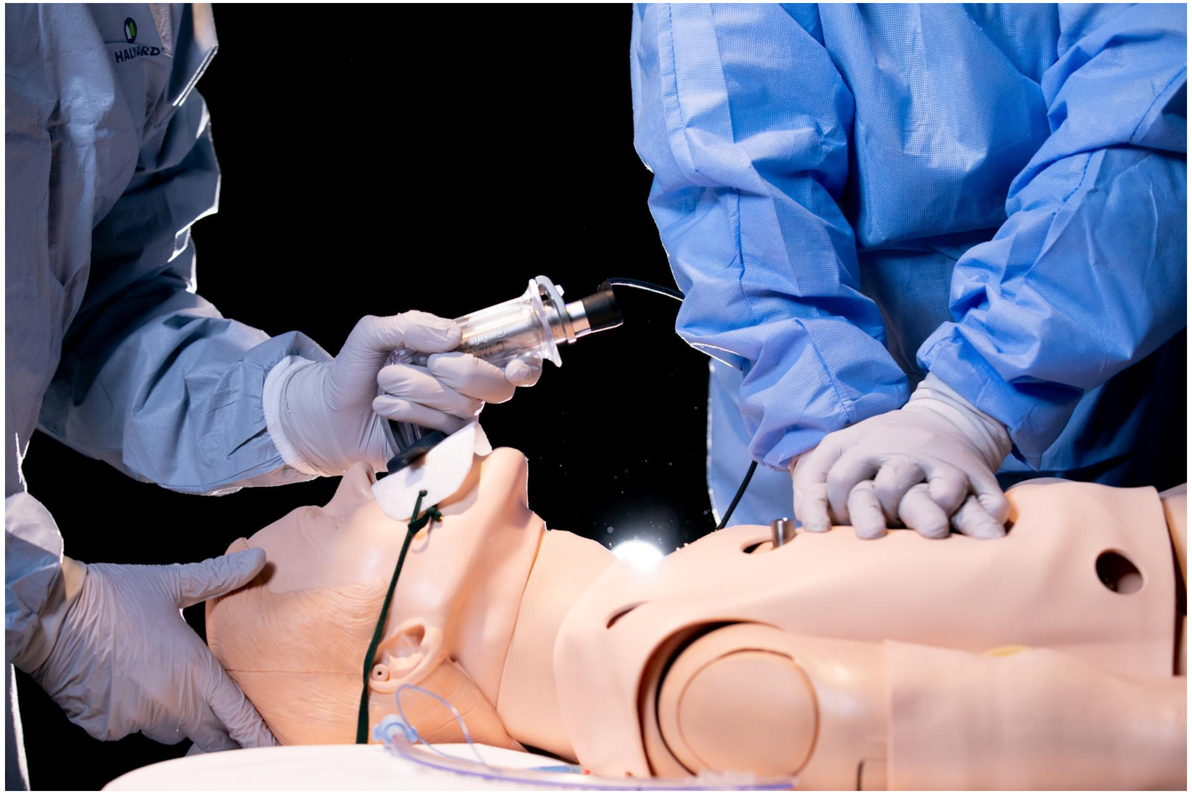

### Scenario 5.2 VS 6.2

1920x1280 pixels; 8-bit; 2.3MB

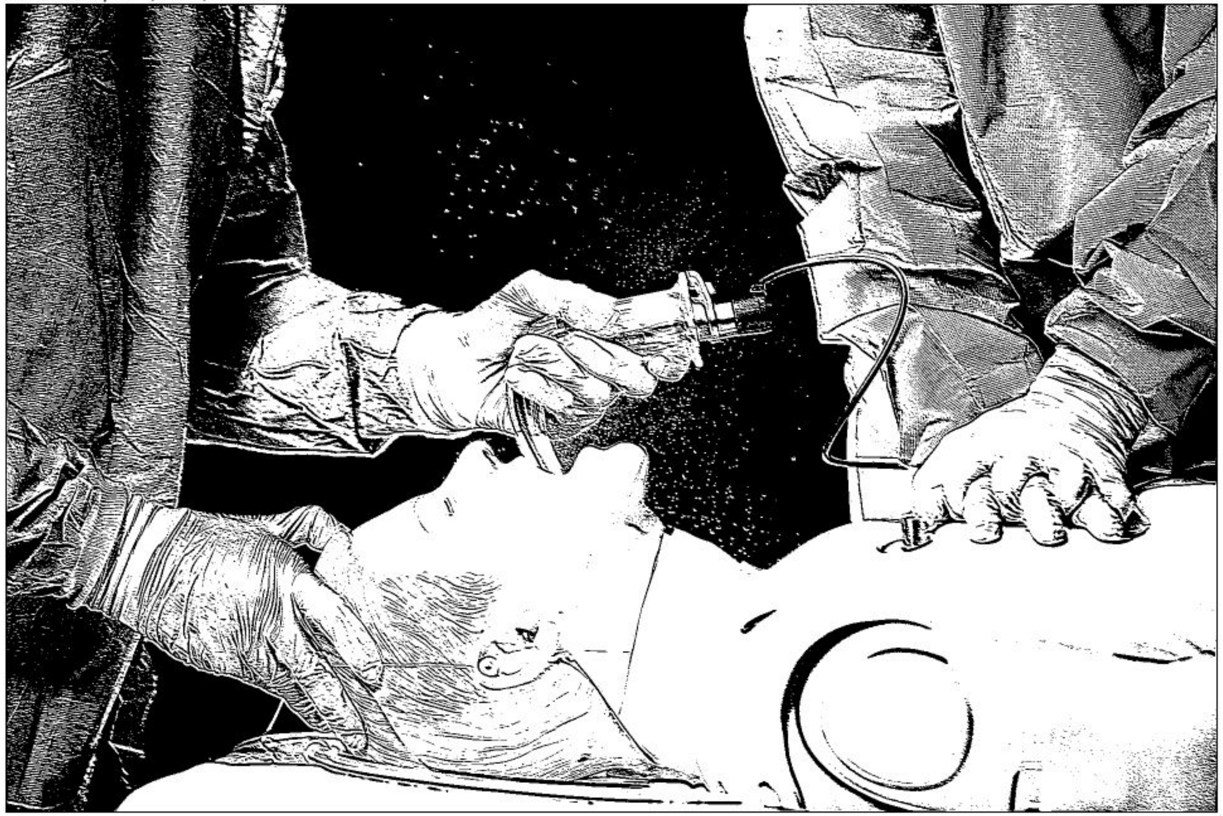

1920x1280 pixels; 8-bit; 2.3MB

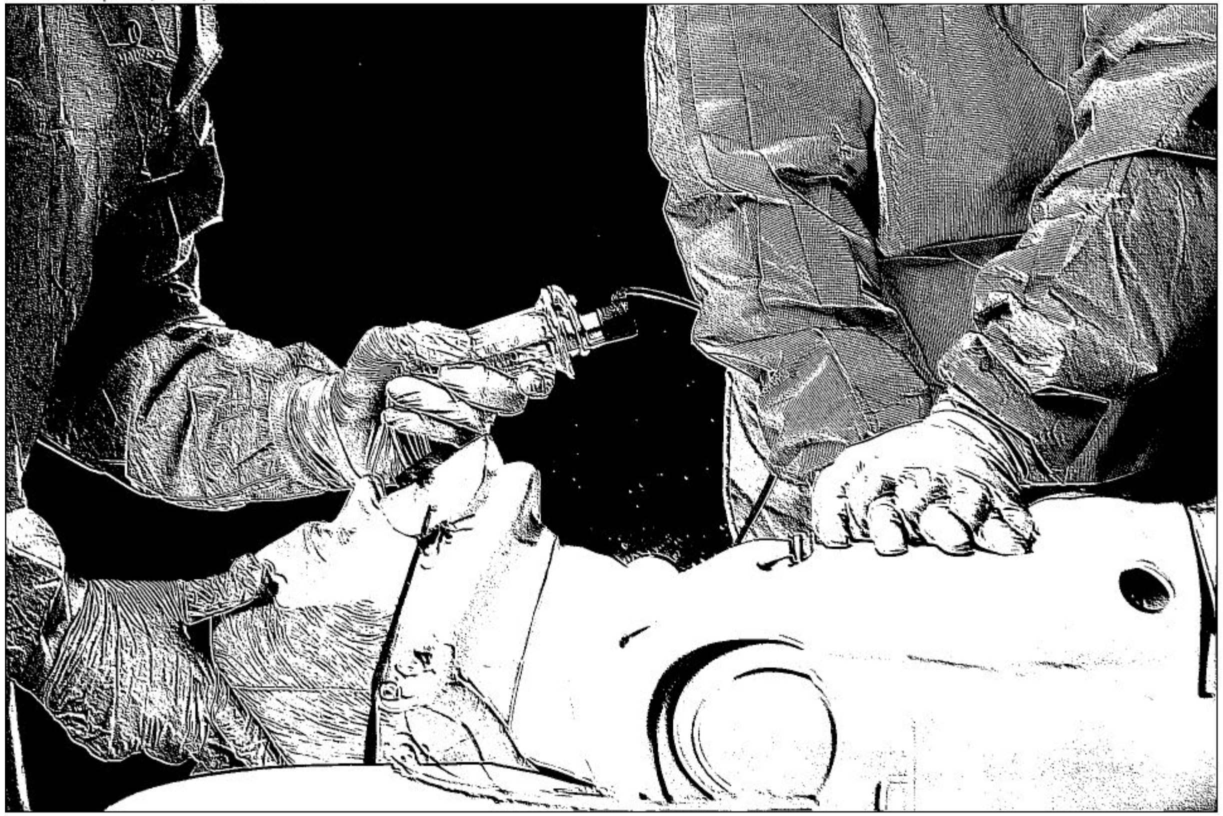

### Scenario 5.2 VS 6.2

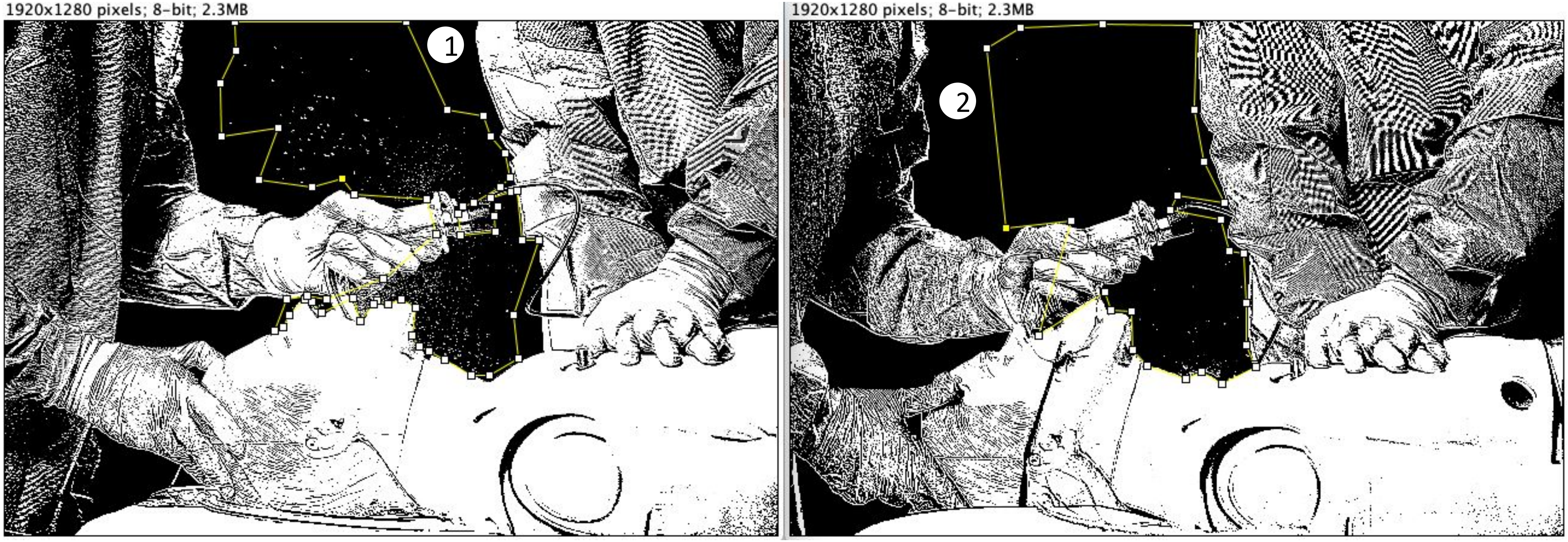

### Scenario 5.2 VS 6.2

1920x1280 pixels; 8-bit; 2.3MB

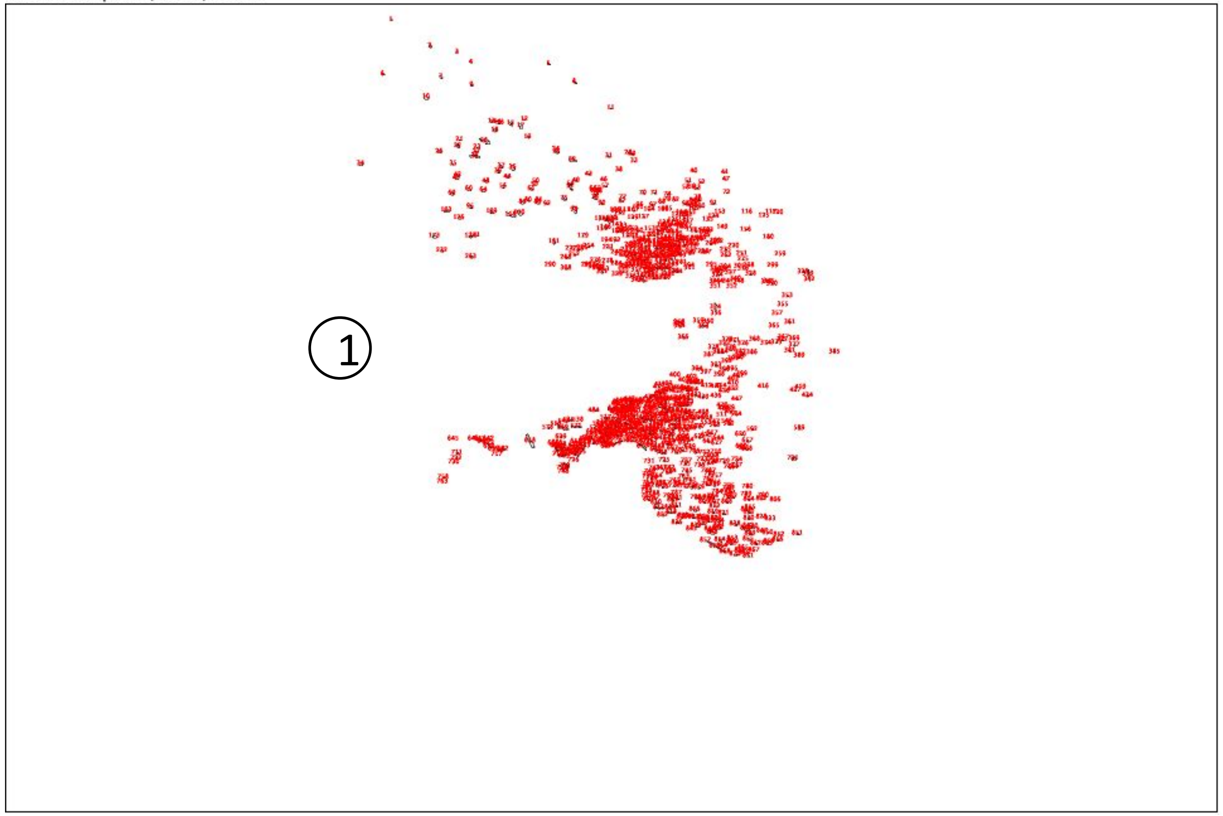


1920x1280 pixels; 8-bit; 2.3MB

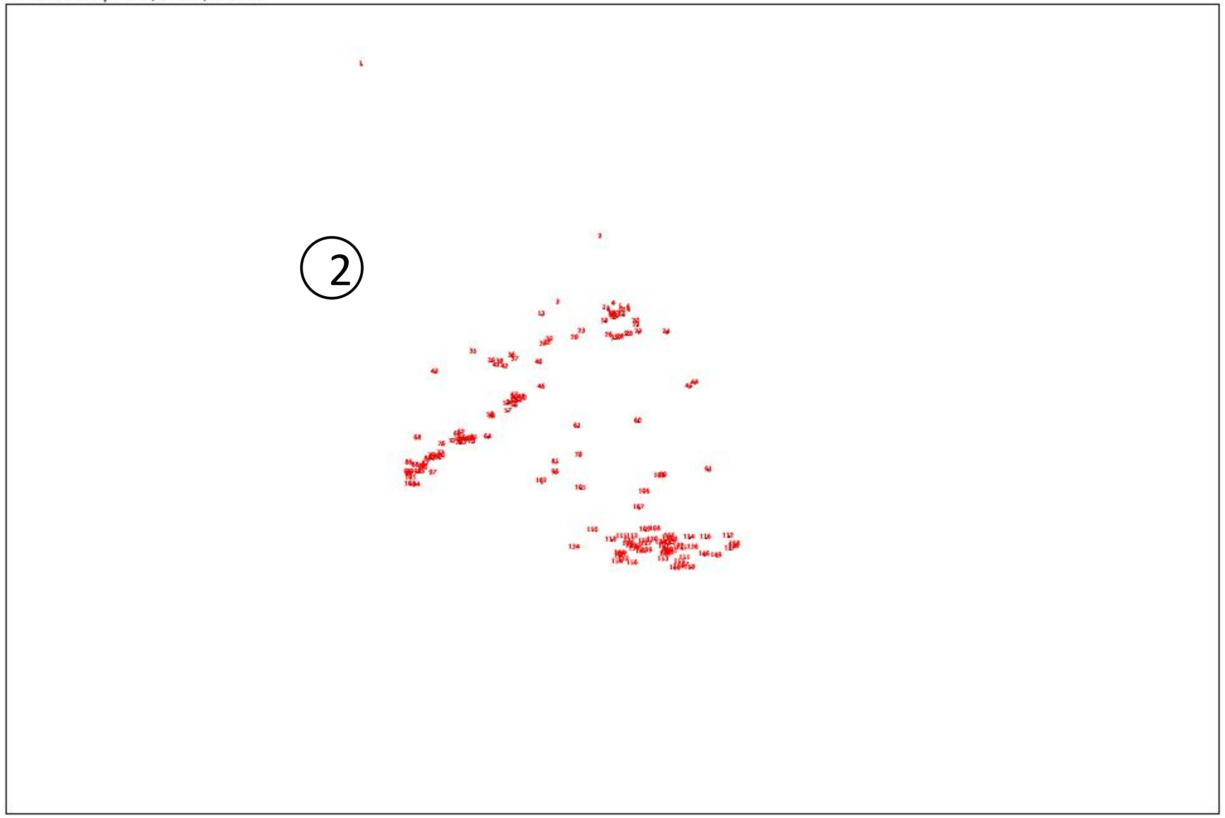


Result:

| | Slice | Count | Total Area | Average Size |
|---|---|---|---|---|
| 1 | Scenario 5.2.jpg | 871 | 7178 | 8.241 |
| 2 | Scenario 6.2.JPG | 160 | 616 | 3.850 |

***(Unit of area: PIXEL)***

*Count = number of areas*
*Total Area = total size of all areas*
*Average Size = average size of each area*

*Unit of area: PIXEL*

### Scenario 5.3 VS 6.3

*Size of the picture is 1920x1280 pixels*

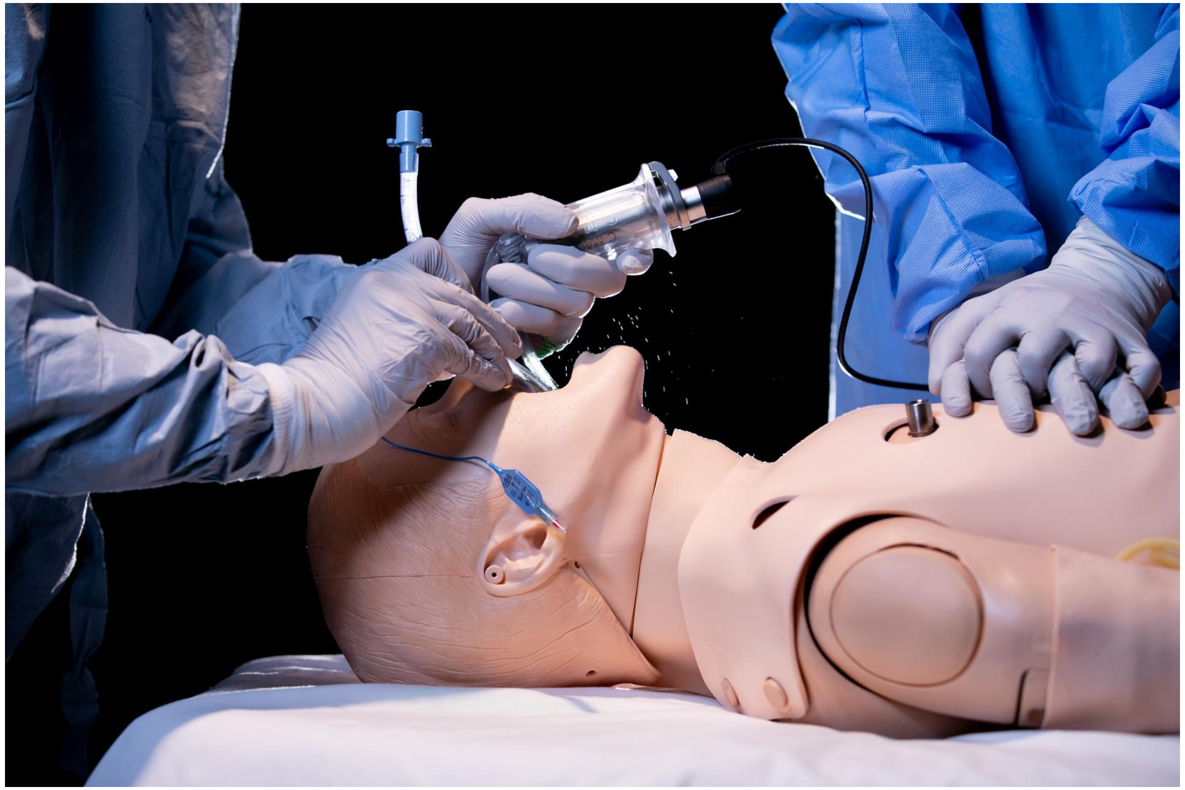

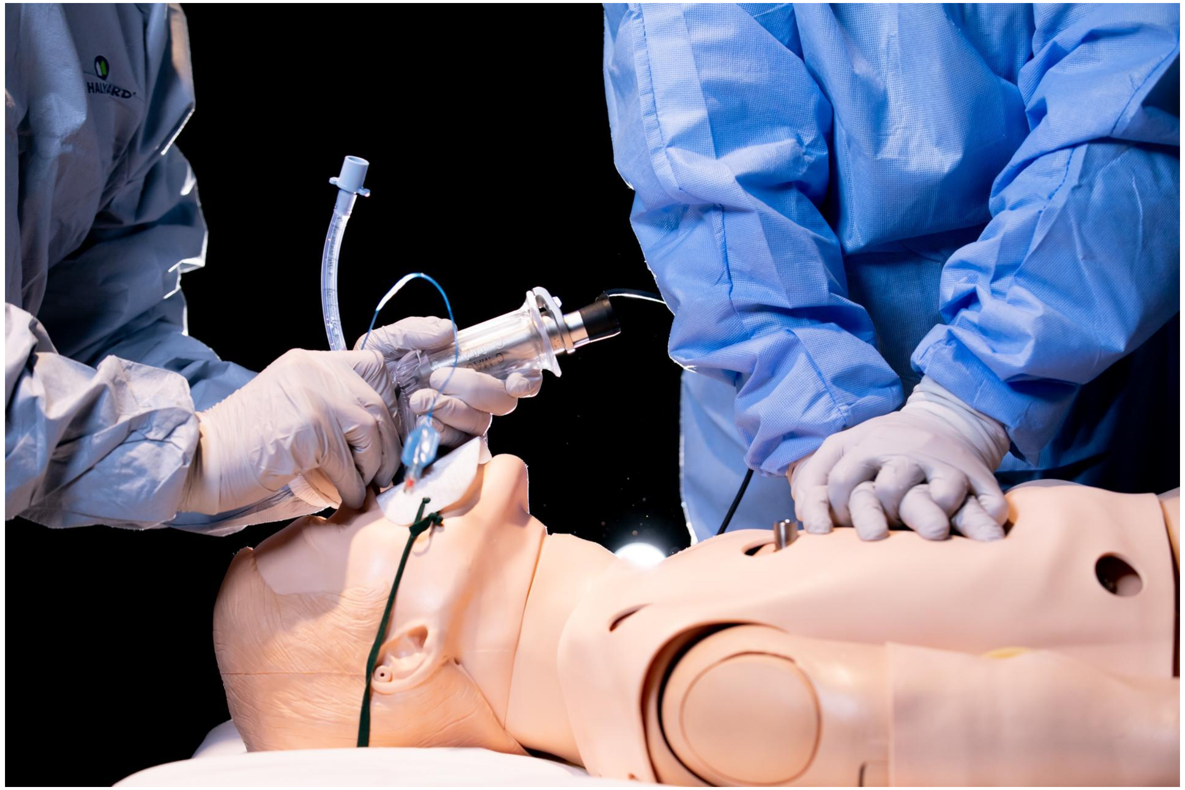

### Scenario 5.3 VS 6.3

1920x1280 pixels; 8-bit; 2.3MB

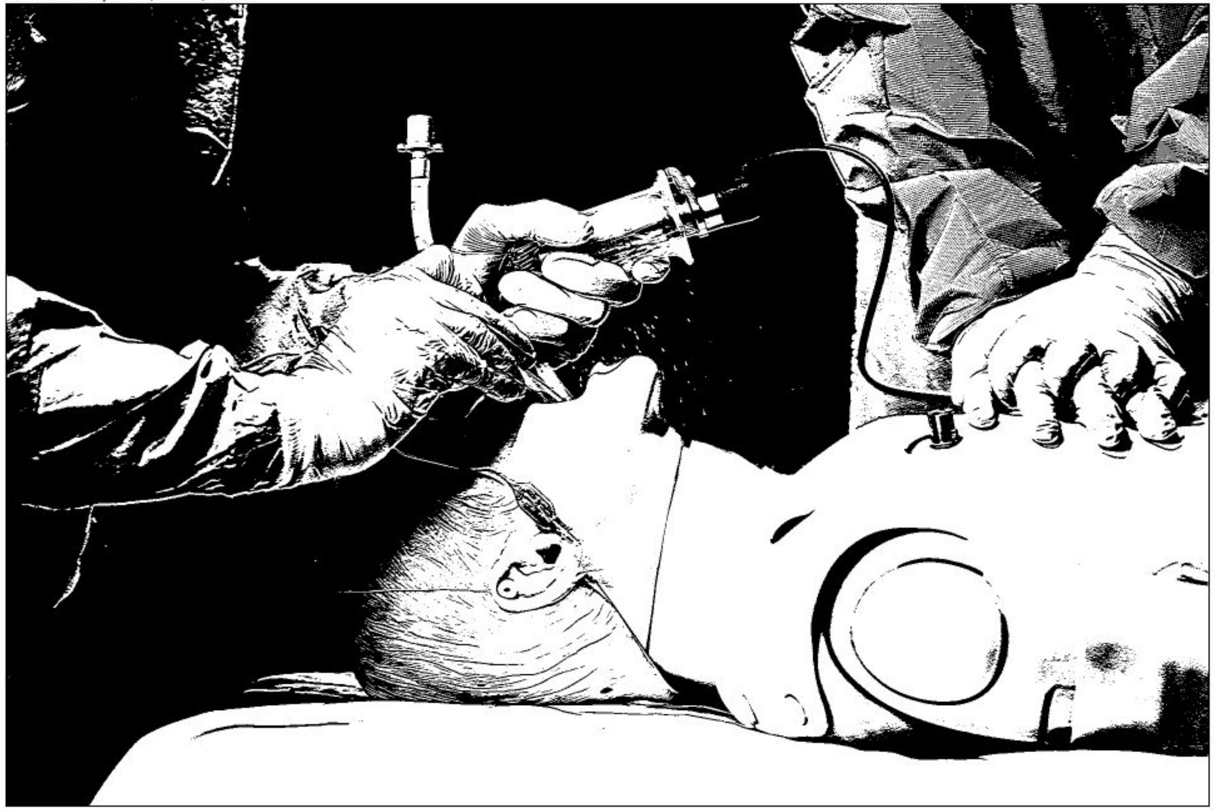

1920x1280 pixels; 8-bit; 2.3MB

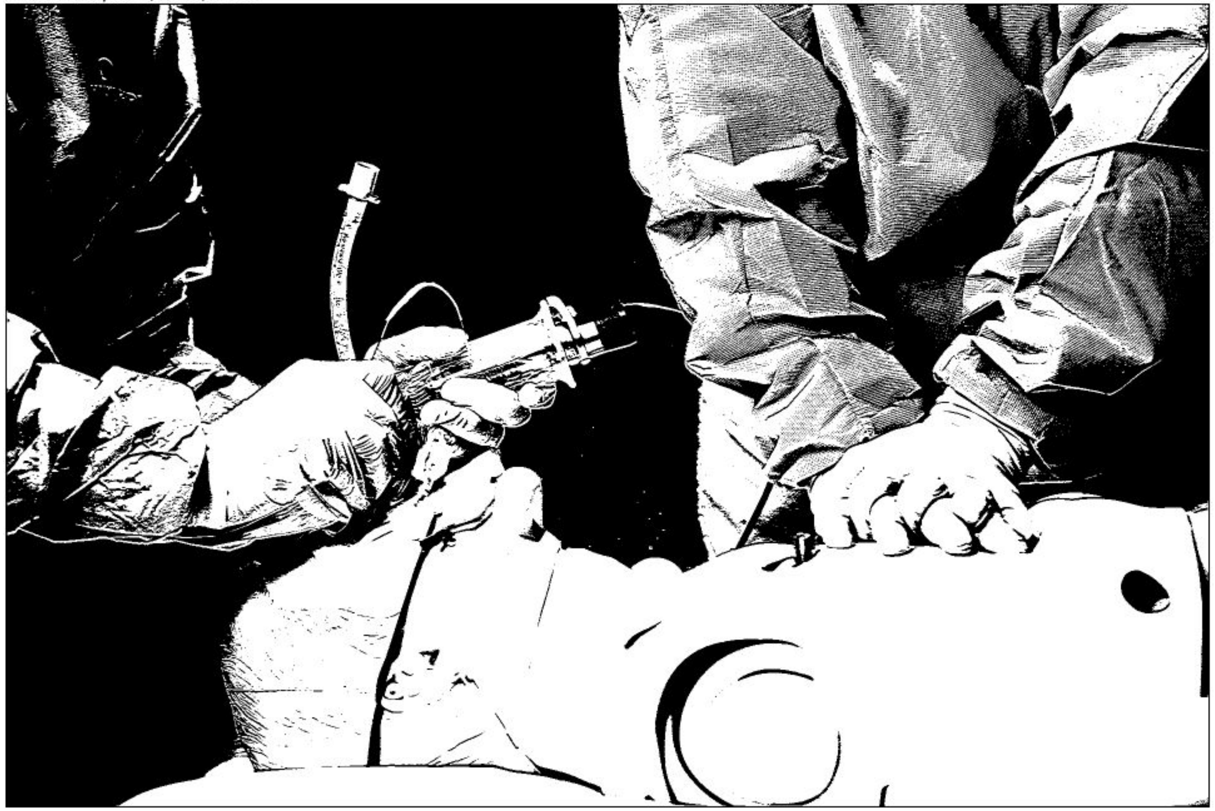

### Scenario 5.3 VS 6.3

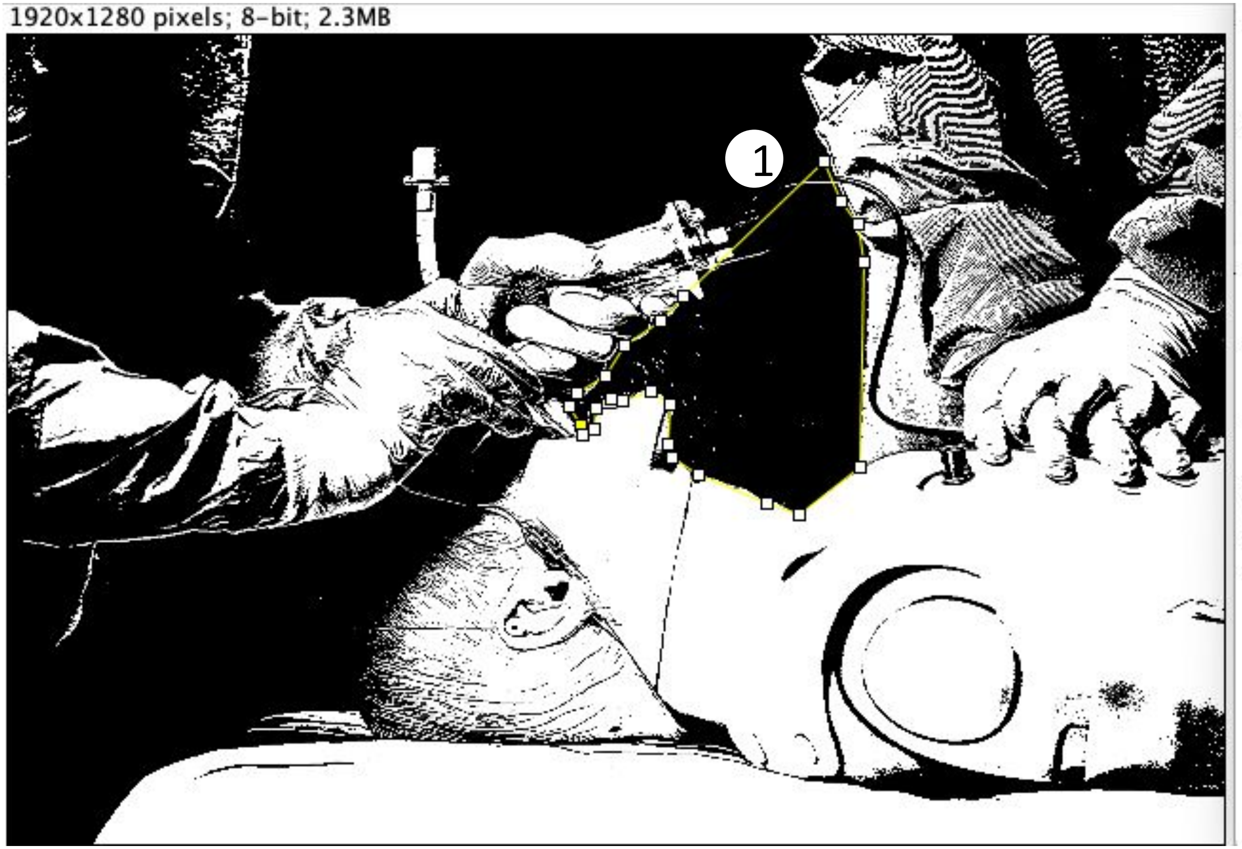


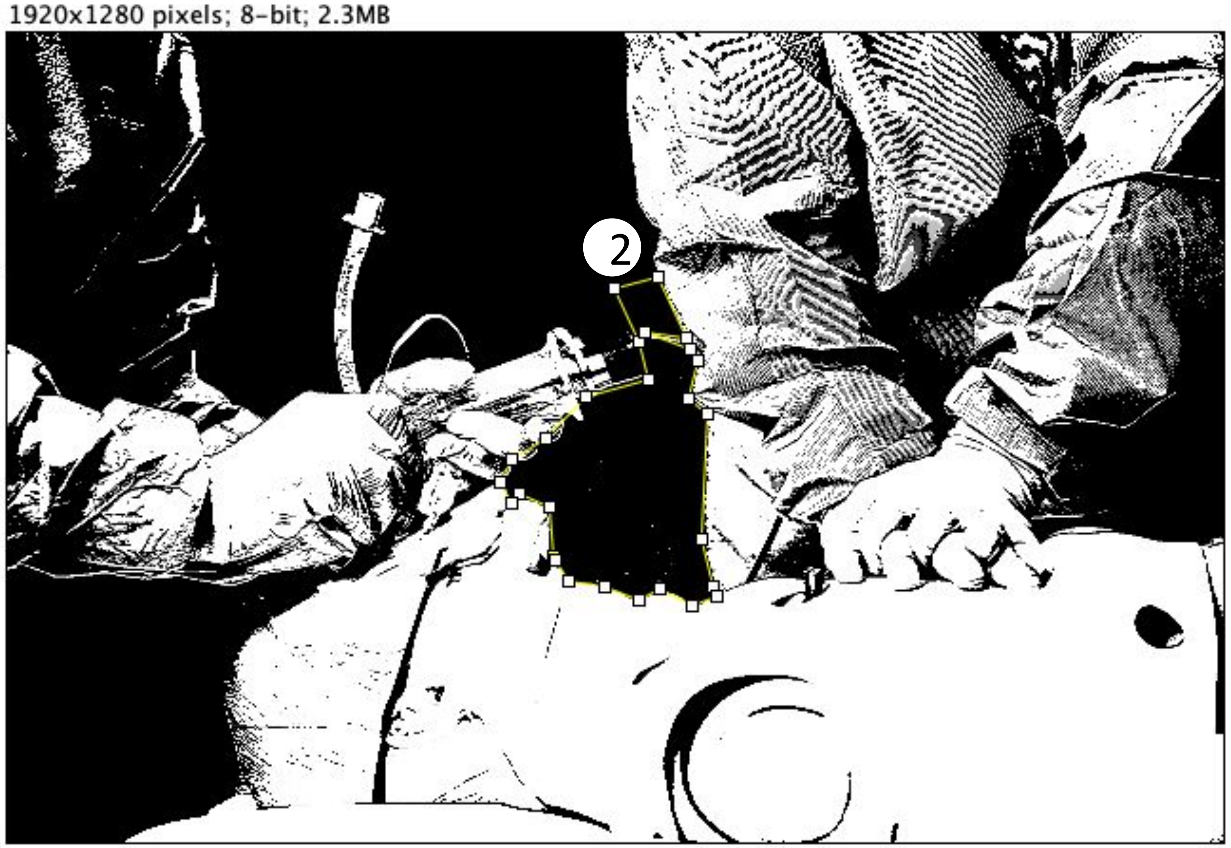

### Scenario 5.3 VS 6.3

1920x1280 pixels; 8-bit; 2.3MB

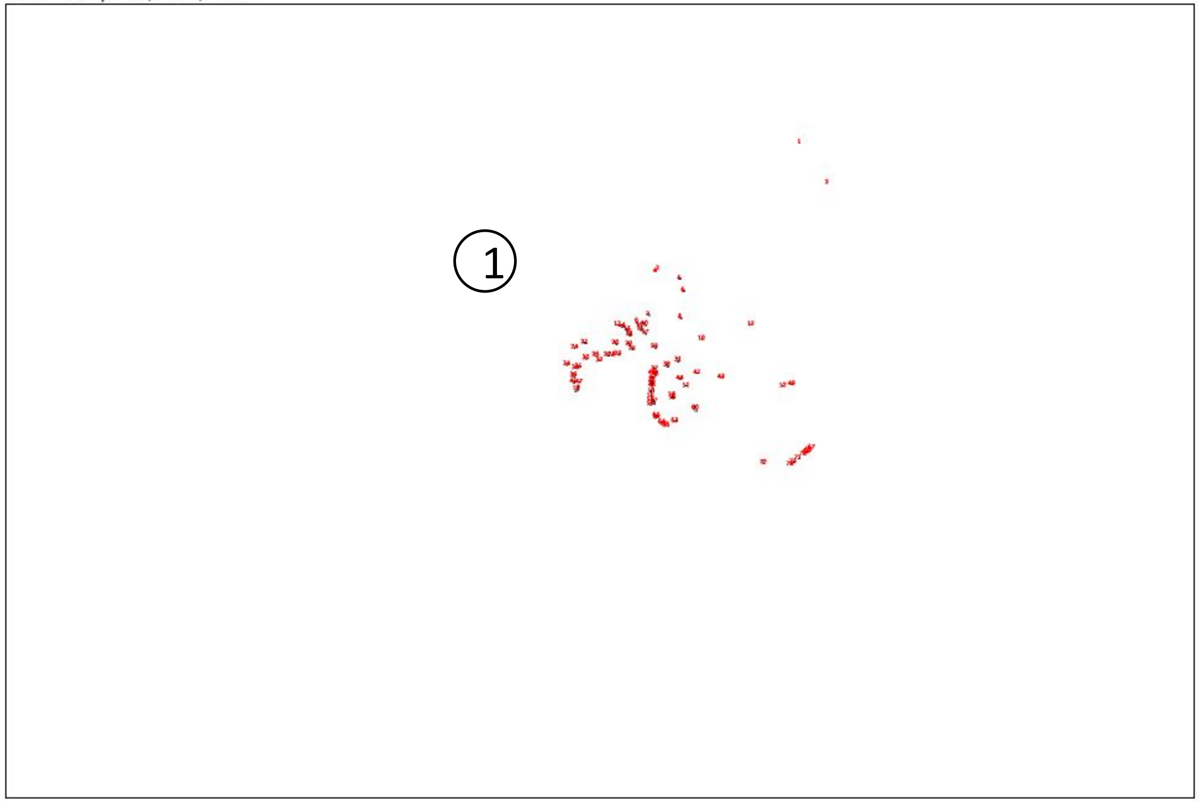


1920x1280 pixels; 8-bit; 2.3MB

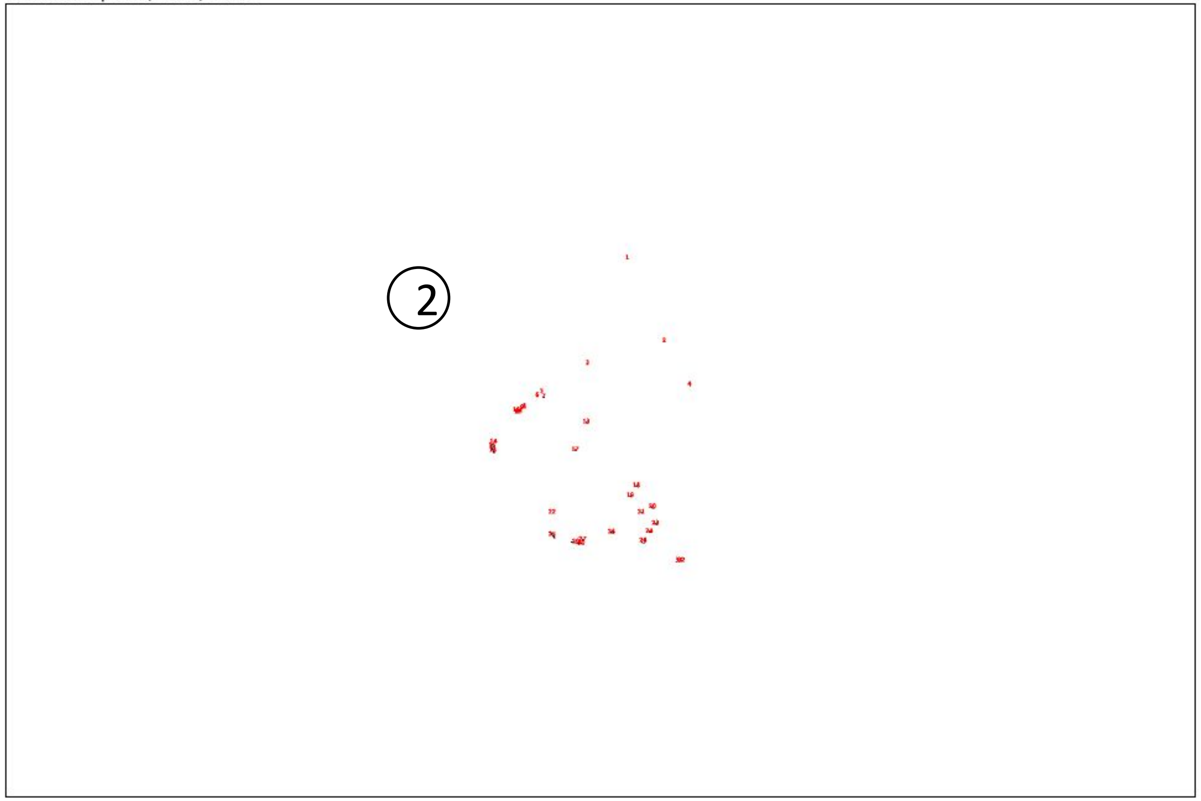


Result:

| | Slice | Count | Total Area | Average Size |
|---|---|---|---|---|
| ① | Scenario 5.3.jpg | 74 | 583 | 7.878 |
| ② | Scenario 6.3.JPG | 32 | 206 | 6.438 |

***(Unit of area: PIXEL)***

*Count = number of areas*
*Total Area = total size of all areas*
*Average Size = average size of each area*

*Unit of area: PIXEL*

### Scenario 7.1 VS 8.1

*Size of the picture is 1920x1280 pixels*

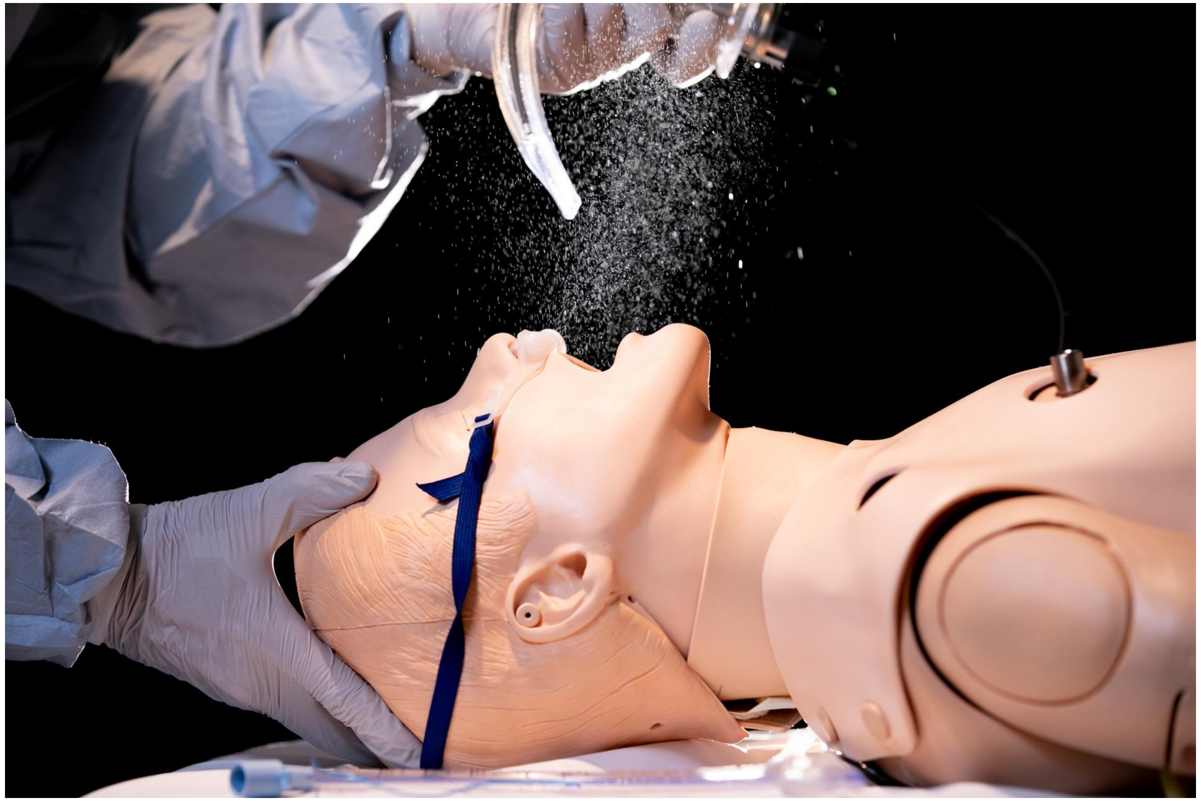

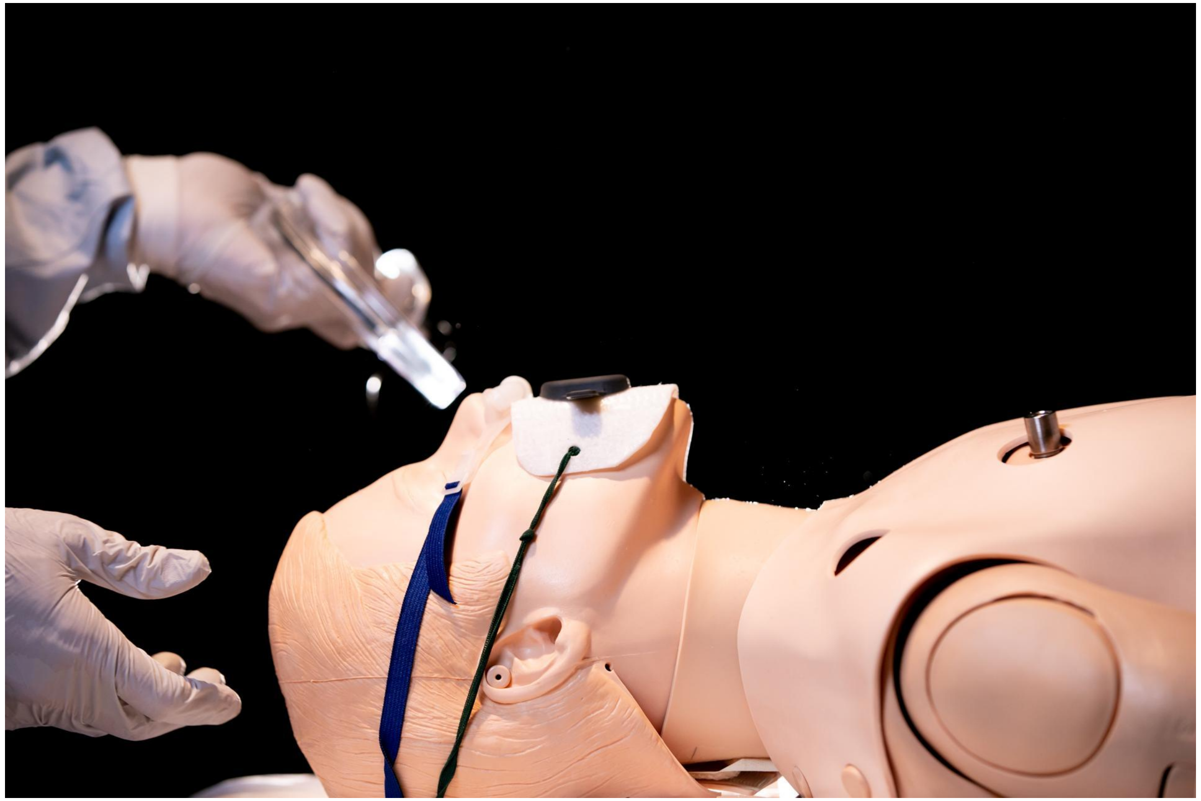

### Scenario 7.1 VS 8.1

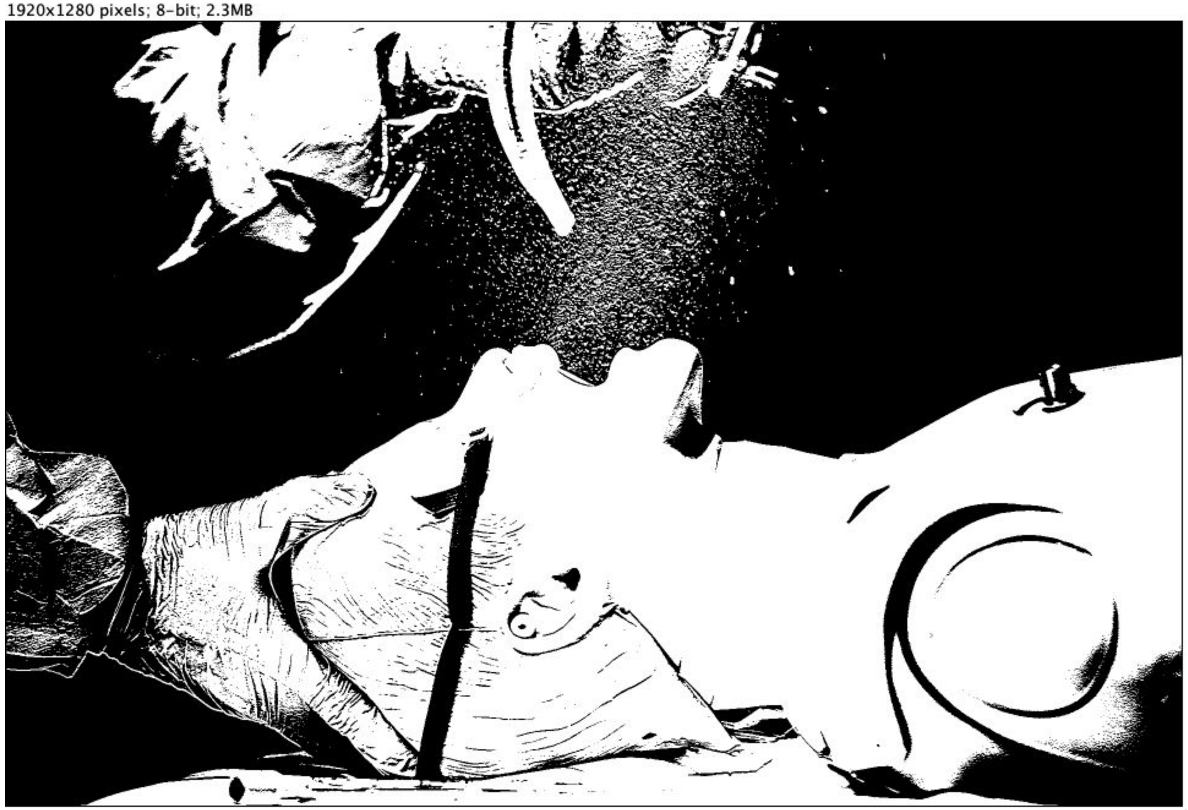


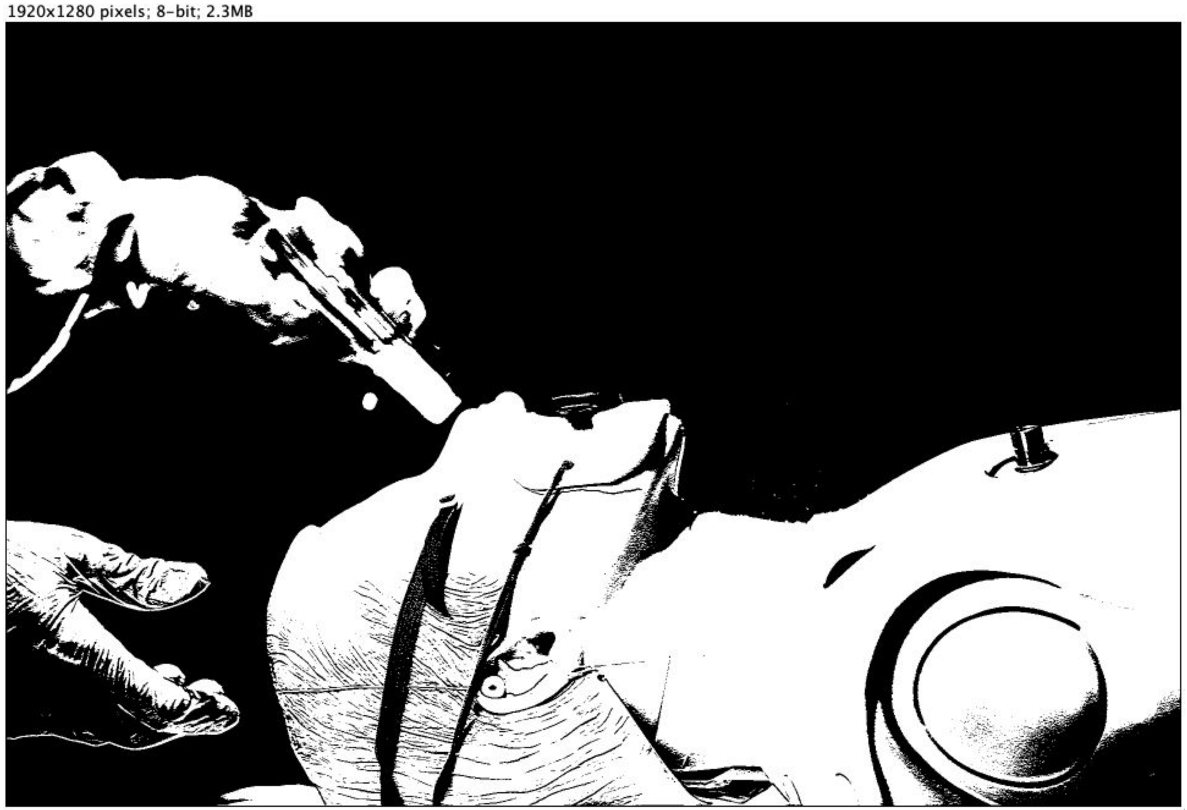

### Scenario 7.1 VS 8.1

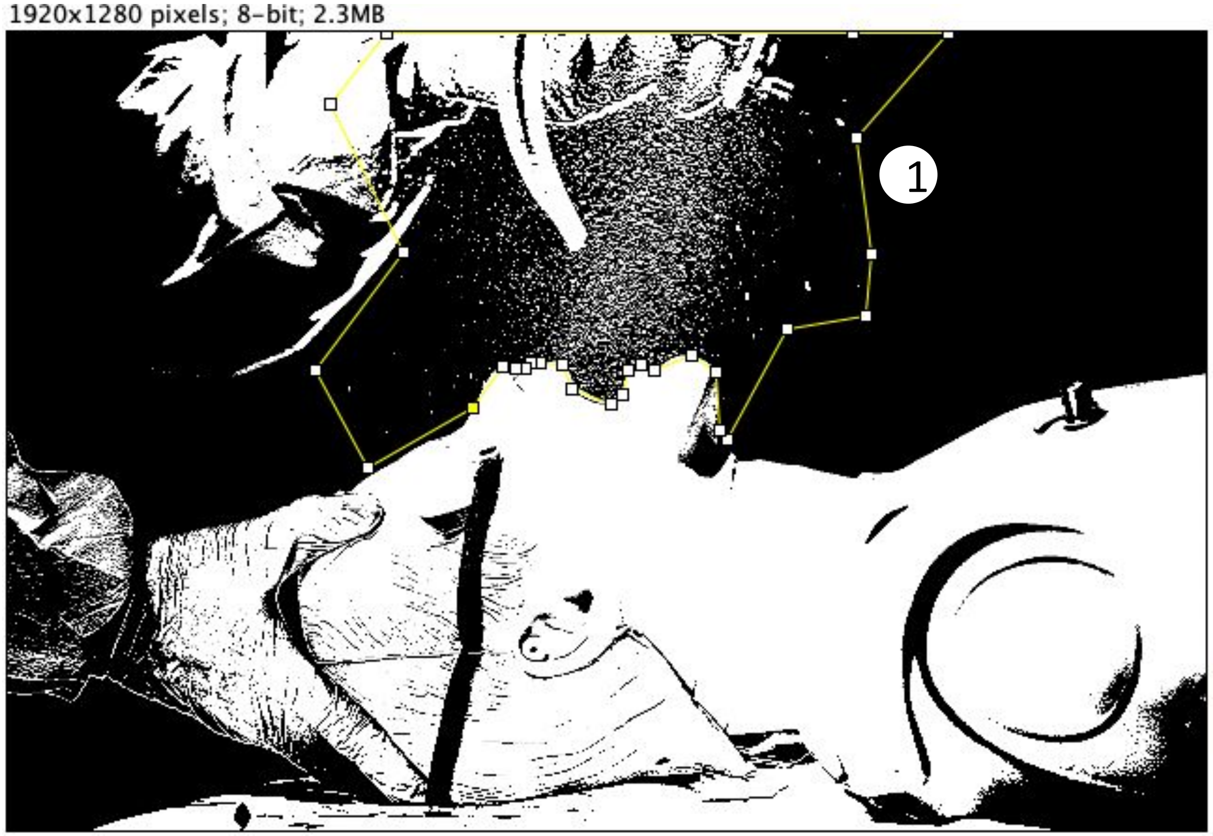


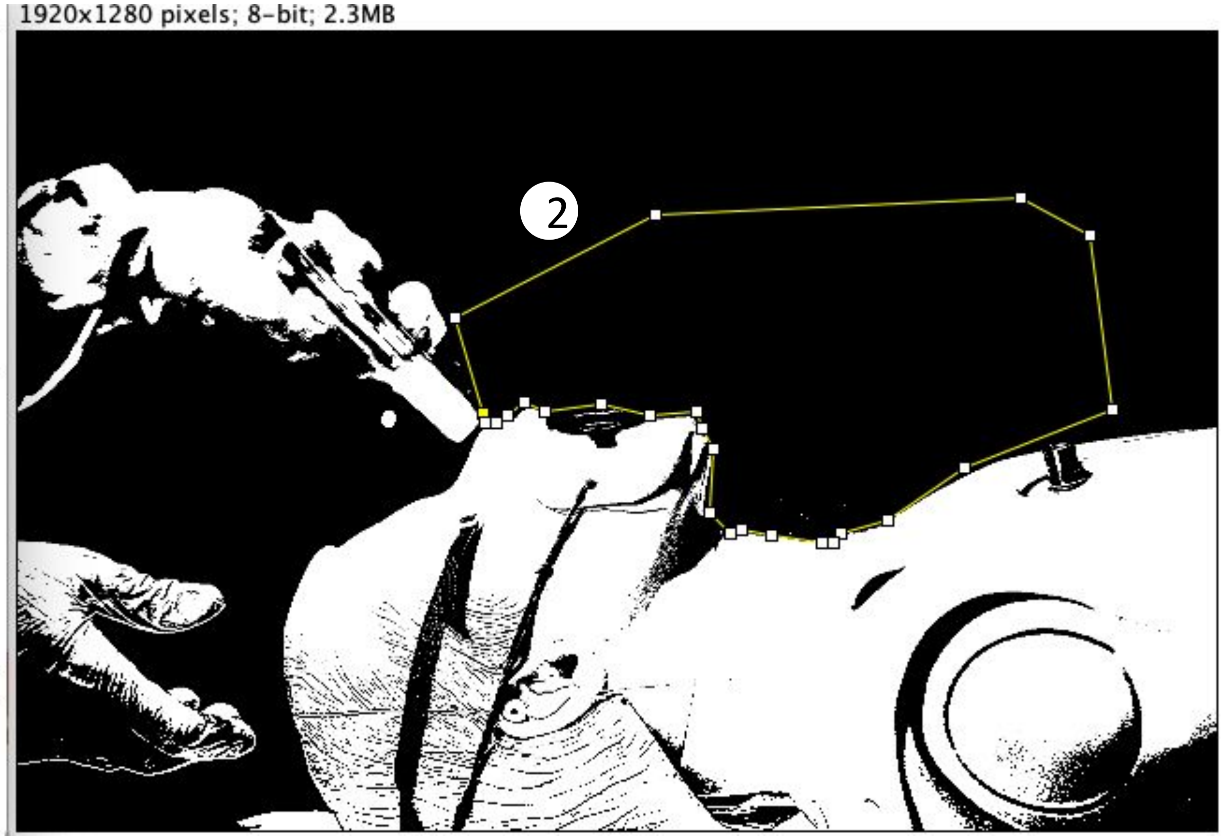

### Scenario 7.1 VS 8.1

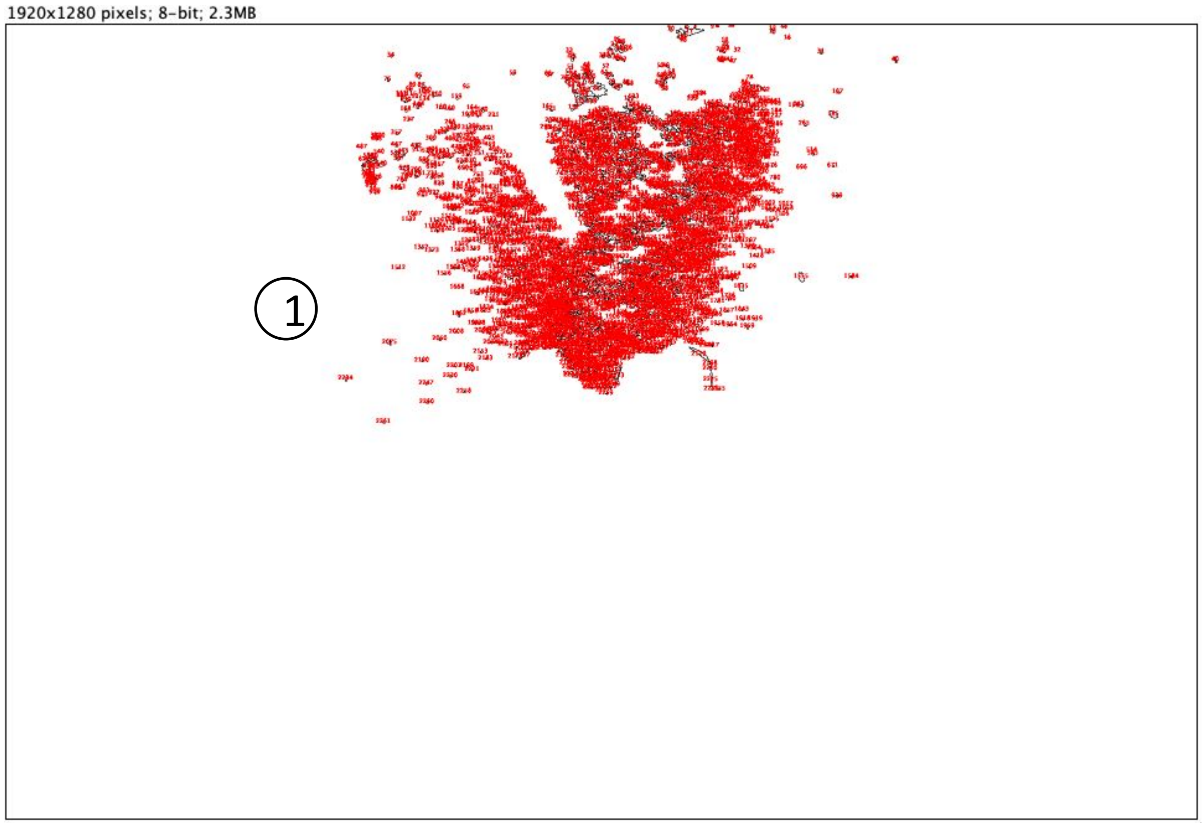


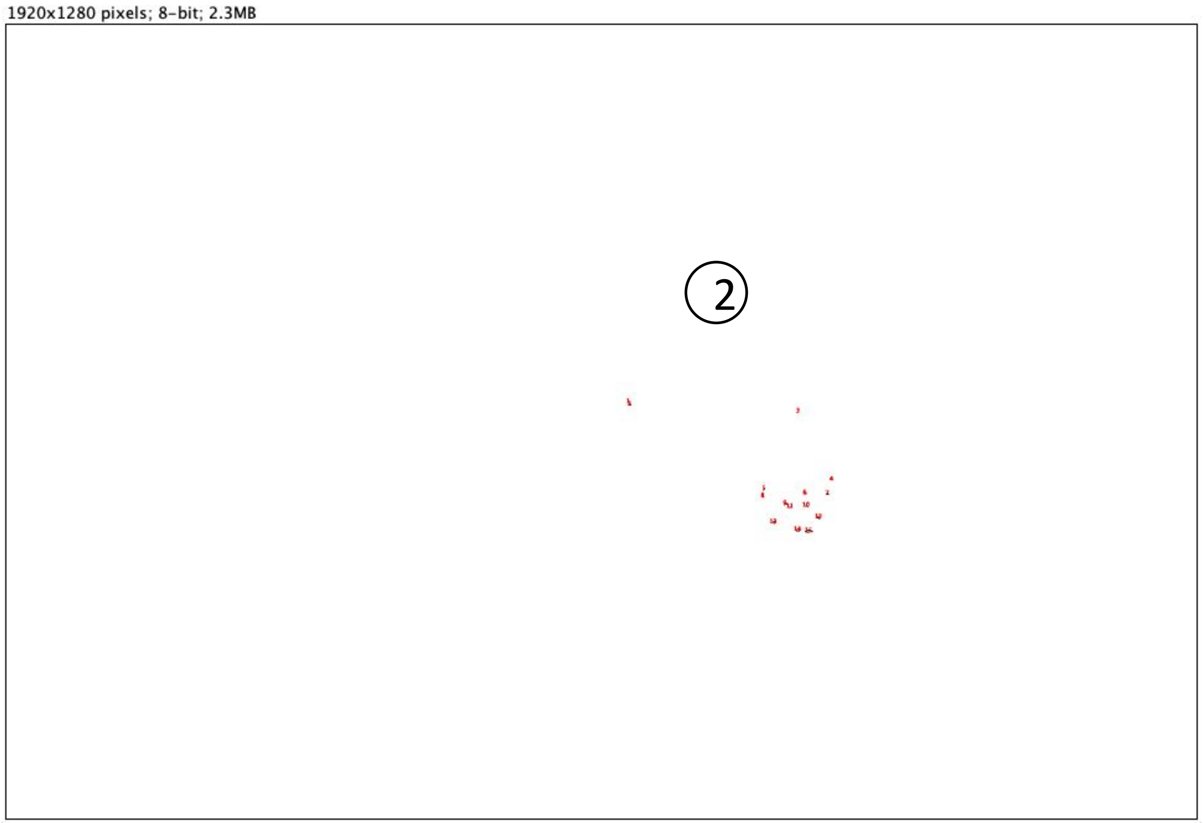


Result:

| | Slice | Count | Total Area | Average Size |
|---|---|---|---|---|
| 1 | Scenario 7.1.JPG | 2261 | 37142 | 16.427 |
| 2 | Scenario 8.1.JPG | 15 | 78 | 5.200 |

*(Unit of area: PIXEL)*

*Count = number of areas*
*Total Area = total size of all areas*
*Average Size = average size of each area*

*Unit of area: PIXEL*

### Scenario 7.2 VS 8.2

*Size of the picture is 1920x1280 pixels*

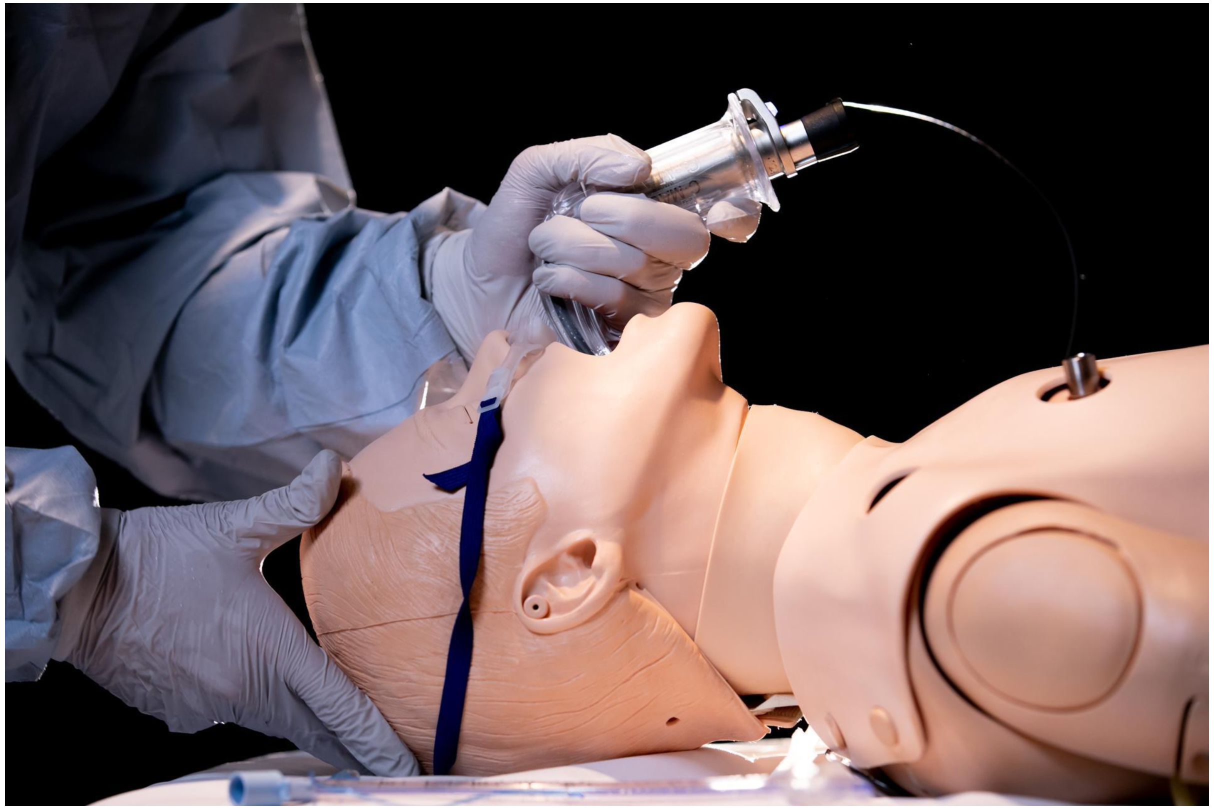

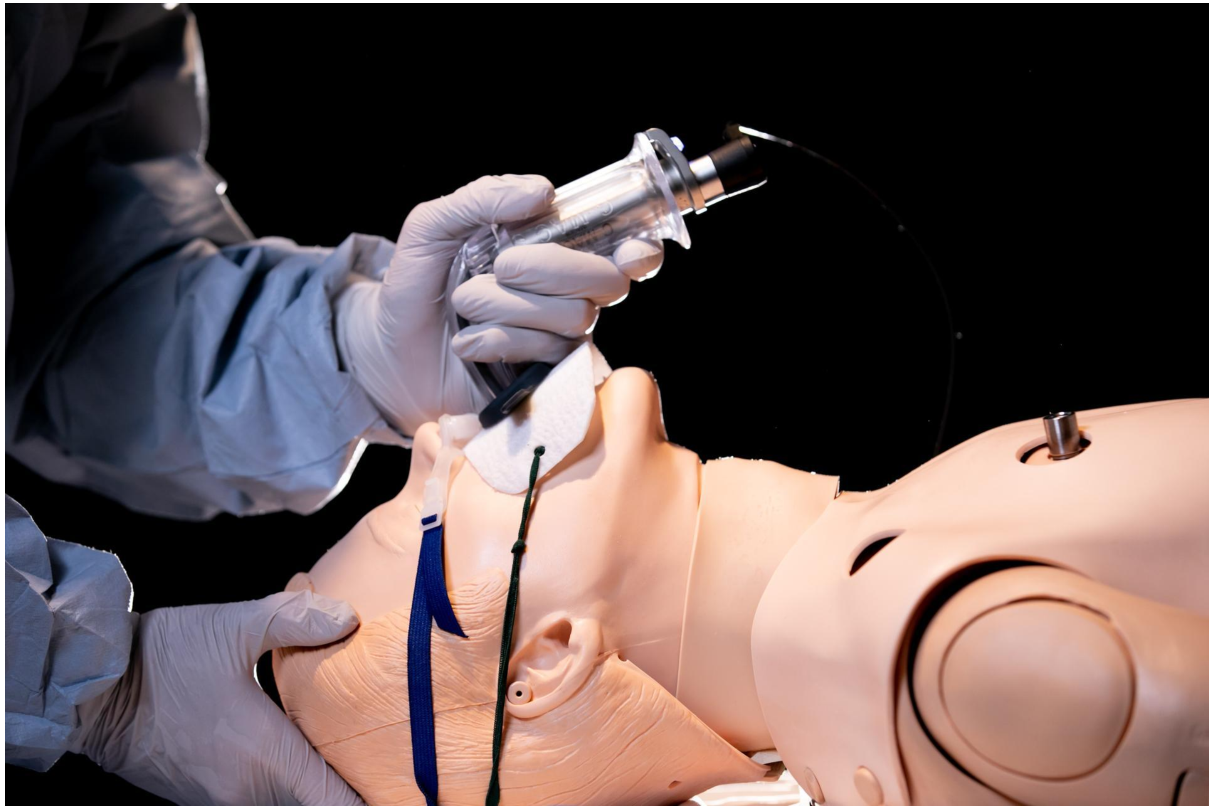

### Scenario 7.2 VS 8.2

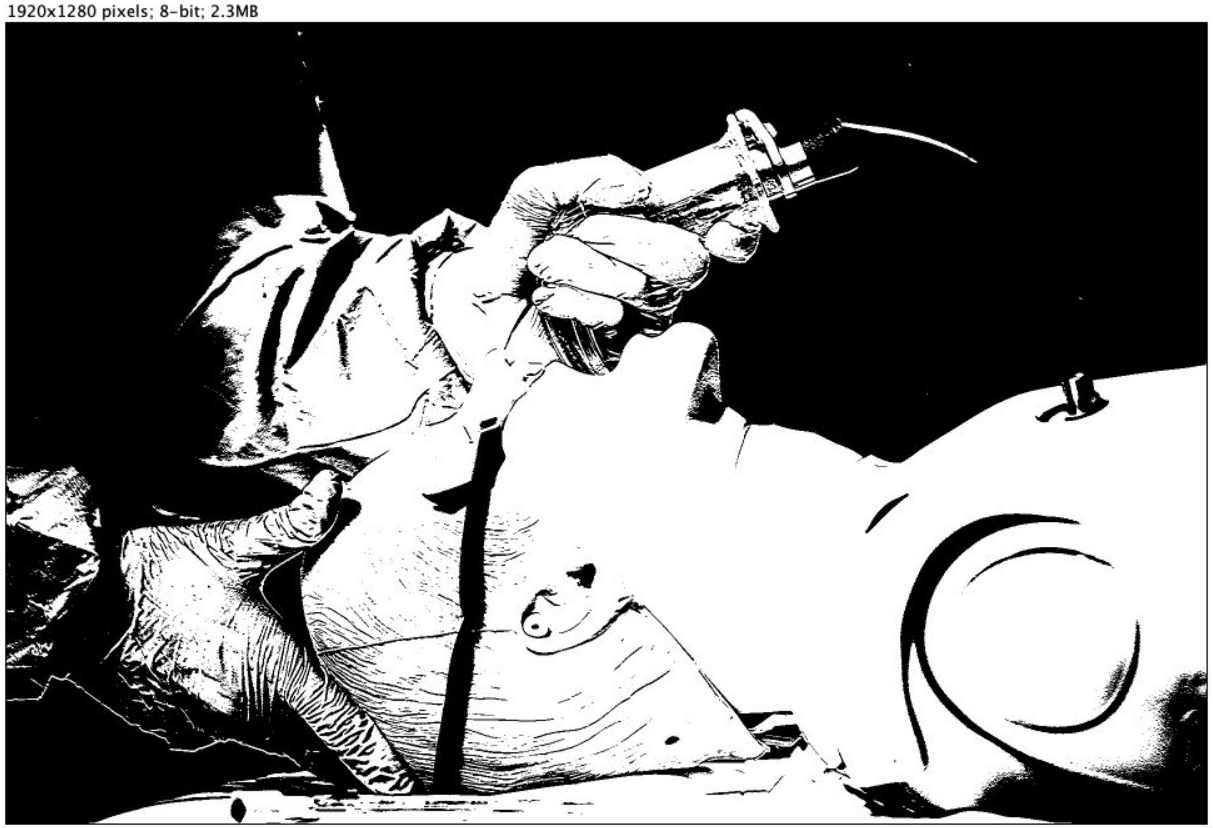


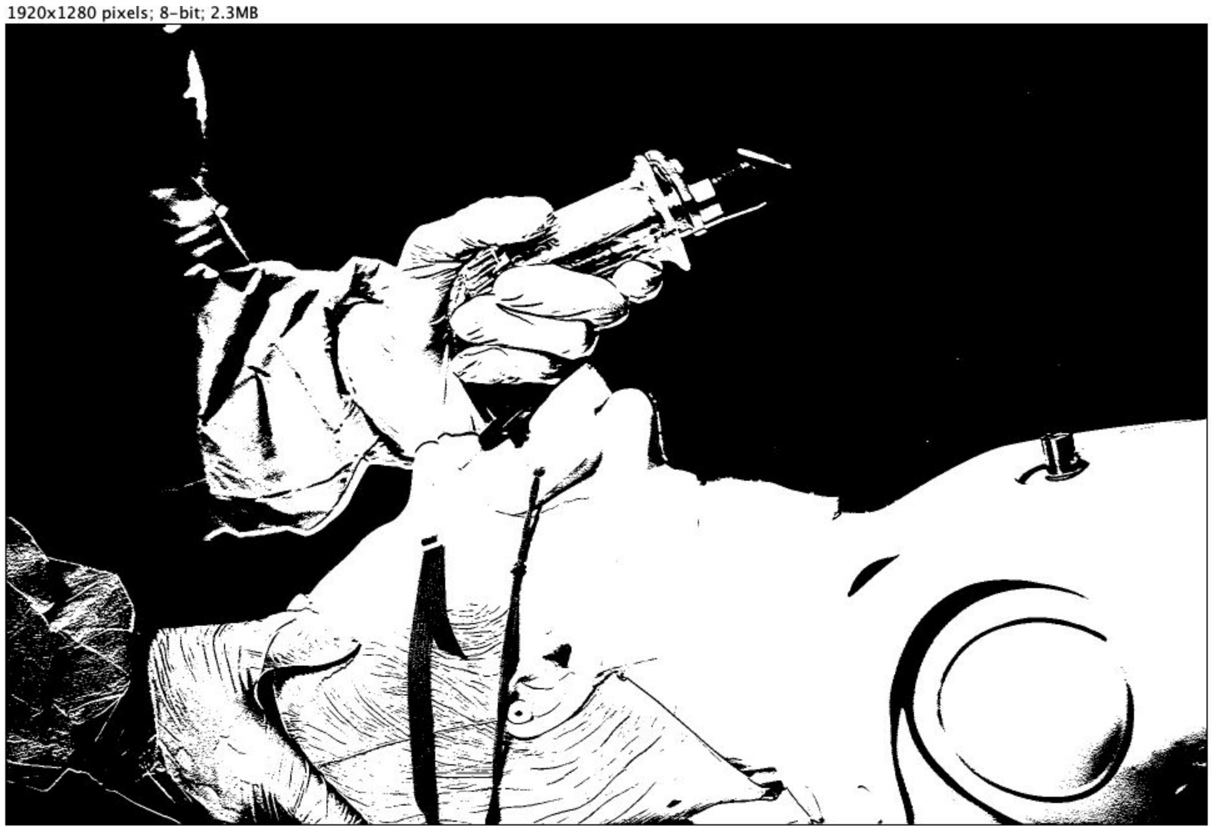

### Scenario 7.2 VS 8.2

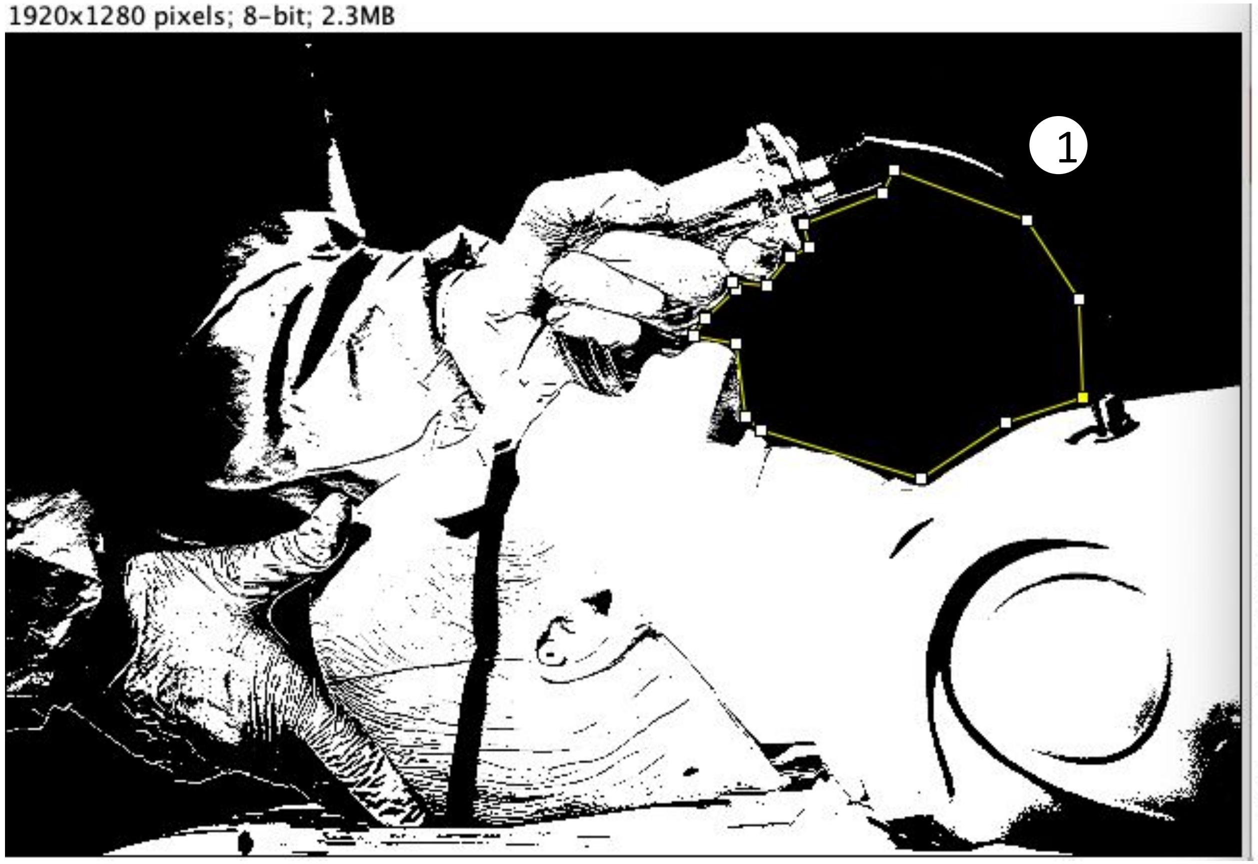


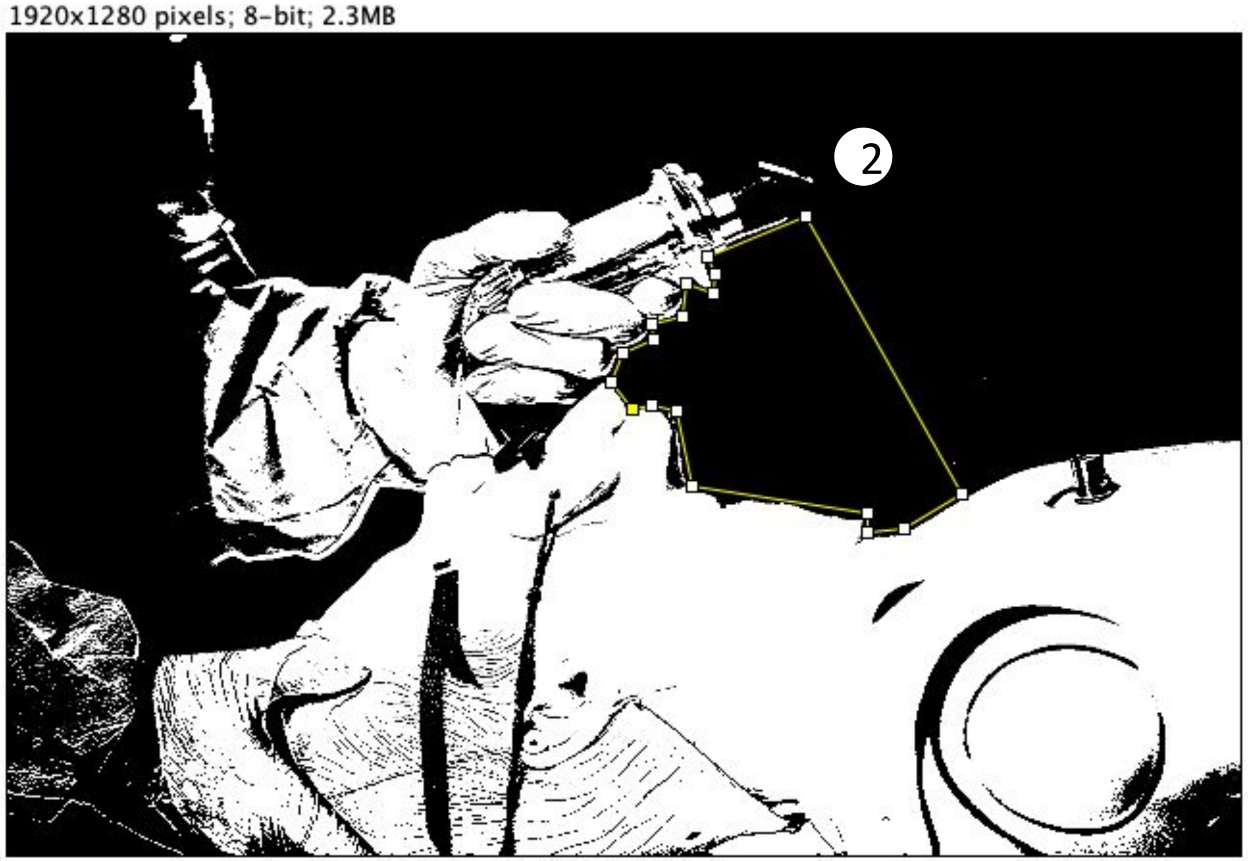

### Scenario 7.2 VS 8.2

1920x1280 pixels; 8-bit; 2.3MB

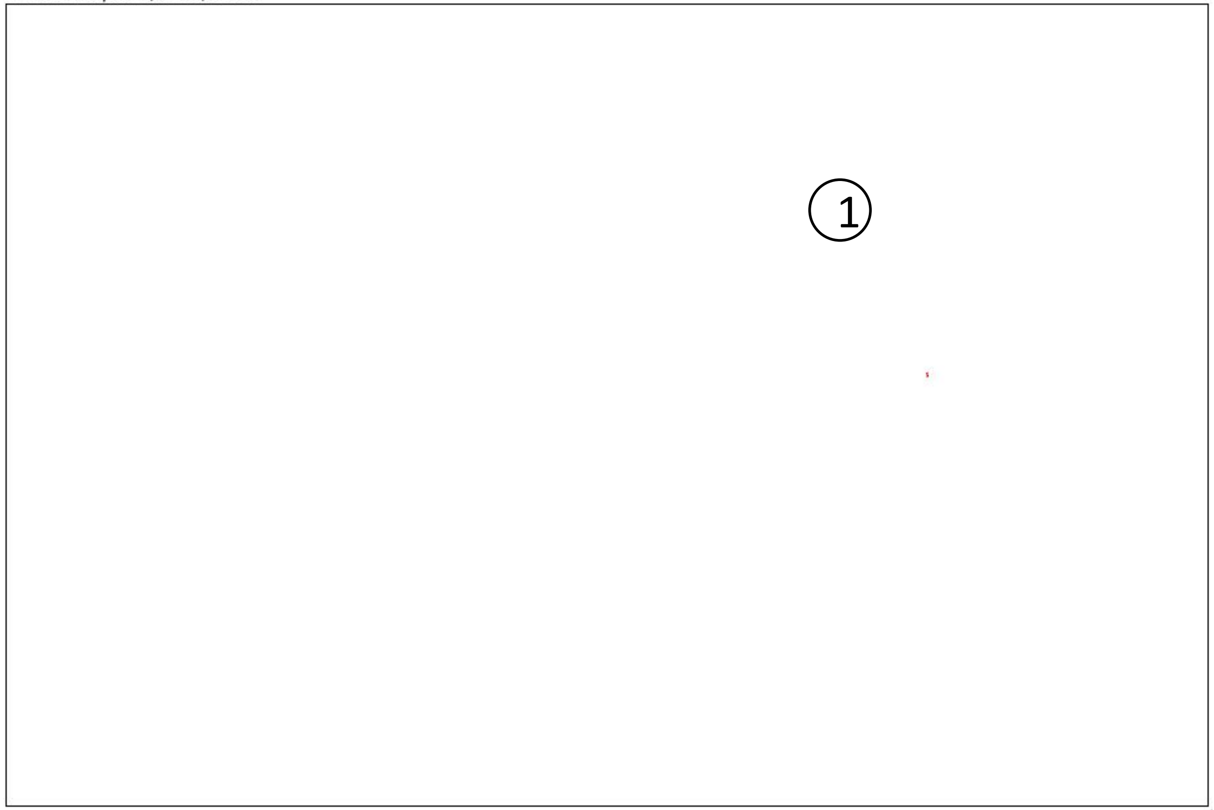


1920x1280 pixels; 8-bit; 2.3MB

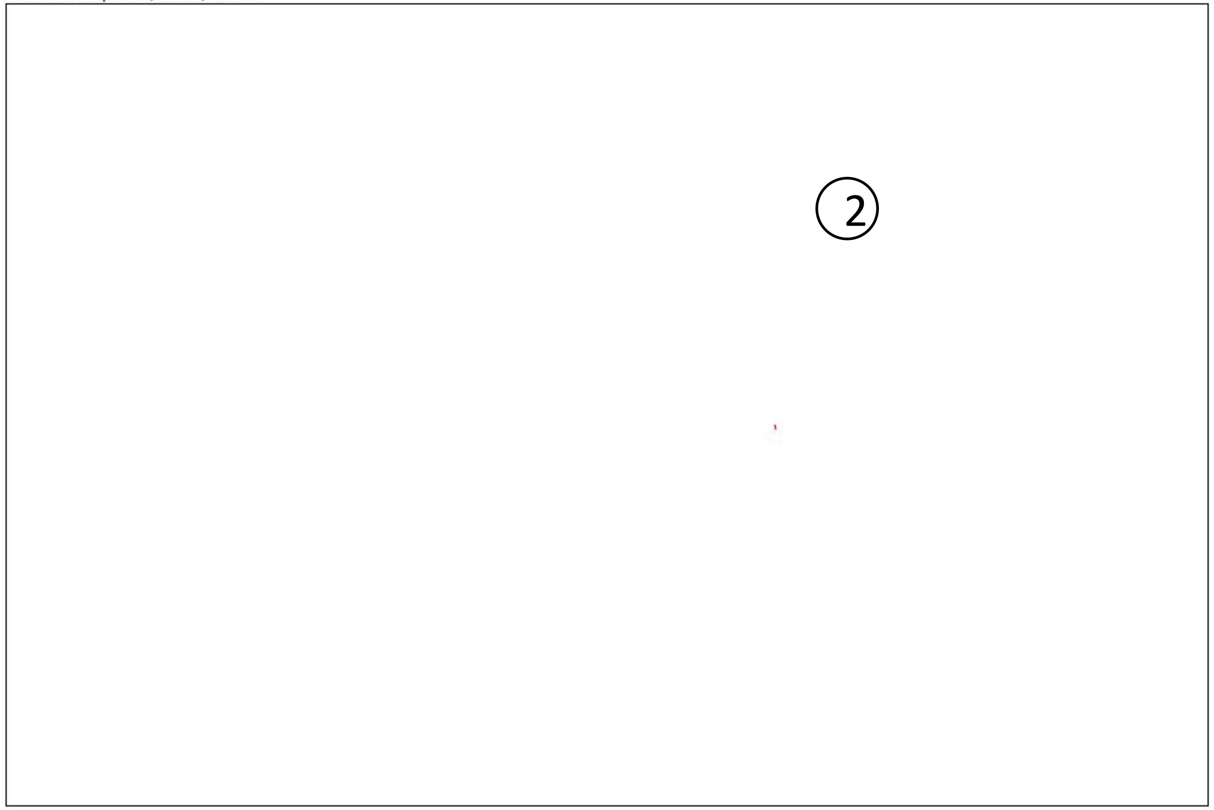


Result:

| | Slice | Count | Total Area | Average Size |
|---|---|---|---|---|
| ① | Scenario 7.2.tif | 1 | 2 | 2 |
| ② | Scenario 8.2.tif | 1 | 1 | 1 |

***(Unit of area: PIXEL)***

*Count = number of areas*
*Total Area = total size of all areas*
*Average Size = average size of each area*

*Unit of area: PIXEL*

### Scenario 7.3 VS 8.3

*Size of the picture is 1920x1280 pixels*

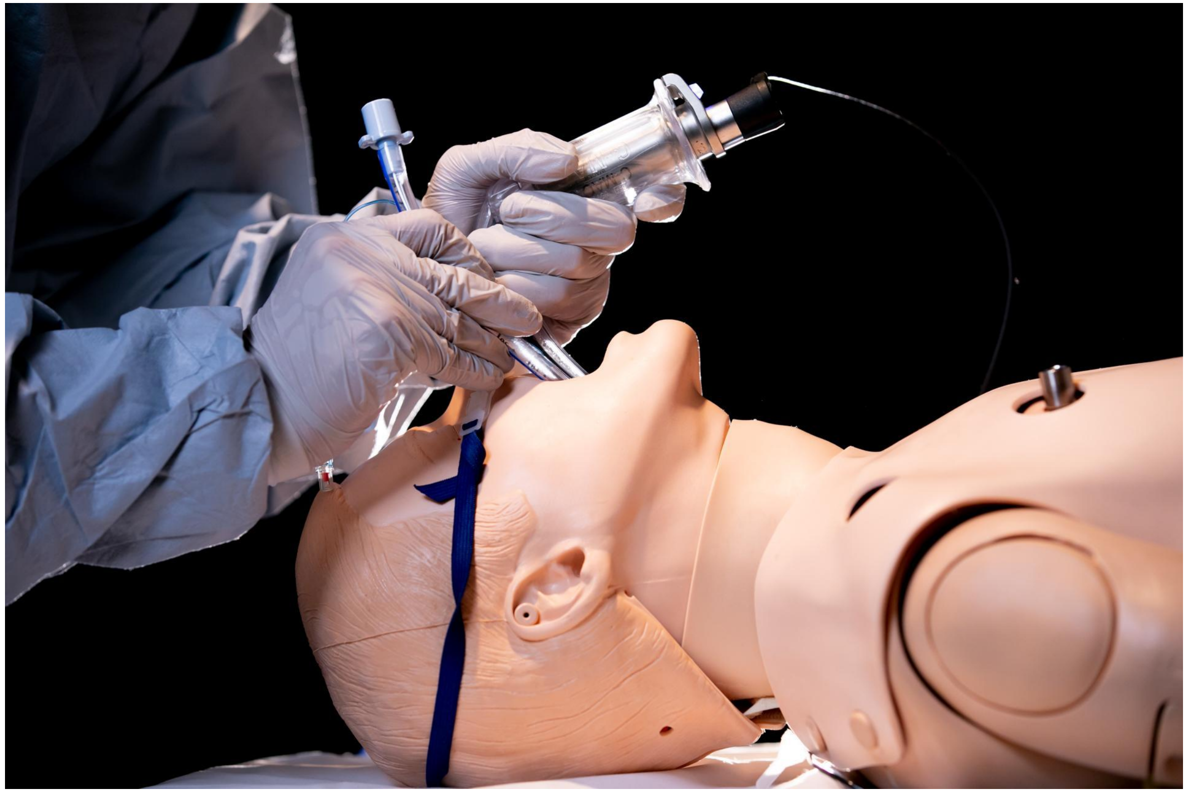

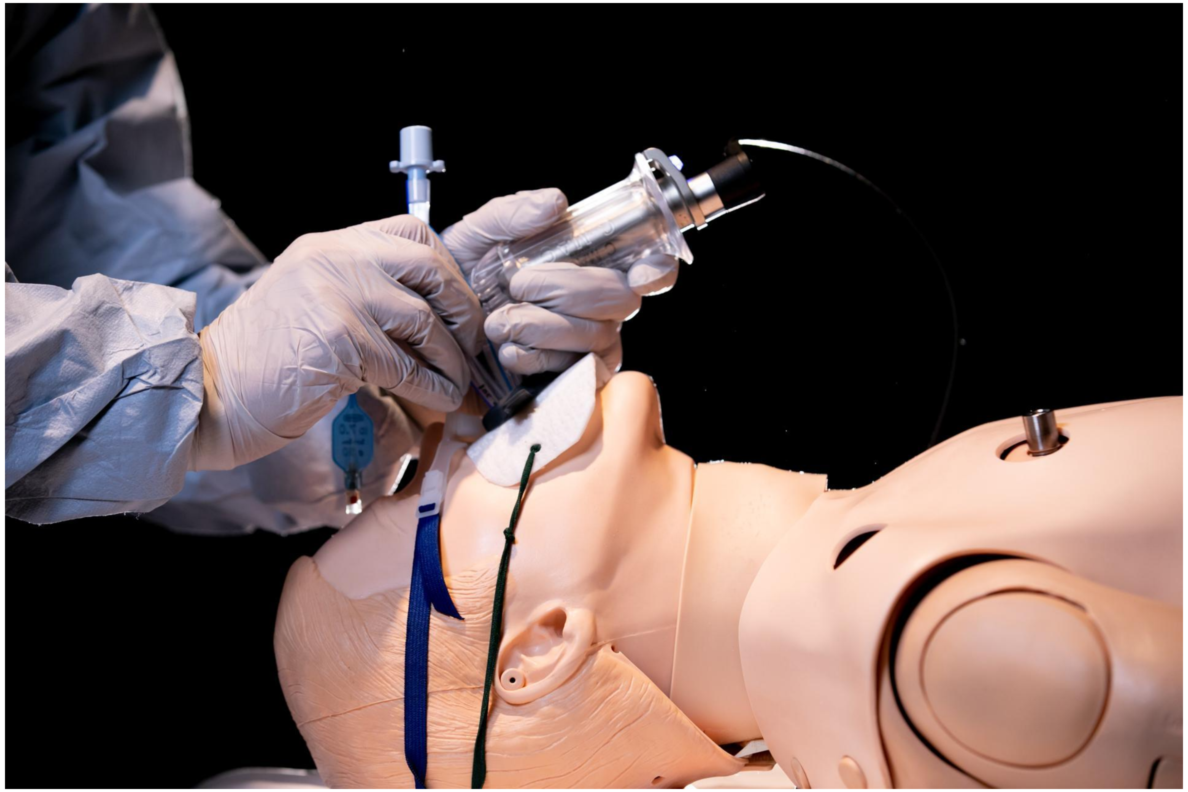

### Scenario 7.3 VS 8.3

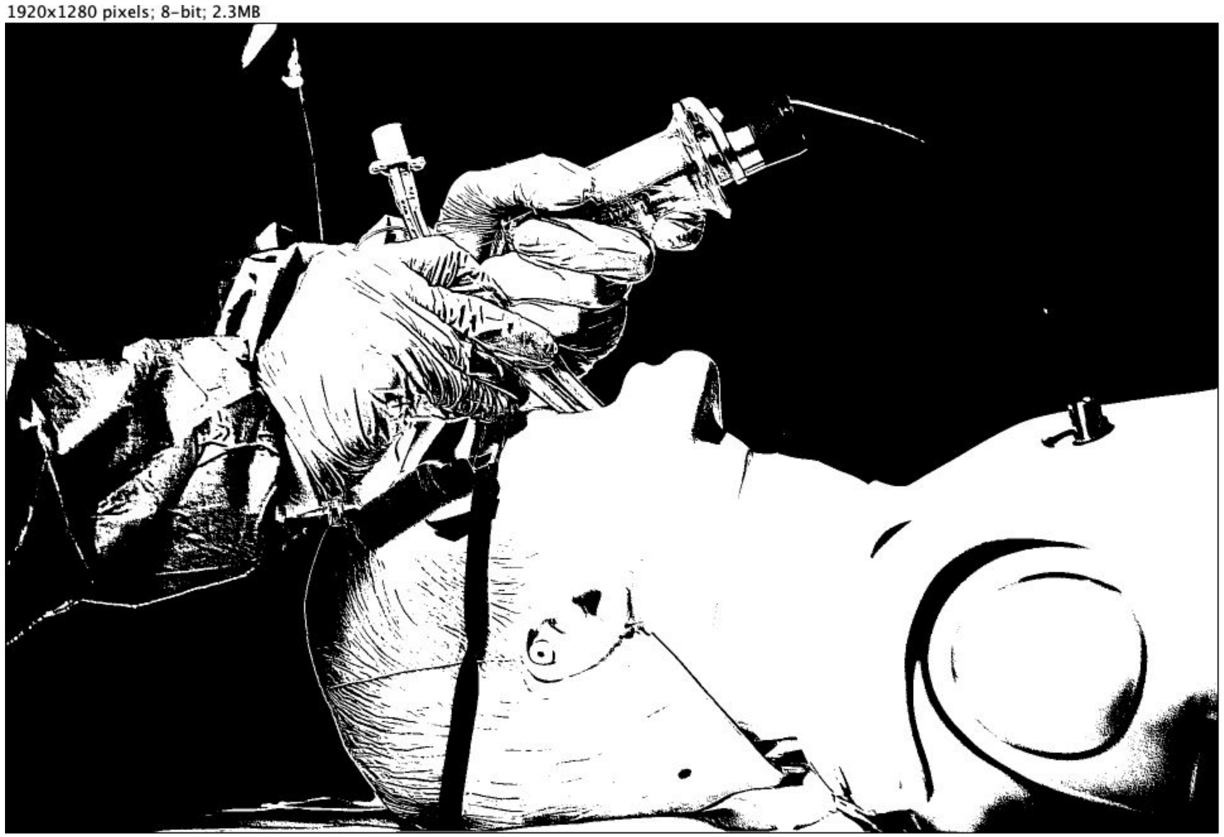


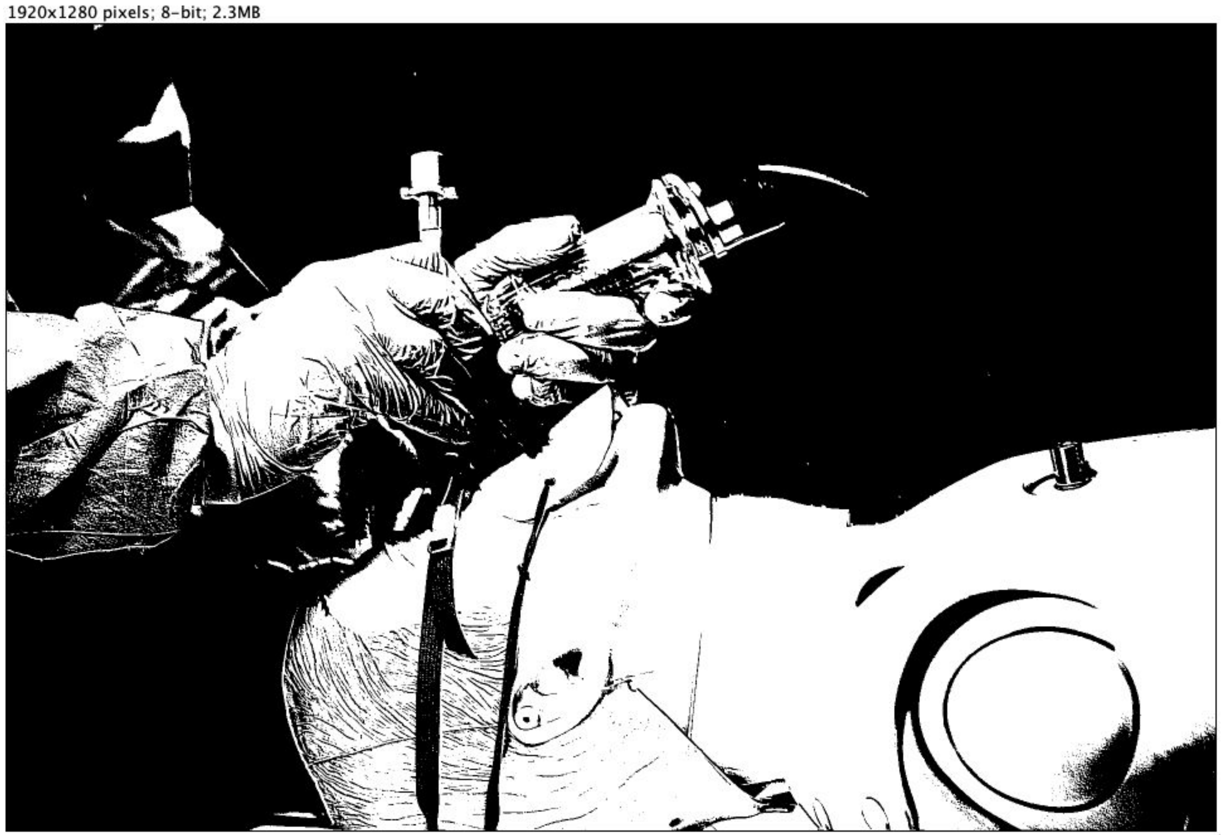

Scenario 7.3 VS 8.3

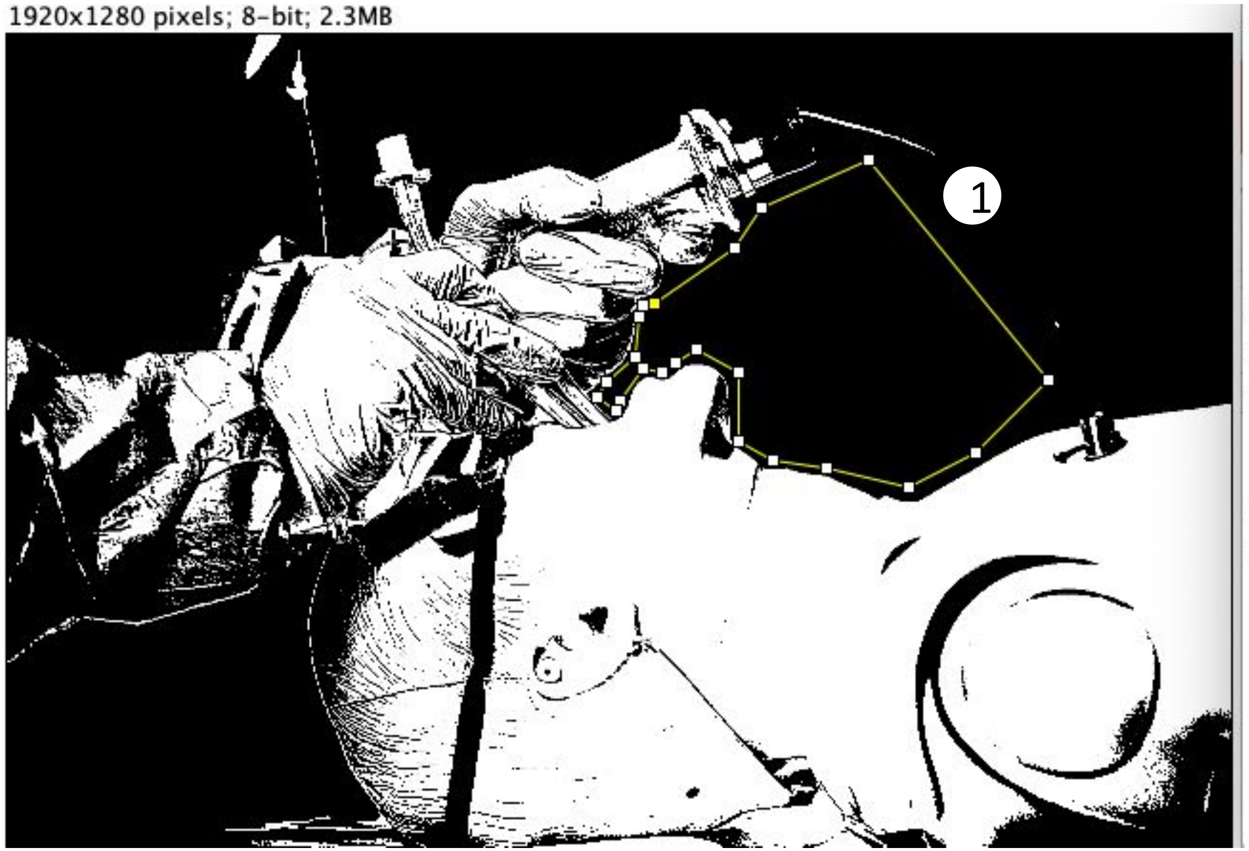


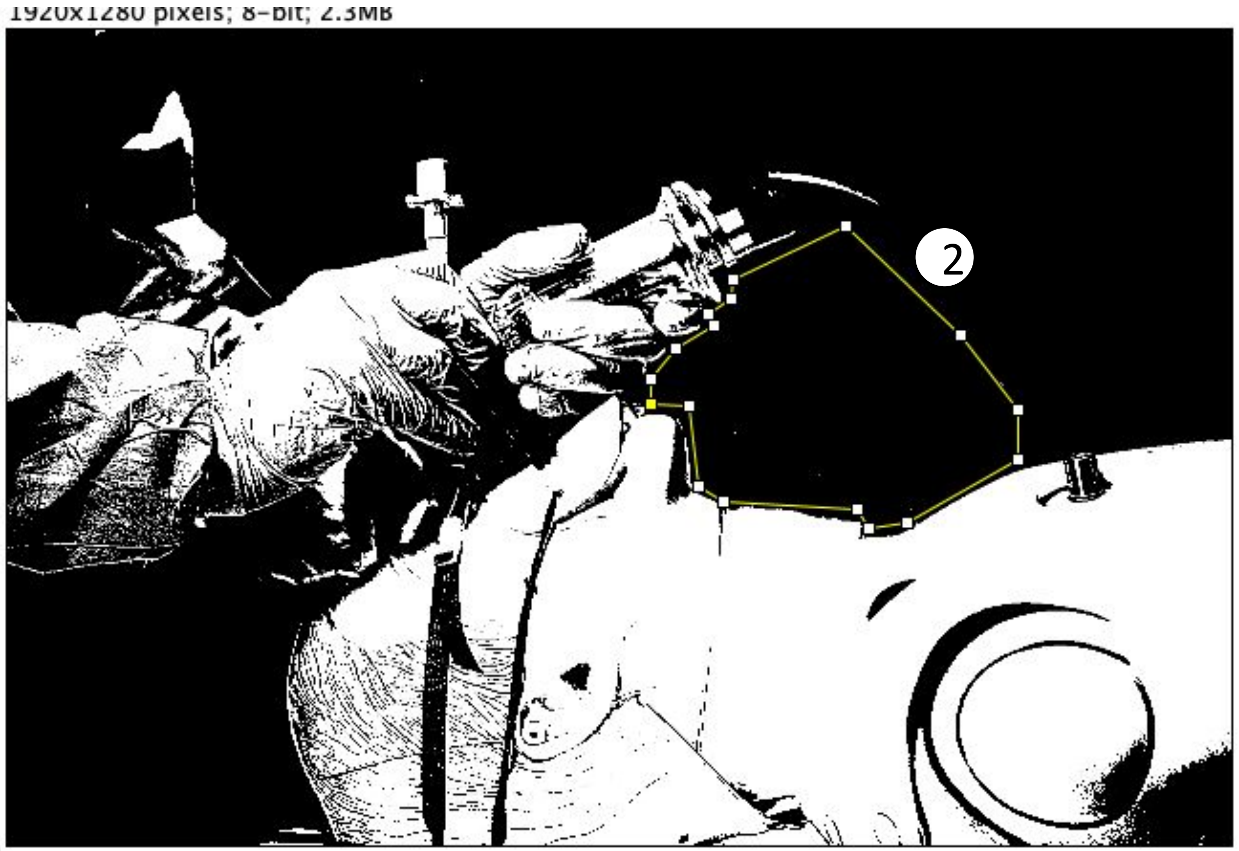

### Scenario 7.3 VS 8.3

1920x1280 pixels; 8-bit; 2.3MB

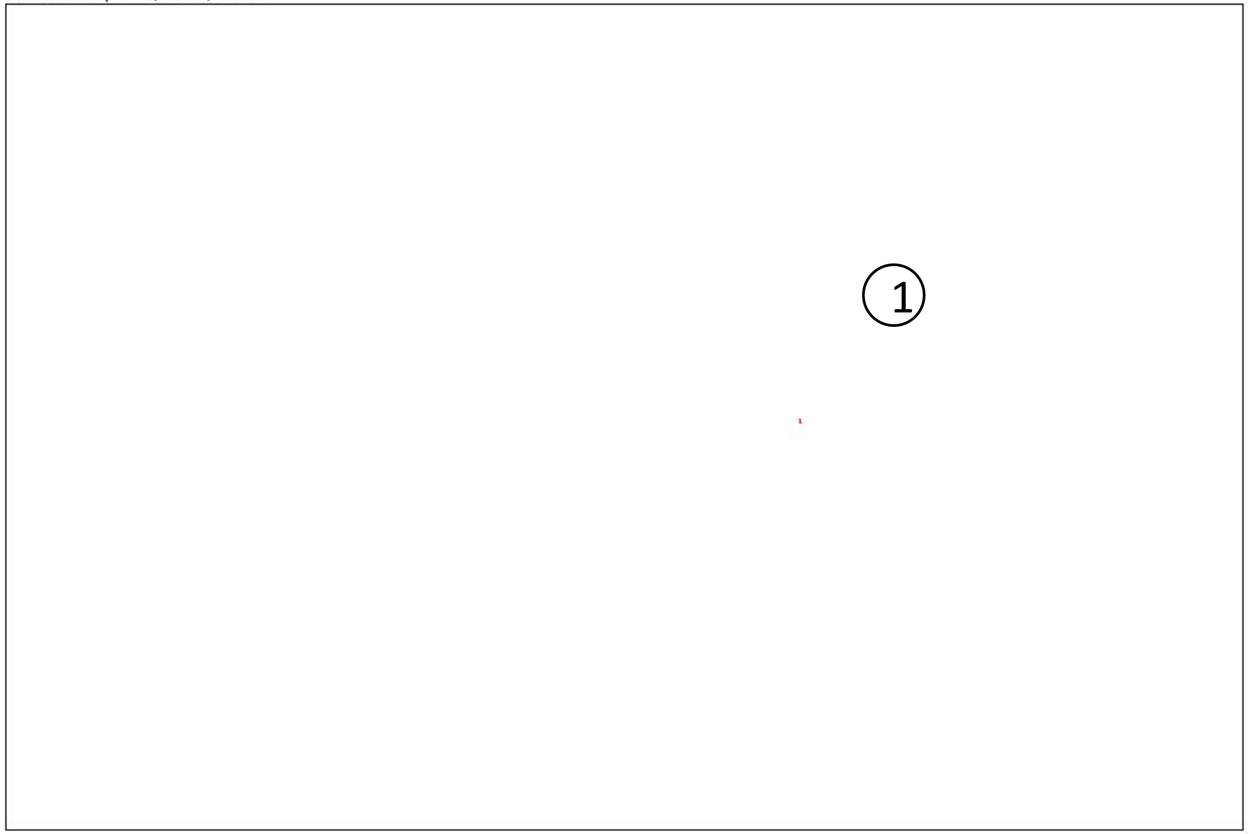


1920x1280 pixels; 8-bit; 2.3MB

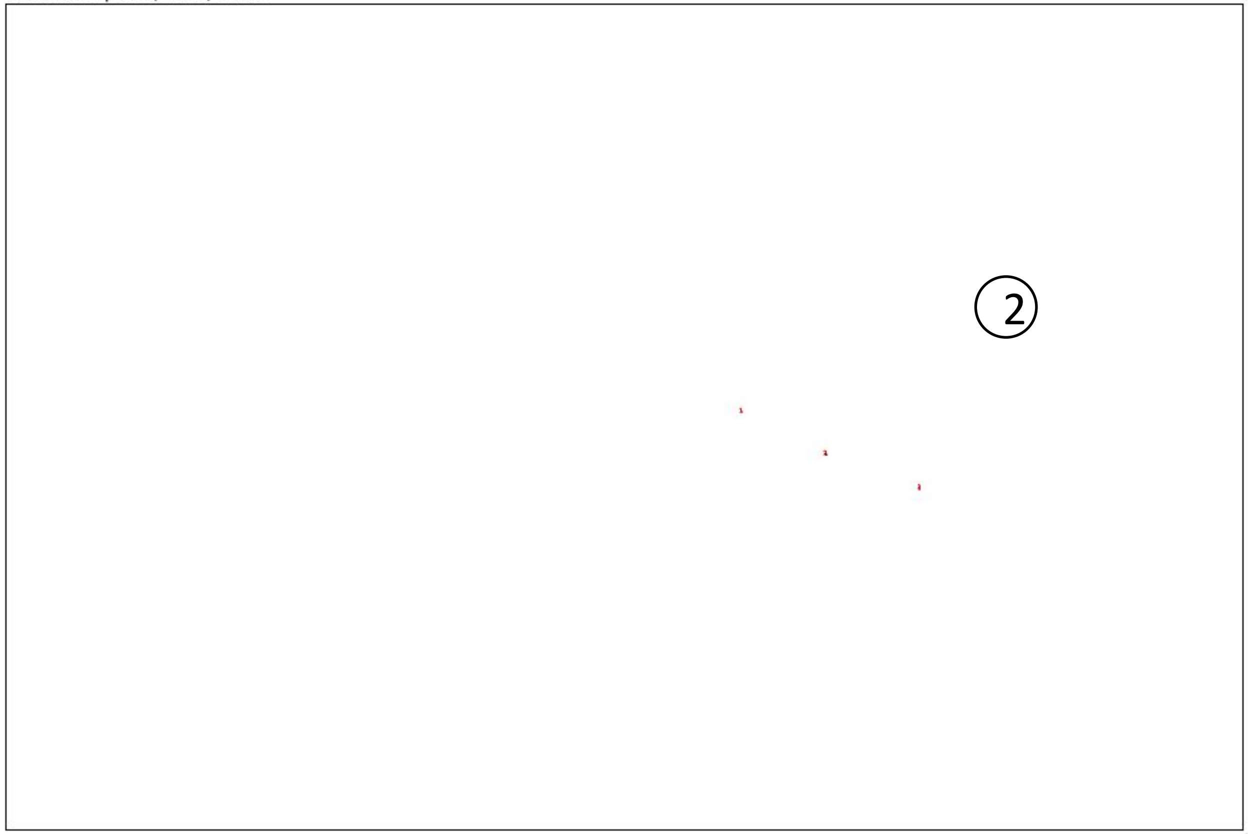


Result:

| | Slice | Count | Total Area | Average Size |
|---|---|---|---|---|
| 1 | Scenario 7.3.JPG | 1 | 2 | 2.000 |
| 2 | Scenario 8.3.JPG | 3 | 16 | 5.333 |

***(Unit of area: PIXEL)***

*Count = number of areas*
*Total Area = total size of all areas*
*Average Size = average size of each area*

*Unit of area: PIXEL*

### Study Limitations:

- The interference of the background light may affect the scope of measurement
- Manually polygon selections may affect the accuracy of the data
- The flow of aerosols and droplets set on the mannequin cannot always be consistent

*More details are provided in main text.*

### Software used for measurement: **ImageJ**

Version: 2.1.0/1.53c

Build: 5f23140693

Date: 2020-08-02T03:43:22+0000

Copyright 2010-2021

https://imagej.net/Fiji

All photos are 1920x1280 pixels
